# Global Programmatic Survey on Governance and Surveillance of Nontuberculous Mycobacteria

**DOI:** 10.64898/2026.07.27.26358874

**Authors:** Nityanand Jain, Liga Kuksa, Dissou Affolabi, Fabiola Eliosa Arias Munoz, Angelica Scappaticcio, Sushil Pandey, Lisa Shephard, Rumina Hasan, Lorenzo Guglielmetti, Gert-Jan Wijnant, Emmanuel Andre, Leen Rigouts, Natalie Lorent, the NTM Global Policy Study Group

## Abstract

**Background:** Nontuberculous mycobacteria (NTM) are clinically important pathogens but often weakly positioned within health systems, with unclear institutional ownership, variable notification arrangements, and limited surveillance visibility.

**Methods:** We conducted a multilingual online survey among programme-facing national and subnational mycobacterial stakeholders from November to December 2025. Using an adaptive hierarchical recruitment strategy, we targeted 217 countries and jurisdictions. One response per programme was requested. We assessed institutionalisation, notification, agenda maturity, thematic discussion priorities, and barriers to action. We also derived relative policy momentum from three urgency domains (diagnostics, clinical management, and surveillance) using within-WHO region hierarchical clustering. Findings were used to develop a heuristic roadmap for staged NTM governance and system visibility.

**Results:** We received 193 programme-level responses, including from subnational jurisdictions, representing 171 of 217 targeted countries (78.8% jurisdictional coverage). NTM institutionalisation status was heterogeneous: 34% reported NTM integration within the NTP, while 37% expressed intents to institutionalise within the NTP in nearest future. Mandatory notification was reported by 21% of programme units, voluntary notification by 10%, and notification under consideration by 24%. NTM appeared to enter policy discussions along a gradient, with clinical management and diagnostics attracting earlier attention than surveillance, training needs, and financing. Globally, pulmonary NTM was discussed more frequently than extrapulmonary disease. Across WHO regions, policy momentum clustering separated programmes into higher and lower profiles, with surveillance consistently being the weakest domain. Financing and lack of epidemiological data were identified as the most actionable barriers. There was broad support for TB-NTM surveillance integration.

**Conclusions:** NTM governance is heterogeneous and frequently weakly anchored globally. The findings do not support a single universal institutional model; rather, existing mycobacterial platforms may provide pragmatic starting points for improving programme visibility, coordination, and reporting. The proposed heuristic roadmap outlines staged governance options according to burden, capacity, and institutional context.

## 1. Introduction

Nontuberculous mycobacteria (NTM) are a heterogeneous group of ubiquitous environmental mycobacteria found in water, soil, and built environments. Environmental exposure is presumed to be common, although disease occurs mainly in susceptible hosts [1,2]. As opportunistic pathogens, they can cause chronic, often difficult-to-manage disease across a broad clinical spectrum, from pulmonary infection to skin, soft tissue, lymphatic, and disseminated disease [3–5]. Diagnosis is challenging as symptoms are often non-specific, microbiological isolation does not always indicate active disease, and interpretation depends on repeated sampling and species identification [6,7]. Clinical management is similarly complex, often requiring prolonged and toxic antimicrobial therapy and specialist follow-up [8]. These features place NTM in an unusual position – widely encountered in the environment, clinically consequential in selected patients, and heavily dependent on specialised laboratory and clinical interpretation.

Over the past decade, reported NTM burden has appeared to increase in several settings, especially in higher-income and industrialized countries [9,10]. Japan is often cited as an example where pulmonary NTM disease has become more prevalent than tuberculosis (TB), as TB incidence continues to decline [11]. Whether this reflects changing microbial epidemiology, increasing host susceptibility (e.g., ageing populations, immunosuppression), increased awareness, or improvements in surveillance capacity remains uncertain. Nonetheless, the policy relevance of NTM does not depend on burden trends alone. In many settings, limited mycobacterial diagnostic capacity and frontline awareness also mean that patients with possible NTM disease may not undergo further species-level investigation after initial negative (molecular) TB testing and may instead remain caught in empiric TB care pathways [12,13].

This could lead to delayed diagnosis, limited visibility in routine systems, and missed opportunities to distinguish NTM from non-responding ‘chronic TB’ [12,13]. Clearer institutional ownership, along with more structured mechanisms for aggregation or reporting where feasible, may help move NTM from a dispersed clinical problem to a more visible health-system issue. Improved system visibility could support more comparable burden estimation, more rational use of mycobacterial laboratory capacity, and clearer identification of where referral and specialist expertise are needed. For mycobacterial reference laboratories and TB programmes, such arrangements may also provide a pragmatic way to connect microbiological and clinical signals that are already being encountered but not routinely linked. Importantly, in this study, we considered surveillance primarily as a mechanism for system visibility, burden estimation, service planning, and coordination; not necessarily as a TB-equivalent strategy for controlling person-to-person transmission.

Much of the existing NTM literature has focused on epidemiology, risk factors, and clinical management, repeatedly highlighting the need for better global burden estimates, diagnostics, and surveillance [9,10]. Yet a more fundamental implementation question remains underexplored: who, in practical programme terms, is meant to act on this agenda? Clinical relevance alone does not generate policy response. Without clear responsibility, reporting pathways, and capacity to respond, NTM may remain visible as a localized problem without having scalable disease management and control measures. Corollary to this framing, the central unanswered question is not only how NTM enter clinical and surveillance systems, but where responsibility for recognizing, counting, and responding to NTM lies, or could realistically be anchored.

We acknowledge that the relevance and urgency of addressing NTM organization are unlikely to be perceived uniformly across countries. In some settings, it may remain secondary to immediate TB control needs, particularly where mycobacterial testing capacity is limited or dependent on external referral. Yet this variation is itself part of the problem. In a period of constrained TB financing and uneven diagnostic infrastructure [14,15], the key issue is not whether all jurisdictions should respond to NTM in the same way, but whether NTM is being recognized *at all*, where it is landing within health systems, and what conditions might support a more systematic response over time.

This makes a global programme-level perspective especially important. We, therefore, conducted a cross-sectional survey to (i) identify the programmatic organization of NTM within health systems, using mycobacterial stakeholders as the entry point; (ii) assess programme engagement through reported discussion, agenda-setting, and policy urgency; (iii) identify perceived barriers to action; and (iv) derive a heuristic, findings-based roadmap for how NTM governance and system visibility may evolve across settings. In doing so, we sought to generate a global baseline of how NTM are currently positioned within health systems, how programme attention and institutional responsibility are configured, and which governance pathways may be most feasible in different contexts.

## 2. Methods

From November to December 2025, we conducted an online questionnaire survey to characterise programme attention to NTM, map institutional and governance patterns, and identify reported prerequisites for broader policy engagement. Ethical approval was not required as the study was deemed a minimal-risk, non-interventional survey under local regulations as confirmed by local research ethics committee at UZ Leuven. No patient-level data were collected. Survey responses were de-identified before analysis and are presented in aggregate form. A list of participating programmes/jurisdictions is reported separately.

### 2.1. Sampling frame

We used the World Health Organization (WHO) TB burden estimates dataset as the jurisdictional sampling frame [16], comprising 215 countries and jurisdictions. We sought programme-level participation from all 215 jurisdictions and additionally included Kosovo and Taiwan, resulting in 217 targeted jurisdictions. In Australia, Canada, and the United States, state-, province-, or territory-level programmes were approached as distinct analytic units because TB programme responsibilities are substantially federated and NTM-related practices and policies were known to vary materially across subnational settings. These programmes were therefore treated as implementation-relevant units rather than being subsumed under a single national observation. Representatives from French overseas departments were also approached.

### 2.2. Respondents

We used a hierarchical recruitment strategy to identify suitable respondents with national-level authority or operational responsibility for TB/NTM/mycobacteria programme implementation, surveillance, diagnostics, or policy development. We prioritised national TB programme (NTP) officials, including NTP directors, deputy directors, programme managers, national surveillance focal points, and TB registry representatives, as well as NTP members and advisors where relevant. When NTP contacts could not be established, we approached senior leadership from the national mycobacteria reference laboratories (NRLs), including directors, heads, managers, or senior laboratory technicians *(only via referral)*. This focus on NTPs and NRLs was deliberate, recognising that many countries operate combined TB-leprosy control programmes and that TB and NTM share substantial overlap in clinical presentation and diagnostic work-up.

If neither NTP nor NRL contacts were available, we contacted senior officials within ministries of health (MoH) or national public health institutes within the MoH (e.g., heads of infectious or communicable disease control units, chief medical officers, chief epidemiologists, or directors of national public health or phthisiology institutes). In a small number of settings where institutional contacts were not reachable or where referrals were provided by NTP/NRL teams, we invited internationally recognised TB/NTM experts based in the respective jurisdictions. This approach enabled broad jurisdictional coverage while prioritising the most relevant available national programme-facing respondent in each setting. Importantly, as the first global mapping exercise of this kind to our knowledge, responses in our study were intended to reflect the perspective of the most relevant available programme-facing national actor in each setting, rather than a formal whole-of-country institutional consensus.

### 2.3. Survey questionnaire

A 25-question survey instrument was developed following a consultative, multi-step process engaging the study authors individually followed by group discussion via email and online meetings. The study authors brought a wide spectrum of expertise in pulmonology, infectious diseases, NTM and TB programme evaluation, mycobacterial diagnostics, global health policy, and statistical and methodological frameworks. The survey was designed to be completed in 10 to 12 minutes and consisted of multiple question formats, including single and multiple-choice items, Likert-type agreement scales, ranking tasks, and array tables to capture variations in policy and systems capacity archetypes.

### 2.4. Survey administration

The survey questionnaire was drafted, hosted, and circulated using a subscription to the LimeSurvey Cloud platform licensed via KU Leuven. This allowed secure, online, multilingual deployment while supporting confidential survey administration through auto-generated respondent identifiers. The survey was developed in English and translated by Google Translate (auto-translate function in LimeSurvey) to Arabic, Chinese (Simplified), French, Russian, and Spanish. French and Russian translations were cross-checked by study authors who are native speakers (NL and LK) while for the remaining languages a back translation was performed using DeepL Translator. Translations were reviewed for conceptual equivalence.

The survey link was disseminated via targeted email invitations to the identified respondents. The contact emails were collected from institutional websites, online published literature, aggregate platforms such as ResearchGate, or from personal connections. Contact information for certain jurisdictions was also obtained from the affiliated WHO Supranational Laboratories (Australia – Adelaide and Brisbane, Belgium, Benin, Chile, and Pakistan) or WHO Country Offices. Connections were also made using social media platforms including Facebook, LinkedIn, and WhatsApp to maximize outreach.

Several spaced reminders, as deemed necessary, were sent over a 3-week period (staged rollout). In the European Union (EU), the survey was also circulated by the European Centre for Disease Prevention and Control (ECDC) to national TB focal points. One response per jurisdiction was requested by default; in selected federated settings, subnational TB programmes were invited as separate programme units.

Because recruitment was adaptive, where an initially identified contact did not respond after reminders, was no longer in post, or could not be reached using available contact information, an alternative eligible contact within the same jurisdiction was approached. Consequently, more than one individual was sometimes invited within a jurisdiction, although only one programme-level response per jurisdiction was retained by default. A total of 502 recruitment-related emails were sent; however, this count included initial invitations, replies, referral requests, and follow-up correspondence and therefore could not be interpreted as the number of unique invitations.

Multiple submissions from the same jurisdiction were reviewed and resolved using a predefined strategy. Where the same individual submitted more than once (n = 3), we retained the most recent submission. Where an initial response was obtained from an expert but a subsequent response from an NTP/NRL contact was received (n = 11), we retained the programme-facing NTP/NRL response for analysis; however, all respondents were offered optional inclusion in the collaborative study group.

### 2.5. Consent for participation

Participation was voluntary. Informed consent was obtained electronically at the beginning of the survey. Respondents were informed that identifying information was collected for respondent verification, follow-up correspondence, and optional inclusion in a collaborative study group, and that survey responses would be de-identified for analysis and reported in aggregate. Respondents were also informed that any verbatim quotations used in publications would be presented only with pseudonymized identifiers. Inclusion in the collaborative study group was optional and was not a condition of survey participation or retention of responses for analysis.

### 2.6. Data analysis

All analyses were conducted at the individual programme level (n = 193). Treating sub-national programmes as separate analytic units allowed us to capture genuine heterogeneity in federated systems, which we considered relevant for future implementation planning and technical assistance. Descriptive analyses were used to summarise respondent characteristics, institutionalisation status, notification status, discussion patterns, and thematic priorities. Categorical variables were presented as counts and percentages. Continuous data, including TB incidence [16] and socio-demographic index 2023 (SDI) [17], were summarised using mean and standard deviation (SD). TB incidence was transformed using log(1+TB incidence) to accommodate zero values and reduce skewness. Summary tables were generated using the *<gtsummary>* package where applicable.

The six WHO regions were used as the primary analytical strata. For additional descriptive granularity, countries and jurisdictions were also classified using a simplified version of the United Nations M49 Standard [18], following the approach used by the United Nations Industrial Development Organization (UNIDO) [19]. Additional stratification was done using UNIDO-defined income and economic groupings, according to the 2025 classification [19]. To reduce identification risk and redundancy across classification systems, we reported results for nine economic groupings that included at least five represented jurisdictions and had limited overlap with the other reported classification schemes.

To examine contextual patterning in institutionalisation and notification statuses, we fitted separate univariable multinomial logistic regression models with log(1+TB incidence) and SDI as predictors using the *<nnet>* package. Global associations were assessed using likelihood-ratio (LR) tests and category-specific effects were summarised as relative risk ratios (RRRs) with 95% confidence intervals (CIs). To aid interpretation, predicted category shares were plotted across the observed ranges of TB incidence and SDI. We prespecified rule-based analytic constructs for institutionalisation and agenda maturity. Institutionalisation classified programme units as NTP-integrated, intent within NTP, outside NTP, or orphaned according to reported current or planned institutional responsibility. Agenda maturity translated reported formal and informal discussion frequency into five stages, ranging from not on agenda/unclear to active agenda. These constructs were intended to translate related programme-reported items into governance-oriented interpretable summaries. Full definitions are provided in the supplementary material.

Associations between agenda stage and categorical variables, including institutional stage, WHO region, and respondent type, were assessed using Chi-squared (χ^2^) tests with Monte Carlo simulation-based P-values. Effect size was summarised using Cramér’s V, followed by examination of standardized residuals to identify cells contributing most strongly to observed patterns (|residual| > 2). For continuous variables, differences were assessed using Kruskal–Wallis test, with effect size summarised using epsilon-squared. Because agenda stage was treated as an ordered construct, monotonic associations were explored using Spearman rank correlation and Jonckheere–Terpstra test using the *<clinfun>* package.

Relative policy momentum was derived from three urgency domains – clinical management, diagnostics, and surveillance – using within-WHO region hierarchical clustering on jurisdiction-level domain scores. Euclidean distance with Ward.D2 linkage was used in the main analysis. Cluster robustness was assessed through sensitivity analyses using alternative linkage strategies and ordinal-aware Gower distance specifications implemented through the *<cluster>* package, as well as bootstrap resampling with Jaccard similarity to evaluate stability of cluster membership. An overall urgency (hereafter “*composite*”) index was constructed for each jurisdiction as the arithmetic mean of the three numeric domain scores, ranging from 0 (not aware) to 4 (strong policy momentum). The effect of categorical and continuous predictors was assessed as described for agenda staging above.

To assess the influence of “not aware” responses, we performed low-end recoding stress tests and complete-case exclusions. We also assessed absolute policy momentum using two prespecified thresholds: (i) a composite urgency score of ≥ 3 and (ii) a stricter criterion requiring all three domain scores and the composite score to be ≥ 3. Agreement between region-relative and absolute global classifications was summarised using cross-tabulation and Cohen’s kappa (κ).

For ranking items, respondents were asked to order predefined barriers and enabling factors from highest to lowest importance. These barriers and enablers were analysed descriptively within institutionalisation and momentum strata. Mean ranks (lower mean ranks indicated greater perceived priority) and their uncertainty were visualised using bootstrapped 95% CIs generated with the *<boot>* package. All derived classifications, clustering analyses, and the resulting governance framework were exploratory and intended to support pattern recognition rather than to represent validated measures or causal relationships. The analyses were conducted in R version 4.5.2 via RStudio. Further methodological details are provided in supplementary material.

## 3. Results

Given our iterative and adaptive recruitment strategy, response coverage was assessed at the jurisdiction and programme-unit levels rather than using the number of unique individuals invited. At the jurisdictional level, responses represented 171 of 217 targeted countries and jurisdictions, corresponding to 78.8% jurisdictional coverage. At the programme-unit level, 193 responses were obtained from 290 invited national and subnational programme units, corresponding to 66.6% programme-unit coverage. The 193 responses included 25 from subnational programmes – seven from Australia, four from Canada, one from a French overseas department, and 13 from the United States (**Supplementary Table S1**). We did not receive any partially completed submissions.

Among the 193 responses, most respondents were programme-facing, with NTPs comprising 64.8% (125/193) of respondents, followed by NRLs 16.1% (31/193). The respondent mix varied by region: NTP respondents predominated in SEARO (100.0%) and AFRO (88.6%), whereas non-NTP respondents contributed a larger share in EURO (**Table 1**).

**Table 1.**
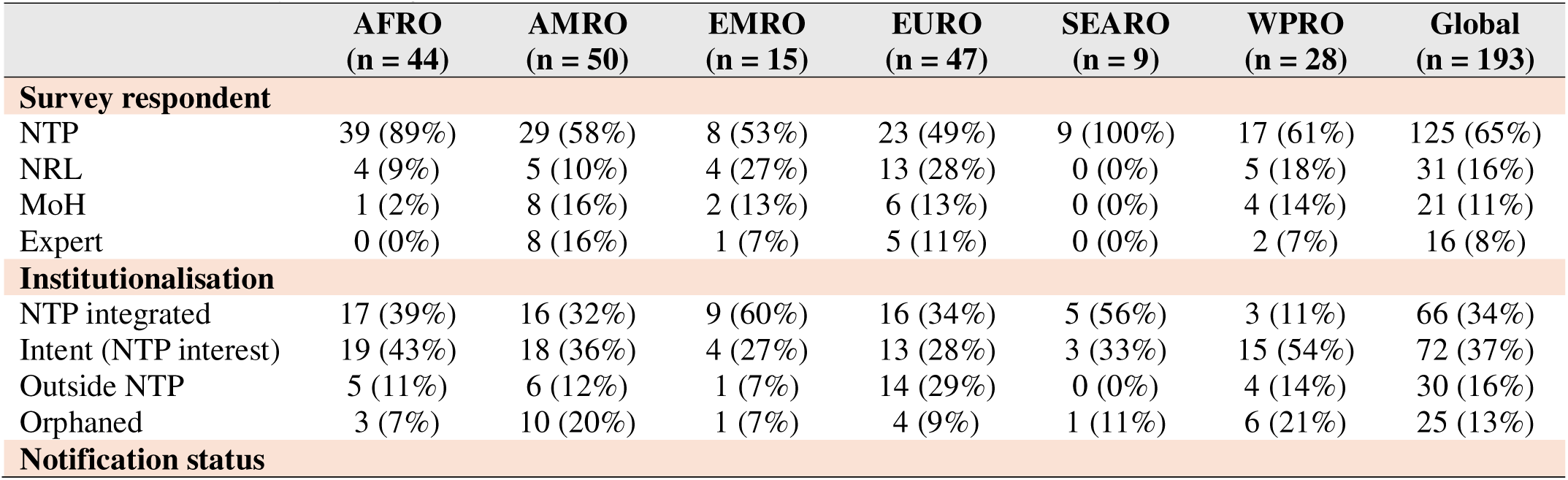
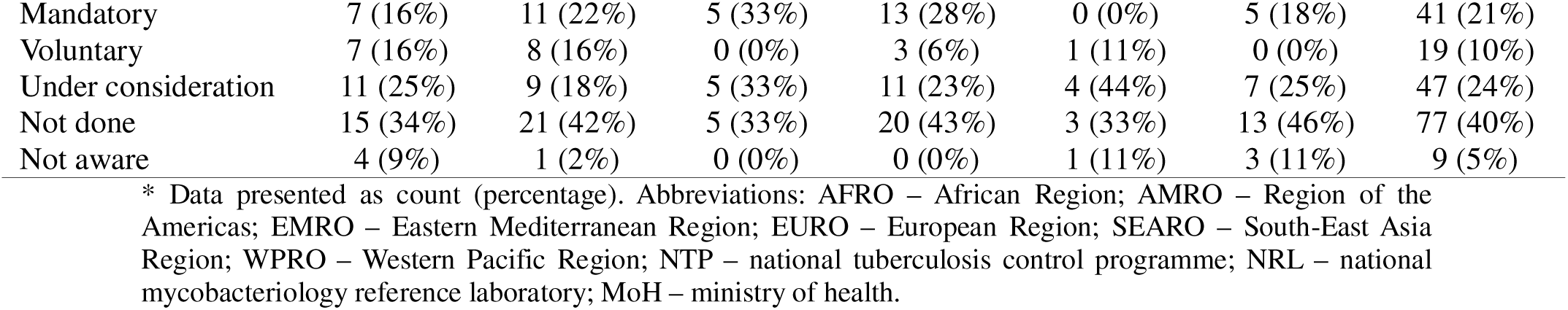
Summary of respondent characteristics, NTM institutionalisation and notification status stratified by WHO region*.

### 3.1. NTM institutionalisation status

Institutionalisation was heterogeneous: 34% of respondents reported that NTM were already integrated within the NTP, 37% indicated interest in institutionalising NTM within the NTP in the next 1-3 years, 16% located NTM outside the NTP, and 13% reported no clear institutional home and no NTP interest in assuming responsibility i.e., orphaned status. Even within these categories, arrangements were often partial or transitional. For example, 36% (24/66) of NTP integrated settings reported pulmonary NTM-only inclusion, while 30% (9/30) of outside-NTP settings reflected planned rather than established placement. Exploratory multinomial models suggested that institutionalisation varied by epidemiologic and development context (global P = 0.018 for TB incidence; P = 0.004 for SDI; **Supplementary Table S2**). Interestingly, higher-TB incidence and lower-SDI settings appeared more likely to show NTP-linked institutionalisation (**Figure 1**).

**Figure 1.**
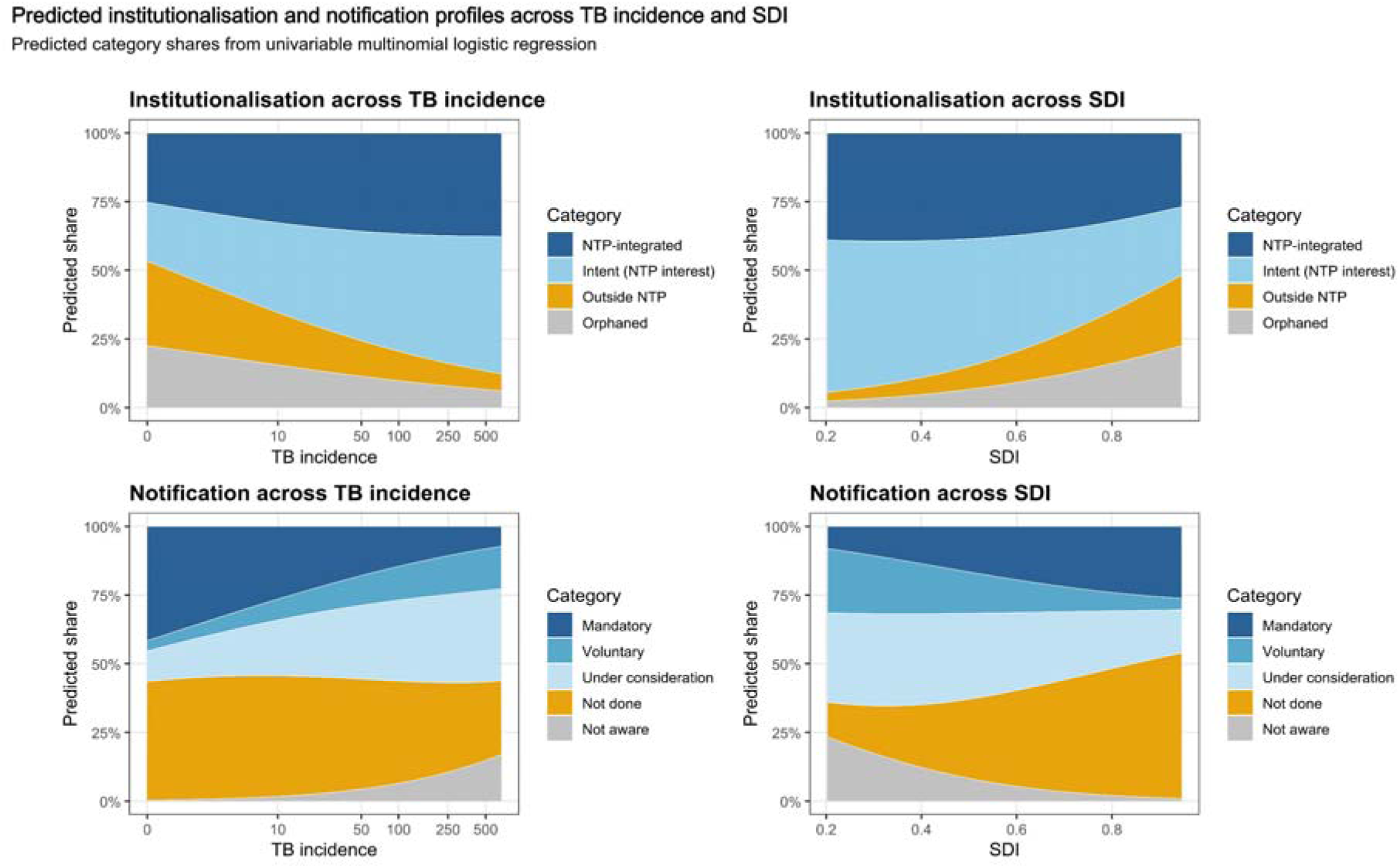
Predicted institutionalisation and notification profiles across TB incidence and SDI. Predicted category shares from univariable multinomial logistic regression models show how institutionalisation and notification status vary across TB incidence and SDI. TB incidence was modelled using log(1+TB incidence) and displayed on the original scale for readability; SDI was modelled per 0.1 increase and displayed on the original SDI scale (0 to 1). These plots illustrate structural gradients in NTM governance architecture rather than causal effects. NTP – national TB programme; SDI – socio-demographic index.

### 3.2. NTM case notification

We assessed surveillance entry points by describing NTM notification status and the pathways through which cases are currently captured and reported. Notification statuses were also observed to be heterogeneous across programmes and regions: 21.2% (41/193) reported mandatory notification, 9.8% (19/193) voluntary notification, and 24.4% (47/193) that notification was under consideration. The remaining 44.6% (86/193) reported that notification was either not being considered in the short term or that respondents were unaware of its status.

Notification appeared most advanced in NTP-integrated settings, where 40.9% (27/66) reported mandatory notification (**Supplementary Table S3**). Notification status varied by both epidemiologic and development context (global P < 0.001 for both TB incidence and SDI; **Supplementary Table S2**). Settings with a high TB incidence appeared less likely to report mandatory notification but at the same time more likely to report voluntary or under-consideration arrangements, whereas higher-SDI settings appeared more likely to show more formalized notification structures (**Figure 1**).

### 3.3. Agenda staging

To operationalize NTM agenda maturity, we classified formal and informal discussion frequency into five agenda stages, ranging from “not on agenda/unclear” to “active agenda.” NTM most often occupied an emerging agenda position (41.5%; 80/193), suggesting that discussion was present but not yet consistently formalized. Latent agenda settings accounted for 20.7% (40/193), whereas both not on agenda/unclear and active agenda settings each represented 18.1% (35/193). Transitional arrangements were rare (1.6%; 3/193).

Agenda stage was significantly associated with institutional stage (Monte Carlo chi-square P < 0.0001; Cramer’s V= 0.27), indicating that policy visibility tended to track with institutional anchoring. This association was driven primarily by over-representation of NTP integrated settings in the emerging agenda group and of orphaned settings in the not on agenda/unclear and latent agenda groups (**Supplementary Figure S1**). Interestingly, there was an over-representation of settings with NTM institutionalised outside the NTP in the active agenda group. Taken together with the finding that 70% of respondents in outside-NTP settings were from NTPs or NRLs, this suggests that TB- and mycobacteriology-facing actors may remain engaged in NTM agenda-setting even where NTM is institutionally located elsewhere.

Further analyses showed that agenda maturity was not associated with WHO region (P = 0.420), respondent type (P = 0.671), TB incidence (Kruskal–Wallis P = 0.455), or SDI (P = 0.445). There was likewise no clear monotonic trend between agenda stage and either log-transformed TB incidence (Spearman rho= -0.10, P = 0.176) or SDI (rho = 0.12, P = 0.102).

### 3.4. Thematic priorities

We then examined how NTM policy discussions were structured across the policy cycle i.e., what programmes talked about as NTM enter (or fail to enter) the agenda. The first clear signal was that where NTM were not on the agenda, discussions were not merely low priority but remained ad hoc and undefined without a specific focus (77.1%). As NTM entered the agenda and discussions matured, the focus showed a clear progression. Clinical management and diagnostics served as the entry points, rising from 17.1% at the not on agenda/unclear stage to 75.0% at the latent stage and remaining high through emerging and active stages (86.2% to 91.4%). In contrast, system-level domains became more prominent only in later stages; funding-related priorities increased from 8.6% to 57.1%, and training/awareness from 11.4% to 77.1% across the same agenda gradient (**Figure 2; Supplementary Table S4**).

**Figure 2.**
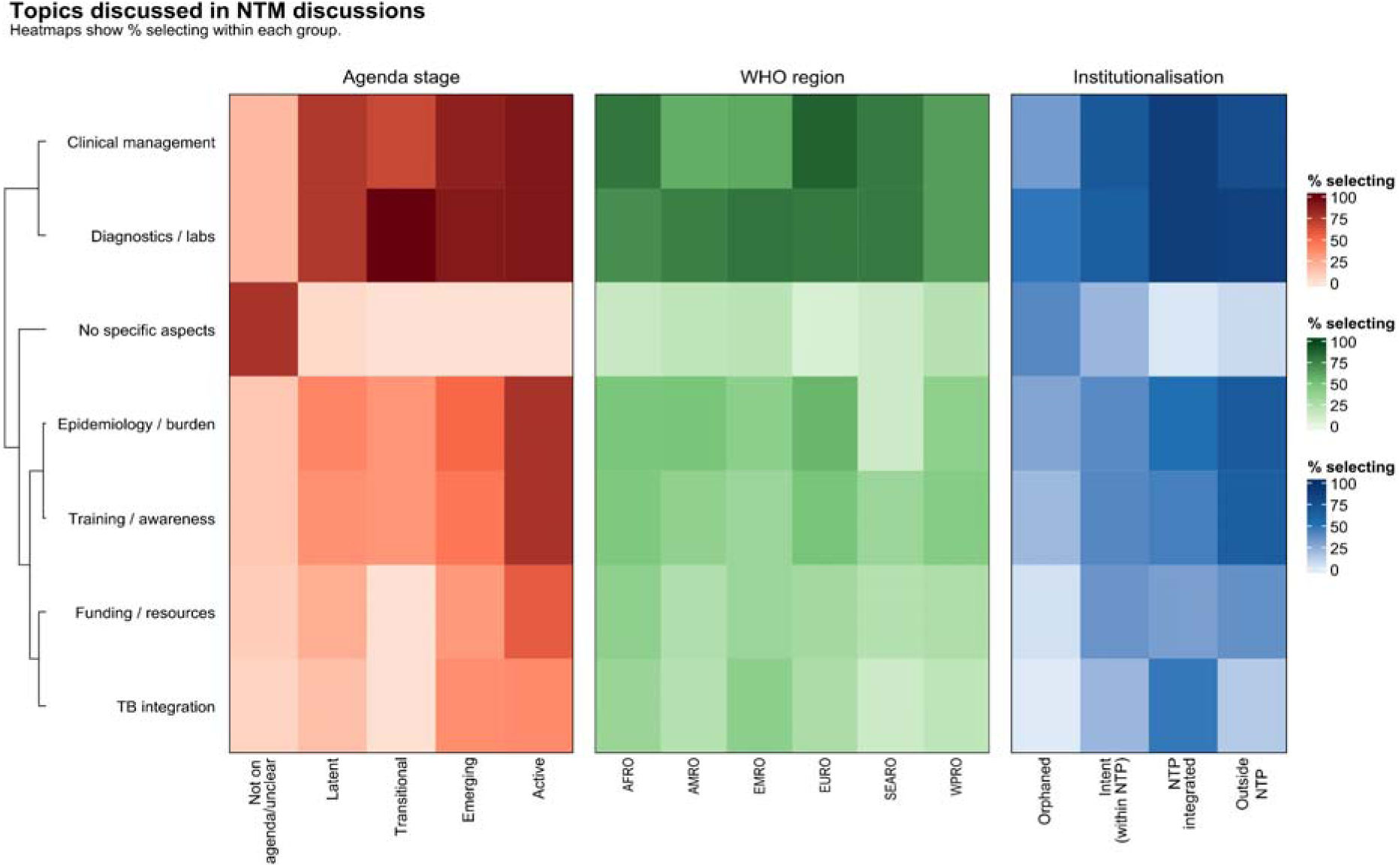
Topics discussed in NTM discussions by agenda stage, WHO region, and institutionalisation. Rows represent discussion topics. Each panel is a heatmap showing, for each topic, the percentage selecting the topic (Yes) within each category of agenda stage (left), WHO region (middle), and institutionalisation (right). Topics were hierarchically clustered (complete linkage; for row ordering only) using Euclidean distance on the concatenated vector of within-category selection percentages across agenda stage, institutionalisation, and WHO region.

Regionally, system-building conversations were unevenly prioritized; most notably, epidemiology, surveillance, and burden estimation was rarely selected in SEARO (11.1%) compared with ∼40–55% in other regions. Topic selection showed no consistent gradient across UNIDO development groups. Differences were generally modest, with funding/resource needs more frequently noted in low-income economies (45.5%; n = 22; **Supplementary Table S5**). Institutionalisation patterns suggested that clear programme ownership was a practical indicator of coordination across the TB-NTM interface. When NTM activity was reported as institutionalised within NTP, discussion focused strongly on implementation domains (clinical and diagnostics each 89.4%) with frequent discussions on TB-NTM integration (47.0%). By contrast, when NTM were institutionalised outside the NTP, respondents more often emphasised talks on cross-cutting needs such as epidemiology and burden estimation (66.7%) and training/awareness (63.3%; **Figure 2**).

### 3.5. NTM pulmonary vs extrapulmonary programmatic visibility

Globally, responses clustered more often in the higher-frequency categories for pulmonary than extrapulmonary NTM, with 53.9% (104/193) of responding programmes reporting pulmonary NTM to be discussed *often* or *all the time*, compared with 30.6% (59/193) for extrapulmonary disease. Conversely, *never* responses were more frequent for extrapulmonary than pulmonary NTM (26.4% vs 14.0%). This difference was observed consistently across institutionalisation types, respondent categories, WHO- and M49-regional groupings, development strata, and economic blocs (**Supplementary Table S6**), indicating a broad visibility gradient in favour of pulmonary NTM.

### 3.6. Relative policy momentum

In all six WHO regions, programmes grouped into two distinct region-relative response clusters for perceived urgency to strengthen national NTM policy – higher (n = 99/193; 51.3%) and lower (n = 94/193; 48.7%). The differences in relative clustering splits were evident from the % share classified as higher momentum across the regions (e.g., AMRO 68% vs SEARO 11%; **Supplementary Table S7 and Supplementary Figure S2**). Cluster assignments were highly consistent across alternative linkage choices under Euclidean distance (ARI often = 1.00), with bootstrap resampling indicating stable profile memberships (B = 500; mean Jaccard range: 0.82 to 0.99). Although agreement was lower under ordinal-aware Gower distance, the substantive separation was structurally and directionally preserved (**Supplementary Tables S8-S11**).

Four key findings emerged from our clustering. First, surveillance was the most significant bottleneck across settings. Urgency to address surveillance (region-relative global mean (SD): 2.79 (0.87) in higher vs 1.29 (0.70) in lower momentum) was rated lowest in both momentum profiles, even in settings where clinical management and diagnostics were perceived as advancing. In fact, addressing policy on diagnostics and clinical management was consistently rated more urgent (region-relative global diagnostics: 3.05 (0.73) in higher vs 1.52 (0.86) in lower momentum; clinical management: 3.04 (0.68) in higher vs 1.43 (0.96) in lower momentum). This cross-domain ordering was preserved across clustering specifications and within regions and sub-regions (**Figure 3; Supplementary Table S7**). Even in higher momentum settings, the mean composite score did not cross the moderate momentum threshold in AMRO and SEARO (mean composite: 2.53 and 2.33, respectively).

**Figure 3.**
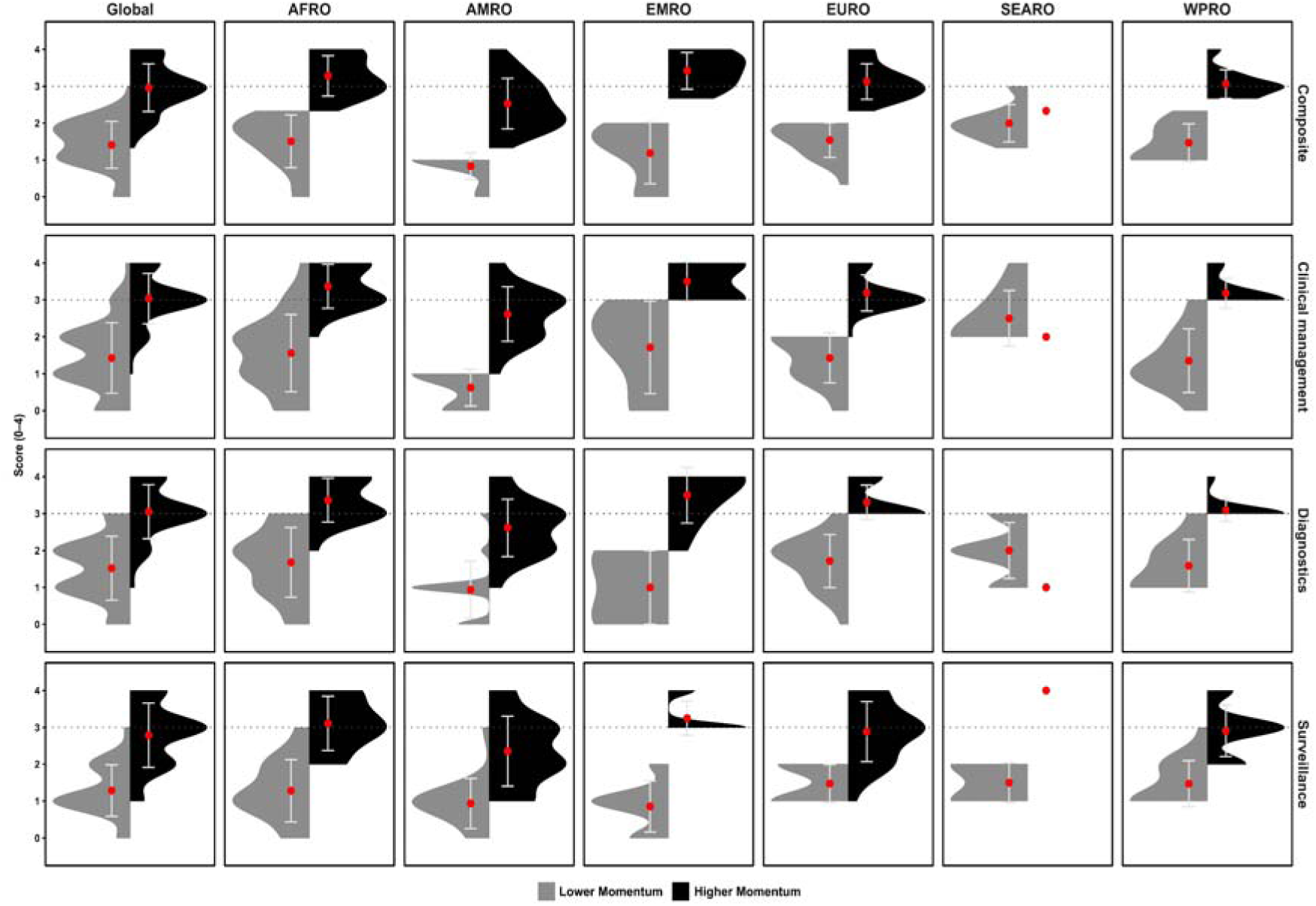
Perceived national policy urgency (“momentum”) for NTM action across composite and its three domains, by WHO region. Split half-violin (bean) plots showing the distribution of programme-level scores (0 to 4) for the composite urgency index (mean of three domains) and for clinical management, diagnostics, and surveillance, stratified by relative momentum groups (“Low momentum” in grey vs “High momentum” in black) defined via within-region hierarchical clustering of domain scores (Euclidean distance; Ward.D2 linkage). Red points indicate the mean and whiskers indicate ± 1 standard deviation (SD) for each group. The horizontal dotted line marks the high-urgency reference absolute threshold (score = 3 or higher). Regions shown are Global, AFRO, AMRO, EMRO, EURO, SEARO, and WPRO; higher values indicate stronger perceived policy urgency/momentum for national/subnational action.

Second, “not aware” responses were concentrated in lower-momentum profiles, suggesting that limited urgency often reflected low salience as well as constrained policy and system capacity. This was particularly pronounced in AMRO and EMRO with high “not aware” proportions across domains. Sensitivity analyses further indicated that how these “not aware” were distributed within a region mattered more than its overall prevalence. Collapsing “not aware” and “absent” into a single category produced near-identical profile assignments in most regions (agreement 96% to 100% in AFRO, AMRO, EMRO, and EURO; **Supplementary Tables S12-S13**). Similarly, excluding jurisdictions with “not aware” in any domain had minimal impact in all regions except AMRO (agreement 50%). This pattern suggests that in AMRO, “not aware” responses were more often domain-specific and concentrated among borderline swing jurisdictions, making them a key axis of within-region separation. In other regions like EMRO “not aware” tended to be confined to clearly lower-momentum settings and did not materially affect profile assignment.

Third, momentum profiles cut across conventional geographic and economic groupings. For example, 48.5% of high-income industrial economies were classified as region-relative lower momentum (composite mean 1.35), alongside non-trivial “not aware” responses (18.2% clinical; 12.1% diagnostics). Lower-momentum shares, based on region-defined classifications, were also substantial among major economic blocks such as ASEAN (55.6%), EU (48.1%), CARICOM (45.5%), BRICS (44.4%), and OECD (44.1%; **Supplementary Table S7**). Finally, association analyses suggested that momentum profiles were not explained by respondent category, TB incidence, or SDI (Fisher-combined P = 0.54, 0.48, and 0.66, respectively; **Supplementary Tables S14-S15**). By contrast, NTM institutionalisation aligned with momentum. Jurisdictions with NTM activities embedded within NTPs were more frequently classified as higher momentum. Within-region associations were strong in AFRO and AMRO and borderline in WPRO, with evidence combined across regions remaining significant (Fisher’s method P = 0.003; **Supplementary Tables S16**).

### 3.7. Absolute Policy Momentum

To gain further exploratory insights, we analysed how an absolute threshold might classify urgency to address NTM policy. We applied two variations of the criteria; (i) a composite i.e., mean of the three domains on the 0–4 scale, to be ≥ 3 rule and (ii) a stricter rule requiring all three individual domains and composite to be ≥ 3. Agreement between the region-relative profiles and absolute classifications was moderate (composite only ≥ 3: 79.3% agreement; Cohen’s κ = 0.589 vs all three domains and composite ≥ 3: 78.8%; Cohen’s κ = 0.579; **Table 2**). Discordance was mostly led by jurisdictions classified as higher momentum relative to regional peers but still below the absolute ≥ 3 benchmark (39 or 41 jurisdictions based on absolute criteria). Interestingly, the reverse discordance pattern was extremely rare (1 or 0 jurisdictions depending on the absolute criteria).

**Table 2.**
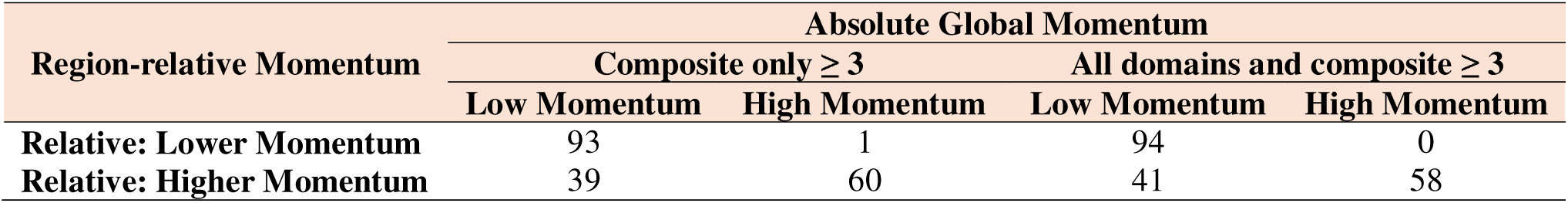
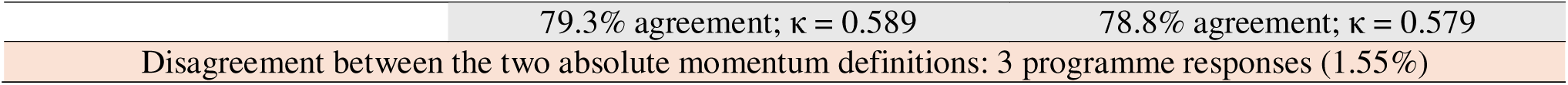
Confusion matrix for relative vs absolute momentum (based on two criteria).

| Region-relative Momentum | Absolute Global Momentum |  |  |  |
| --- | --- | --- | --- | --- |
| | Composite only $\geq 3$ | | All domains and composite $\geq 3$ | |
|  | Low Momentum | High Momentum | Low Momentum | High Momentum |
| Relative: Lower Momentum | 93 | 1 | 94 | 0 |
| Relative: Higher Momentum | 39 | 60 | 41 | 58 |

|  |  |  |
| --- | --- | --- |
| | 79.3% agreement; $\kappa = 0.589$ | 78.8% agreement; $\kappa = 0.579$ |
| Disagreement between the two absolute momentum definitions: 3 programme responses (1.55%) |  |  |

Geographically, discordance was most frequent in AMRO (22/50; 44%), with lower rates elsewhere (e.g., EURO 8/47; 17%). SEARO also showed 2/9 (22%) discordant cases, although estimates were imprecise given the small sample size. Notably, among the discordant “regional leaders” (relatively higher but absolute low), NTM were frequently reported as either already institutionalised within the NTP (46%) or as a willingness to expand within the TB portfolio (23%). Clinical management and diagnostics were typically rated around the moderate range (mean (SD): 2.51 (0.60) and 2.46 (0.64), respectively), whereas surveillance remained lower (mean (SD): 2.00 (0.69)), dragging the composite below the ≥ 3 cutoff. These findings suggest that NTP programme anchoring may enable policy momentum but does not, by itself, ensure absolute cross-domain readiness.

### 3.8. Translating Momentum to Implementation

Having positioned jurisdictions along a spectrum from lower to higher momentum, we next examined what stakeholders saw as the most actionable levers for translating policy urgency into implementable programme action (**Supplementary Figure S3**). The most influential lever was provision of burden/incidence evidence, ranked #1 by 35.6% (16/45) and 52.4% (11/21) of NTP-integrated higher- and lower-momentum settings, respectively (**Supplementary Table S17 and Figure 4**).

**Figure 4.**
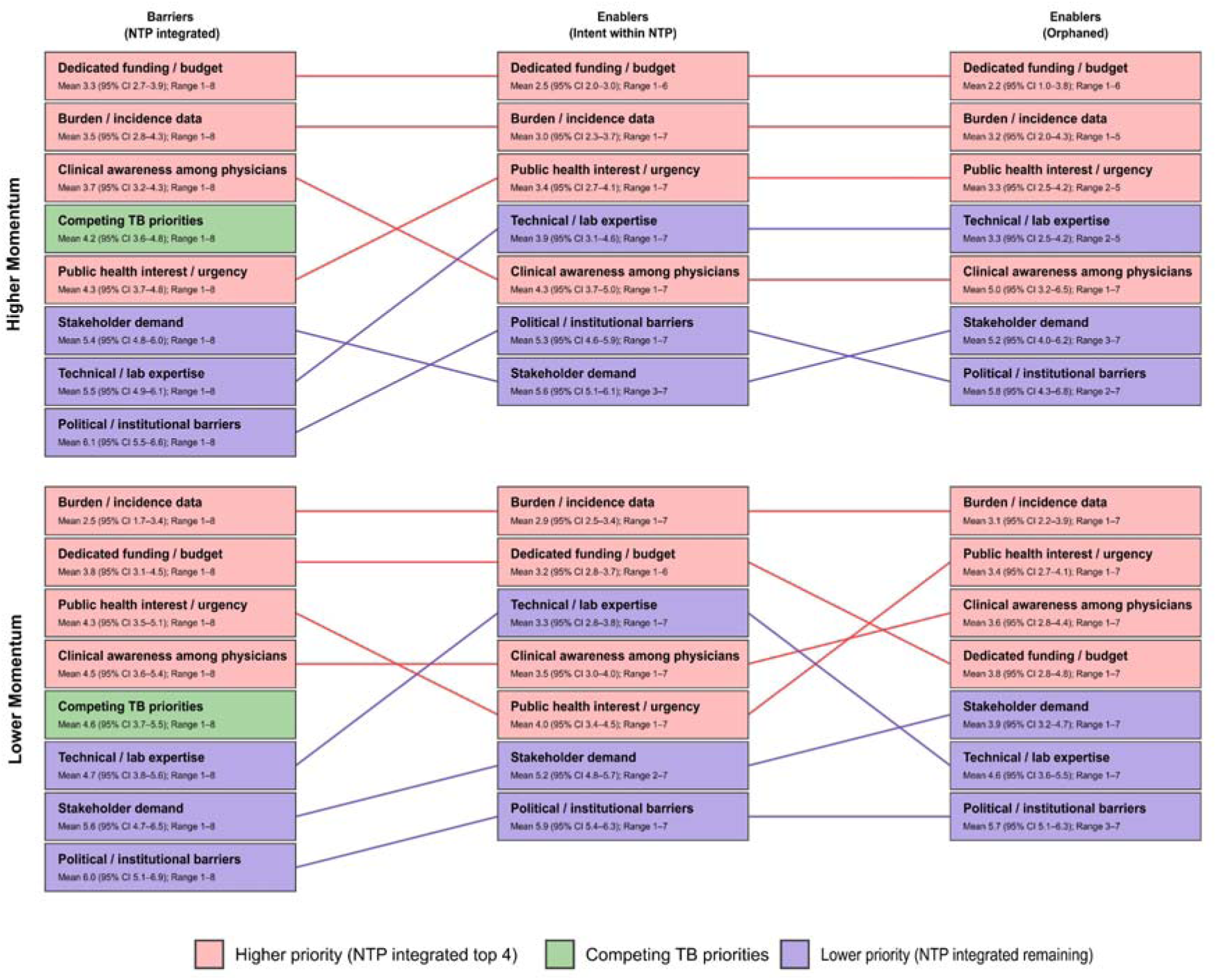
Stakeholder-ranked barriers and enablers for translating NTM discussions and policy urgency into implementable programme action, stratified by region-relative momentum. Within each panel (Higher vs Lower momentum), boxes show the mean rank with bootstrapped 95% CI and observed rank range for each theme. Boxes are further separated by Barriers (NTP-integrated) and Enablers for jurisdictions expressing Intent within NTP or Orphaned institutionalisation. Themes are ordered within each column by mean rank (lower = higher priority). Colours reflect the global NTP integrated ordering: pink = higher-priority themes, green = “Competing TB priorities”, and purple = remaining lower-priority themes. Connecting lines link the same theme across columns and are coloured by the same tier.

Notably, even in the absence of formal notification mandates, in several jurisdictions, structured responses showed that some case-related information was often being captured (**Supplementary Figure S4**). Free-text responses clarified that this occurred typically through ad hoc channels, most commonly via existing TB service platforms (NTP/TB referral centres), NRL-confined laboratory reporting, or clinician notification without programme-level aggregation. Reported constraints included limited capacity for NTM culture and speciation, absence from formal notifiable-disease frameworks, and weak participation from peripheral stakeholders during previous formalization efforts.

This points to a paradoxical surveillance-evidence gap. Supporting this framing, politico-institutional barriers were generally de-prioritised, with over half of NTP-integrated jurisdictions placing them in the bottom two ranks (higher momentum: 55.5%; lower momentum: 52.3%; **Supplementary Table S17**). Lack of stakeholder demand similarly clustered towards the lower ranks (e.g., NTP integrated: 37.7% placed it in the bottom two ranks in higher-momentum settings; intent within NTP: 33.3% placed it last in higher momentum).

The next most important barrier to scaling policy action was availability of dedicated funding. Among NTP-integrated jurisdictions, dedicated funding/budget was ranked in the top three by 66.7% (30/45) and 42.9% (9/21) of higher- and lower-momentum settings, respectively (**Figure 4**). When asked about current funding sources for NTM-related work, over two-thirds of NTP integrated jurisdictions (68%; 45/66) reported no dedicated funding and/or reliance on internal institutional support (*“self-supported”*), underscoring that calls for funding reflect a substantive financing gap rather than a preference for organisational reconfiguration alone. Among the few reporting any funding source, national public-sector research grants were most frequently cited (29% vs 19% in higher vs lower momentum jurisdictions; **Supplementary Figure S5**), suggesting that where resources do exist, they are often confined to research streams rather than sustained programme financing.

While competing TB priorities were frequently acknowledged, they did not dominate as the primary actionable constraint (**Figure 4**). Among jurisdictions with NTM within NTPs, while “competing TB priorities” was ranked in the top three by 35.6% (16/45) of higher-momentum and 42.9% (9/21) of lower-momentum settings, it was rarely ranked #1 (15.6% [7/45] and 4.8% [1/21], respectively). A corollary of this finding was across-the-board high endorsement for both integrating NTM surveillance within NTP channels and supporting a global NTM surveillance effort or registry (**Supplementary Figures S6-S7**), positioning existing NTP/NRL platforms as a pragmatic starting point for early surveillance functions and future global coordination.

## 4. Discussion

In the present study, we assembled 193 programme-level responses, representing 171 countries and jurisdictions, yielding, to our knowledge, one of the broadest programme-level datasets to be reported on NTM policy organization. The wide geographical spread captured perspectives not only from settings where NTM policy and diagnostic capacity are relatively well developed, but also from those where policy attention and system-level entry points are likely to be constrained. The dataset thus provides a rare global snapshot against which future policy development and programme strengthening can be assessed.

A central finding of our work is that institutional ownership of NTM is often weak and, in a small number of settings, completely absent. Fewer than half of jurisdictions reported a clear institutional home for NTM, either within the NTP (34%) or outside it (11%). This weak institutional anchoring may contribute to fragmented arrangements for diagnosis and laboratory oversight, while also limiting the generation of reliable and comparable burden estimates. Prior studies support this interpretation, having drawn attention to uneven surveillance capacities, heterogeneous case definitions, and inconsistent notification practices as barriers to effective scaling of NTM policy and care globally [9,10].

A further indication of the absence of a coherent surveillance function was the variation observed in notification mandates. Overall, 21% (41/193) of respondents reported mandatory case reporting, substantially higher than the few widely cited examples, such as Queensland in Australia and the US CDC Emerging Infections Program (EIP) [20,21]. Among the subset of European settings for which mandatory notification status could be independently verified from online sources, reporting models differed markedly, ranging from clinician-led reporting in Belarus [22], to laboratory-led centralised reporting in Finland [23], to an integrated registry-based system with multi-party inputs in Czechia [24]. Such heterogeneity is also likely to contribute to the surveillance–evidence gap reported by respondents. While financing and the lack of epidemiological data were ranked as the most actionable levers for NTM policy implementation, surveillance remained the weakest policy momentum domain across settings. This creates a familiar reinforcing trap seen in other fields such as paediatric TB and antimicrobial resistance [25,26]. Weak institutional ownership constrains surveillance, while limited surveillance weakens the evidence base needed to secure sustained policy commitment and promote formalization. The same pattern may help explain why NTM appears to enter policy agendas through a staged process, with clinical management and diagnostic concerns gaining recognition earlier, while burden estimation, training, and financing emerge later as systems begin to formalize their response.

Our findings also suggest that this issue is not simply a low-resource or high-burden-country problem. While institutionalisation and notification status appeared to vary across TB epidemiologic and socioeconomic contexts, policy momentum itself did not map neatly onto these gradients. This implies that structural positioning and policy readiness are related but distinct dimensions of NTM governance. Institutional arrangements may arise from the pathways already available within a health system, but momentum depends on whether NTM are recognized as a priority, whether stakeholders have the technical and organisational capacity to respond, and whether the issue can compete successfully for attention, coordination, and financing. Thus, a jurisdiction may have an identifiable institutional home for NTM without strong policy momentum, while another may show growing momentum despite remaining structurally fragmented (**Figure 5**). It is in this context that NTM warrants pragmatic attention, not because of reported increases in detection or burden, but because it is clinically challenging, diagnostically complex, reliant on specialised mycobacterial laboratory capacity, and poorly integrated into routine surveillance pathways.

**Figure 5.**
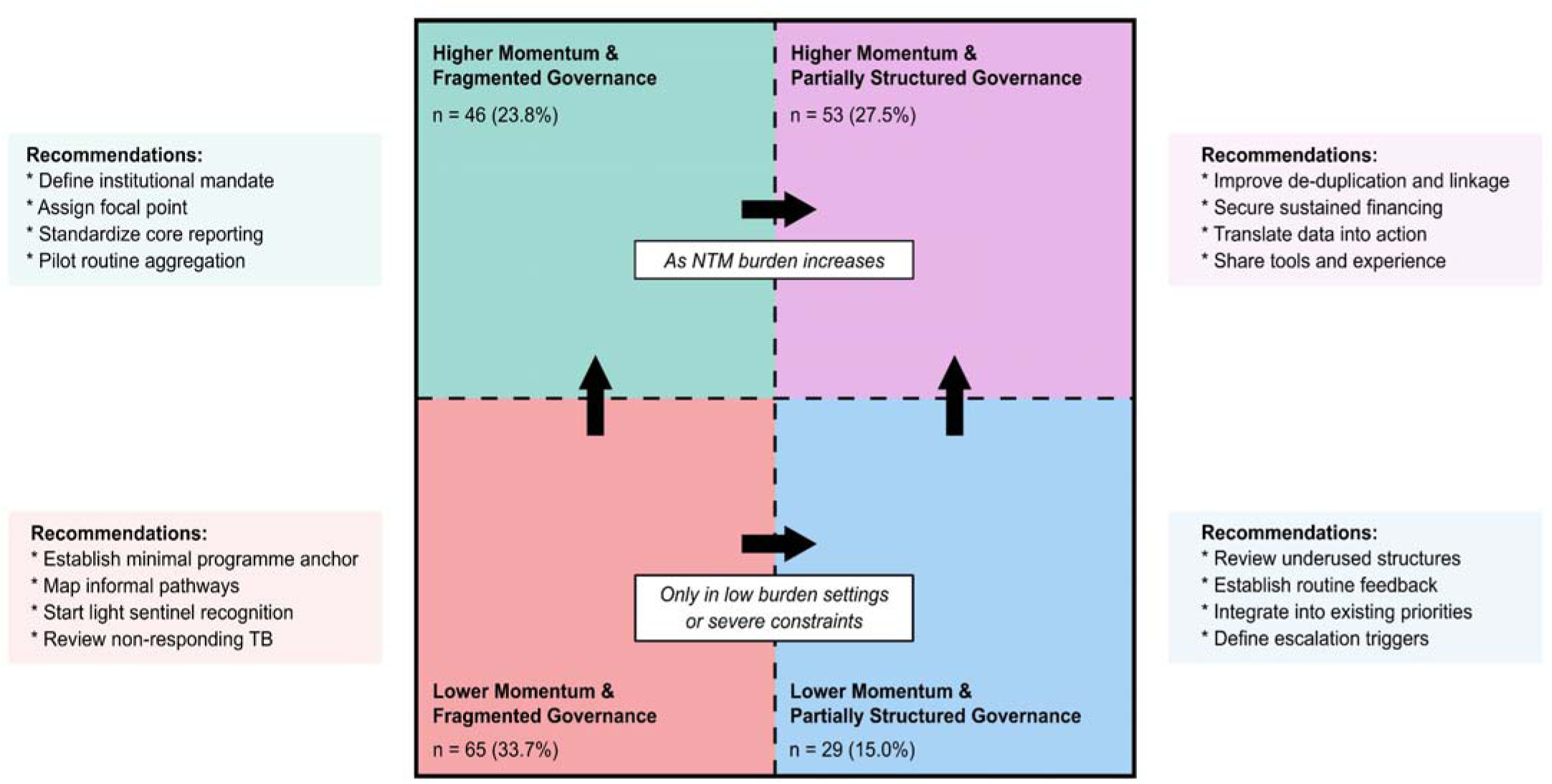
Conceptual positioning of jurisdictions by policy momentum and reported governance structure for NTM, with stage-specific implementation priorities. Jurisdictions were grouped into four categories using a binary cross-classification of policy momentum and governance structure (institutionalisation status and notification status). Counts and percentages indicate the number of jurisdictions falling within each category. Arrows illustrate plausible progression pathways. In most settings, movement towards stronger governance and higher momentum should ideally occur together (upwards + rightwards); however, in low-NTM-burden or severely resource-constrained settings, establishing a minimal governance anchor may need to precede broader momentum-building. Methodology for deriving this binary classification is described in supplementary methods.

This framing also clarifies why TB-facing programmes and mycobacteriology stakeholders were an appropriate entry point for our survey. NTPs and NRLs are among the most globally identifiable and operationally relevant interfaces through which NTM are currently differentiated, discussed, and at least partially recorded. Although NTP respondents predominated overall, non-NTP respondents accounted for a larger share in EURO (51%; 24/47), reflecting settings where TB functions are embedded within broader communicable disease, infection prevention and control, or public health structures. The sampling frame should therefore be understood as a pragmatic reflection of current system organization, rather than a normative claim that NTM inherently belong within the NTPs.

The contextual gradients observed in the exploratory models point in the same direction. In high-TB and low-SDI settings, NTP- or NRL-linked pathways may represent the most available operational home for NTM, whereas in higher-SDI settings responsibility may be distributed across broader public health, respiratory, infectious disease, or specialist service structures, often mirroring existing TB arrangements globally. The stronger emphasis on pulmonary than extrapulmonary NTM discussions in our findings fits this interpretation, suggesting that the most immediate overlap with NTP-linked activities may lie at the pulmonary interface rather than across the full syndromic spectrum. This may also help explain why NTP-linked institutionalisation was associated with higher relative momentum but did not always translate into universal readiness.

A practical reading of our results, therefore, is that in many settings NTP- and NRL-linked systems may provide one of the most plausible early programmatic anchors, or the “*core*”, for NTM-related visibility and coordination, particularly where alternative structures are not yet well defined. Competing TB priorities and politico-legal barriers were not ranked as dominant obstacles, suggesting that NTM are not necessarily perceived as a zero-sum competitor to TB programmes, but as a condition that can be incorporated where governance, reporting pathways, and basic surveillance functions are strengthened. Importantly, this does not imply a single universal starting point. Rather, the route toward stronger NTM governance may differ by setting: as suggested by our exploratory models, some higher-TB contexts may move first through NTP-linked institutionalisation without yet achieving formalized notification, whereas some higher-SDI settings may develop notification more formally even when NTM sits outside NTP structures. Taken together, these findings point toward a possible staged maturity framework for NTM governance (**Figure 6**).

**Figure 6.**
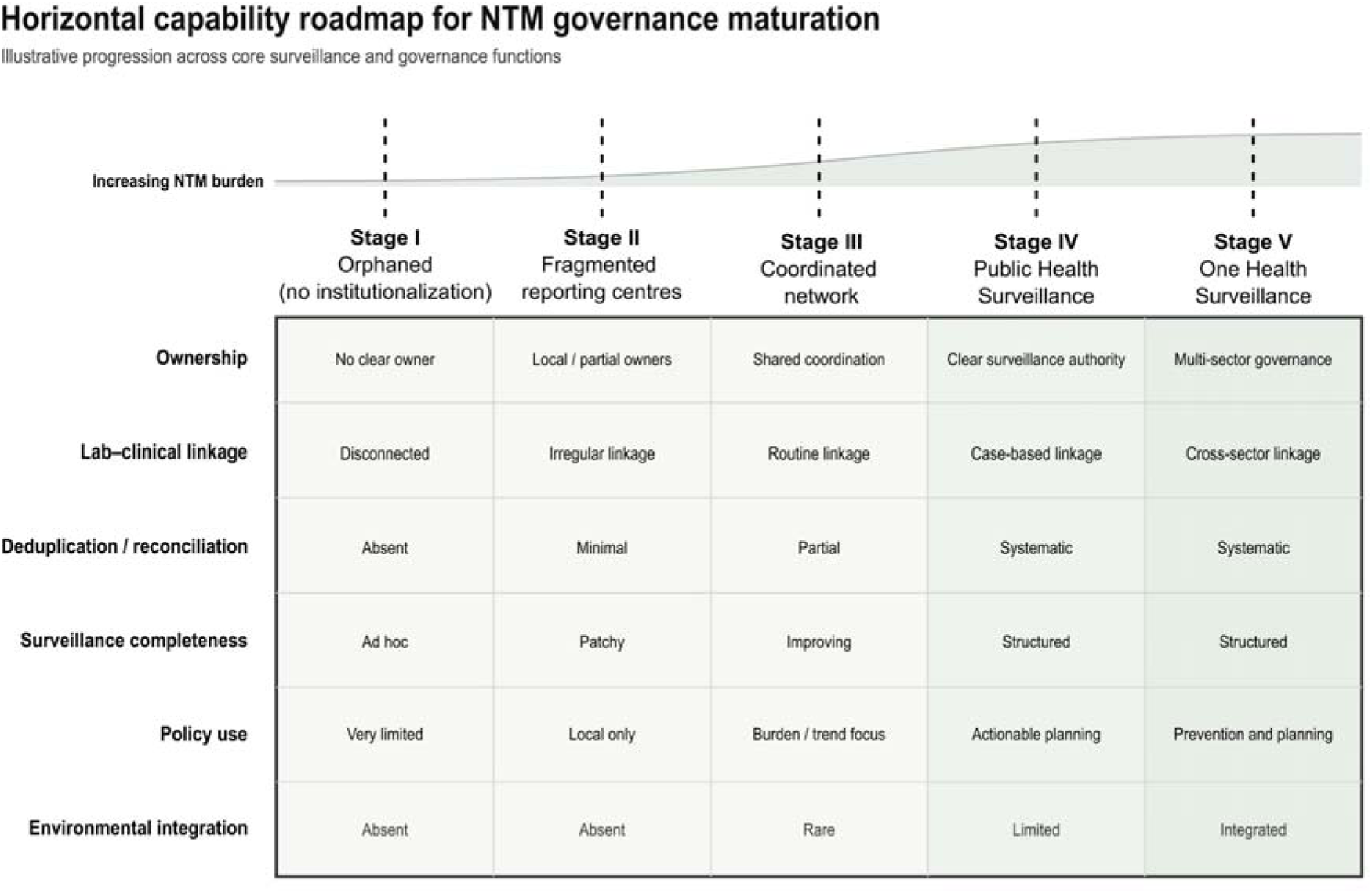
Horizontal capability roadmap for NTM governance maturation. Illustrative framework showing progressive stages in the development of NTM governance and surveillance architecture. Stages range from orphaned settings with no clear institutional owner (Stage I), through fragmented and coordinated reporting arrangements (Stages II–III), to proposed governed public-health surveillance (Stage IV) and, in selected settings, One Health-informed surveillance (Stage V). Across each stage, the figure maps expected progression in six core functions. The framework is intended as a heuristic based on the survey findings and highlights that countries may require different endpoints depending on burden, system capacity, and governance context.

At the lowest stage are orphaned settings, where no clear institutional owner exists. The next stage comprises fragmented reporting arrangements, in which NTM is recognized through local clinical, laboratory, or programme structures, but aggregation remains incomplete and ownership partial. From our discussions with NTPs and NRLs, most countries appeared to fall within this fragmented stage. Fragmented arrangements may exist within or outside NTP/NRL structures. Where they are located within NTP/NRL systems, a hub-and-spoke model may emerge, with reporting led by the NRL from a mycobacteriology perspective. Another possibility is a partial overlap model, in which responsibility is divided by syndrome, with pulmonary NTM retained within the NTP/NRL core and extrapulmonary NTM managed elsewhere. This arrangement may already be reflected in countries where only pulmonary NTM was reported to fall within the NTP ambit (12.4%; 24/193). Where NTM sits entirely outside the NTP, a specialist-led model may apply, with surveillance centred around referral hospitals, tertiary respiratory services, or cystic fibrosis (CF) centres. The prominence of such arrangement in our dataset could also explain why “not aware” responses clustered in low-momentum settings: these may reflect not only low technical capacity, but also low policy visibility, unclear remits, or the absence of an obvious institutional locus from which NTM could be recognized as a programmatic issue.

In any case, we believe that all three models are bound by their own limitations. A stand-alone NRL-centred model may fail to capture sufficient clinical information and may not distinguish total isolates from repeated samples from the same patient. A syndrome-split arrangement may improve operational ownership by allocating pulmonary and extrapulmonary disease to different services but is also likely to perpetuate fragmentation as it does not address the shared need for mycobacterial laboratory linkage. Specialist-led arrangements are likewise unlikely to be sufficient on their own, as expertise may remain concentrated in a small number of centres without creating population-level surveillance ownership.

A possible further stage would be the establishment of a coordinated surveillance network, in which routine linkage between key actors begins to emerge and reporting becomes more systematic, and burden or trend estimation becomes possible, although de-duplication and full case-based reconciliation of cases between different stakeholders may remain incomplete. This intermediate stage may be especially relevant for countries in which some NTM-related activity already exists, but where funding remains ad hoc, internally absorbed, or dependent on research rather than stable programme financing. In such settings, coordination may be the more realistic immediate objective than full institutional consolidation. The next proposed transition would occur at the stage of governed public-health surveillance, where there is clear surveillance authority, systematic reconciliation of cases across data sources, improved completeness, and the capacity to translate surveillance outputs into planning and action. Because NTM exposure is environmental, a One Health-informed model becomes plausible only beyond this stage, and only in selected settings, integrating clinical and laboratory surveillance with health-care-associated and environmental intelligence.

Note that not all countries will need to reach this endpoint. Rather, the appropriate endpoint is likely to depend on local burden, system capacity, and governance context. Lower-burden or lower-capacity settings, for example, may need only light-touch sentinel or NRL-linked surveillance for the foreseeable future, whereas higher-burden or higher-capacity settings may justify broader public-health coordination or, in the longer term, more integrated approaches. This variation in endpoint is consistent with the broad support observed among participants for both TB-linked integration and wider global surveillance development. Respondents appeared more aligned around the need to build surveillance progressively and collectively than around any single universal institutional solution.

From a policy perspective, the practical question is therefore not where NTM should ultimately sit in different health systems, but how existing surveillance-relevant platforms can be used to move jurisdictions from fragmented visibility towards accountable reporting and burden estimation. In this context, NTP/NRL core platforms may, in suitable settings, form part of the technical backbone on which stronger NTM visibility and surveillance functions could be developed. The most feasible near-term step is to enable crosstalk between existing surveillance architectures by standardising a limited set of reporting fields, designating a focal point, improving catchment and aggregation from NRLs and referral centres, and piloting sentinel surveillance through TB and respiratory pathways.

Finally, whatever the eventual endpoint of system maturity, movement beyond orphaned or fragmented arrangements would be necessary for settings seeking to generate credible burden estimates and actionable surveillance information. The value of this study therefore lies not only in documenting system archetypes, but also in making NTM governance more legible for comparative interpretation and context-specific planning. Its immediate policy value is our ability to provide a comparative foundation for national consultation, regional landscape assessment, and consideration of staged visibility or reporting approaches where relevant, with the aim of improving visibility before rising NTM detection becomes unmanaged (*or even mismanaged*) and forces a reactive catch-up.

### 4.1. Limitations

Our study has several limitations. As a cross-sectional survey, it captured reported arrangements at a single point in time and could not determine how long notification mandates, institutional placement, or surveillance arrangements had been in place. Some reported structures may therefore have been newly introduced rather than established practice. Several variables also depended on respondent interpretation. For example, indicating an “intent to include” NTM in the near future may not necessarily imply a formal policy mandate; conversely, some respondents may have reported operational responsibility even where no explicit mandate existed. The questionnaire could not therefore fully distinguish formal authority from de facto practice.

Next, we acknowledge that each response reflected a programme-facing perspective rather than a formal institutional consensus. Nonetheless, the respondent hierarchy was intentionally designed to prioritise actors most likely to contribute to national consultation, technical decision-making, and implementation related to mycobacterial governance within their jurisdictions. At least 138/193 (71.5%) respondents were NTP directors, deputy directors, coordinators or managers, or NRL directors/heads of mycobacteriology. This conservative estimate excludes additional senior ministry, public-health, laboratory, and technical officers among the remaining respondents. The findings therefore represent informed and policy-relevant programme perspectives. Nonetheless, if institutional perspectives diverged within a jurisdiction, this may have influenced the reported classification of institutional ownership, notification status, and perceived policy urgency.

Furthermore, although the multilingual deployment improved inclusiveness, differences in translation or interpretation across languages may have introduced unintended bias. In addition, despite repeated efforts to contact multiple stakeholders across programme structures, some countries and jurisdictions did not respond. Non-response may therefore have led to under- or over-representation of certain governance typologies, institutional arrangements, or levels of surveillance maturity in the final dataset. Also, while treating federated subnational programmes as separate analytic units improved sensitivity to implementation heterogeneity, it meant that these settings contributed multiple programme-level observations; accordingly, the reported estimates were presented at programme-level rather than country-weighted.

Next, some reported barriers are likely to be context-specific and dynamic. For example, competing TB priorities may have appeared less important in settings where NTM burden is currently perceived to be low, but could become more salient as recognition, case detection, or service demand place increasing pressure on resource reallocation. Finally, the maturity framework proposed herein is an interpretive heuristic derived from survey findings rather than a formally validated typology. It is intended to steer and support policy thinking and comparative analysis but will require refinement and validation in future work.

## 5. Conclusions

This study provides a rare global programme-level baseline for understanding how NTM are positioned within health systems. Our findings suggest that NTM should be viewed not only as a clinical or laboratory concern, but also as a health-systems and public-health governance challenge. NTM were reported as sufficiently visible in many settings to raise questions of programmatic ownership, yet reporting and governance arrangements remained weak, heterogeneous, and often fragmented. The immediate policy implication is not that a single institutional home or surveillance model should be imposed across all settings. Rather, existing programme-linked structures may offer pragmatic starting points for improving visibility, coordination, and responsibility where needed. As appropriate endpoints will differ by burden, capacity, and governance context, this framing may help shift the policy conversation from where NTM belongs to how systems can realistically build the functions needed to respond.

## Declarations

### c) Ethical approval

Ethical approval was not required as the study was deemed a minimal-risk, non-interventional survey under local Belgian guidelines as confirmed by local research ethics committee at UZ Leuven. Furthermore, no patient-level data were collected. Data were de-identified and are presented in aggregate such that individual programmes cannot be identified.

### d) Informed Consent

Informed consent to participate and publish the data was taken from all participants as part of the survey questionnaire.

### e) Author contributions

NJ, with input and support from NL, conceptualized the present study and was responsible for methodology. The initial questionnaire was developed by NJ with final draft reviewed and edited by NL, LK, LR, and EA. Translations were verified by NJ, LK, and NL. The survey was designed online by NJ and was circulated to all participants by NJ. Support in establishing contacts was provided by LK, DA, FEAM, AS, SP, LS, RH, LR, EA, and NL. Data collection and curation, investigation, data analysis, and visualizations were done by NJ. NL was responsible for resources, supervision, funding, and project administration. The study group was involved in data collection, data provision, and for the critical reading of the initial draft. Initial manuscript draft was prepared by NJ while all authors were responsible for revising the manuscript. All authors have read the manuscript and agreed to publication.

## f) Acknowledgements

NJ and LK would like to express their sincerest gratitude to colleagues at the WHO Regional Office for Europe (WHO Europe) for their input during the development of the survey. NJ, NL, LK, LG, and LR would also like to thank colleagues at the European Centre for Disease Prevention and Control (ECDC) for their support in the parallel distribution of the survey within EU countries. NJ also extends his gratitude to all those who supported the study by helping in establishing contact with the authorized national representatives. The authors would like to thank all survey respondents who participated in the study but chose not to be listed as collaborative authors. The present study was conducted under the ambit of doctoral (PhD) research work of NJ at KU Leuven.

## g) **Data Availability Statement**

The collected data, in both anonymized and de-anonymized form, cannot be shared without the prior approval and written consent of the respective study participants. Queries regarding data analysis or structured requests for further data insights can be directed to the corresponding author.

