## Supplementary Material for "Global Programmatic Survey on Governance and Surveillance of Nontuberculous Mycobacteria"

**Survey Questionnaire**

***A1. I consent to participate in the present study.** *By agreeing to participate in this survey, you are also providing consent to us for publishing the collective (aggregated) results, as well as any direct quotes you choose to provide. When publishing verbatim responses, we will not identify the country or individual name. Pseudonymized identifiers will be used instead.*

☐ Yes => move to next page
☐ No => end the survey

***B1. Are NTM cases officially notified at a national level? Please also elaborate in the comment box to which authority is the reporting done (name of government or private institution). If under consideration please elaborate on the stakeholders involved e.g., Ministry of Health, University etc.** *Cases = patients with at least 1 positive confirmed NTM isolate.*

☐ Yes, mandatory notifiable condition => elaborate in comment box
☐ Yes, voluntary reporting => elaborate in comment box
☐ No, but under consideration => elaborate in comment box
☐ Reporting not considered in short term => elaborate in comment box
☐ Not aware => elaborate in comment box

**B2. Irrespective of whether NTM burden is monitored nationally, are NTM infections discussed at a national level?**

**B2.1. Formally discussed (presentations, publications, meetings)*
☐ Regularly ☐ Occasionally ☐ Not done at all ☐ Not aware

**B2.2. Informally discussed (group discussions without formal documentation)*
☐ Regularly ☐ Occasionally ☐ Not done at all ☐ Not aware

***B3. In these discussions, which of the following aspects of NTM disease are typically discussed?** *Choose all that are applicable*

☐ Epidemiology, surveillance, and burden estimation
☐ Diagnostic and laboratory-related challenges
☐ Clinical management and treatment pathways
☐ Integration with TB program infrastructure or policy
☐ Funding and resource needs for NTM
☐ Training and awareness among clinicians/laboratory personnel
☐ No specific aspects

**B4. In these discussions, what type of NTM infections are generally discussed?**

**B4.1. Pulmonary infection/disease*

☐ Never ☐ Occasionally/sometimes ☐ Often ☐ All the time

**B4.2. Extrapulmonary infection/disease*

☐ Never ☐ Occasionally/sometimes ☐ Often ☐ All the time

***B5. Are NTM currently under the scope of your current national TB control mission or programme?**

☐ Yes, all forms of NTM are currently under scope => Progress to B6
☐ Yes, only pulmonary NTM are currently under scope => Progress to B6
☐ No, NTM are not under scope => Progress to B8

***B6. As you indicated that NTM are part of your portfolio, what are the current challenges that you face in regards with NTM policy implementation?** *Drag and drop in the order of highest to lowest importance (rank 1 to rank 8).*

☐ Lack of disease burden/incidence data
☐ Competing TB priorities
☐ Lack of laboratory expertise
☐ No dedicated budget for NTM disease management
☐ Political, institutional barriers
☐ Low clinical awareness among physicians
☐ Perceived low public health threat
☐ No stakeholder demand

***B7. What funding sources, if any, currently support work related to NTM (diagnostics, surveillance, research) at national level?** *Choose all that are applicable*

☐ National (private) research grants
☐ National (public) research grants
☐ International research grants
☐ Global donors (e.g., WHO, Global Fund, USAID)
☐ No donors; self-supported by institutions
☐ Others

=> Progress to C1

***B8. You indicated NTM are not part of your national TB program. Is there some other authority nationally who is responsible for monitoring and surveillance of NTM?** *If yes or if there are plans, then in the comment box please indicate the name of such authority.*

☐ Yes, there is another relevant national authority or scientific body => Progress to C1
☐ There are plans on-going to give responsibility to an existing or new institution or authority => Progress to C1
☐ No, there is no relevant national authority or scientific body => Progress to B9

***B9. Since there is currently no national institution or scientific body responsible for NTM, would you be willing to include NTM in your portfolio in the nearest future (next 1-3 years)?**

☐ Yes, would be interested to include NTM
☐ No, not interested to include NTM

***B10. What could support or motivate you in making NTM part of your national TB control mission or programme?** *Drag and drop in the order of highest to lowest importance (rank 1 to rank 7).*

☐ Provision of national disease burden/incidence data
☐ Provision of technical laboratory expertise
☐ Provision of funding/dedicated budget
☐ Removal of political, institutional barriers
☐ Increased clinical awareness among physicians
☐ Increased public health interest and urgency
☐ Clearer stakeholder demand

=> Progress to C1

**C1. How would you describe the perceived urgency of addressing NTM at a national level (among policy stakeholders, Ministry of health etc.)?**

**C1.1. Addressing NTM disease clinical management*
☐ High – there is policy momentum
☐ Moderate – gaining attention
☐ Low – considered marginal
☐ Absent – not on the policy agenda
☐ Not aware

**C1.2. Addressing NTM diagnostics*
☐ High – there is policy momentum
☐ Moderate – gaining attention
☐ Low – considered marginal
☐ Absent – not on the policy agenda
☐ Not aware

**C1.3. Addressing NTM surveillance*

☐ High – there is policy momentum
☐ Moderate – gaining attention
☐ Low – considered marginal
☐ Absent – not on the policy agenda
☐ Not aware

***C2. In the current situation would you, your office, and/or your programme support integrating NTM disease surveillance into TB programmes?**

☐ Yes, full integration
☐ Yes, pilot only
☐ No, TB programme is not suited for this
☐ Not aware

***C3. In the current situation would you or your office support a global surveillance effort or registry for NTM disease cases?**

☐ Yes
☐ Possibly, under pilot or with external support
☐ No

**D1. For the patients with NTM isolates (presumed or confirmed), are the following aspects of the patient's clinical trajectory recorded?**

**D1.1. Treatment initiation status (i.e., whether treatment was started)*
☐ For all patients
☐ For pulmonary NTM cases only
☐ For some patients, randomly selected
☐ For select patients (rare species, severe cases)
☐ For none of the patients
☐ Not aware

**D1.2. Treatment regimen and course (e.g., drugs, dosage, regimen, early discontinuation)*
☐ For all patients
☐ For pulmonary NTM cases only
☐ For some patients, randomly selected
☐ For select patients (rare species, severe cases)
☐ For none of the patients
☐ Not aware

**D1.3. Microbiological outcome (e.g., culture conversion, resistance data)*
☐ For all patients
☐ For pulmonary NTM cases only

☐ For some patients, randomly selected
☐ For select patients (rare species, severe cases)
☐ For none of the patients
☐ Not aware

**D1.4. Clinical or vital outcome (e.g., symptom resolution, patient death, recurrence)*
☐ For all patients
☐ For pulmonary NTM cases only
☐ For some patients, randomly selected
☐ For select patients (rare species, severe cases)
☐ For none of the patients
☐ Not aware

**D1.5. Radiological outcome (e.g., cavitation, consolidations, progression)*
☐ For all patients
☐ For pulmonary NTM cases only
☐ For some patients, randomly selected
☐ For select patients (rare species, severe cases)
☐ For none of the patients
☐ Not aware

***E1. Would you like to be listed as part of collaborative authorship on the resulting publication from the collected data?**
☐ Yes
☐ No

***E2. Your full name**

***E3. Your current designation (position) in regards with the national TB control programme or mission?** *e.g., director, deputy director*

***E4. Country**

***E5. Preferred Email**

** Response is required*

**Thank you for your response.**

### Supplementary Methods

#### S1. Country groupings*

The primary analyses were based on the six WHO regional offices: African Region (AFRO), Region of the Americas (AMRO), Eastern Mediterranean Region (EMRO), European Region (EURO), South-East Asia Region (SEARO), and Western Pacific Region (WPRO). To provide additional geographic granularity, we used the United Nations Standard Country or Area Codes for Statistical Use, more commonly known as the M49 Standard (*Series M, No. 49*). For reporting, the M49 Standard was simplified following the approach used by the United Nations Industrial Development Organization (UNIDO). In this simplified scheme, M49 intermediate and sub-regional levels are combined to yield mutually exclusive, interpretable groups consistent with commonly used global nomenclature (e.g., replacing "Middle Africa" in the M49 Standard with "Central Africa"). We also retained the "other groupings" from the M49 Standard, namely Least Developed Countries (LDCs), Landlocked Developing Countries (LLDCs), and Small Island Developing States (SIDS).

In addition, countries were classified according to UNIDO-defined income groups, based on the World Bank 2025 income classification and structural transformation metrics calculated by UNIDO. UNIDO Statistics (2025 edition) also recognizes 33 economic groupings. For our analysis, we selected a subset of these groupings based on (i) the number of countries in each grouping covered by our dataset (to reduce identification risk for small groups), and (ii) overlap with other classification schemes reported above. We considered only full members, as of December 2025, for each grouping, excluding partner, observer, or associate members. Accordingly, we report aggregated data for:

- Asia-Pacific Economic Cooperation (APEC)
- Association of Southeast Asian Nations (ASEAN)
- BRICS
- Caribbean Community (CARICOM)
- Commonwealth of Independent States (CIS)
- Economic Community of West African States (ECOWAS)
- European Union (EU)
- Group of 20 (G20)
- Organisation for Economic Co-operation and Development (OECD)

* For additional details, kindly refer to the
1. United Nations (UN) M49 Standard (https://unstats.un.org/unsd/methodology/m49/)
2. United Nations Industrial Development Organization Country Classification Edition 2025 (UNIDO; https://stat.unido.org/portal/storage/file/publications/classif/country-classif-report-2025.pdf).

#### S2. Key terminology

Several key analytic constructs were prespecified using rule-based definitions as follows:

##### Institutionalisation

Institutionalisation described whether NTM fell under an established institutional mandate (within the national TB programme [NTP], elsewhere, or not at all), and whether there is stated near-term intent to create such a mandate. Accordingly, there were four possible outcomes – orphaned, intent (within NTP), NTP integrated, and outside NTP.

a) **Orphaned:** NTM are not currently under the scope of the NTP, and the respondent reported no other relevant national authority/scientific body responsible for monitoring/surveillance, along with no expressed interest to include NTM within the NTP portfolio. Orphaned status showed no clear institutional home and no stated near-term pathway to one.

b) **Intent (within NTP):** NTM are not currently under the scope of the NTP, and there is no other national authority responsible, but the respondent indicated explicit interest in including NTM within the NTP portfolio within the next 1 to 3 years. This could also represent ongoing plans for integration.

c) **NTP integrated:** NTM are currently under the scope of the NTP, either covering all forms of NTM or pulmonary NTM only. Within NTP status indicates that institutional responsibility is situated within the NTP's mandate (even if the scope maybe partial or limited to some specific species only).

d) **Outside NTP:** NTM are not currently under the NTP, but the respondent reports that another relevant national authority/scientific body is responsible, or that plans are ongoing to assign responsibility to an existing or new institution outside the NTP.

#### Discussion frequency

Discussion frequency for NTM was considered as an upstream indicator of agenda-setting and institutional attention.

a) **Formal discussions** Structured or scheduled discussions of NTM (e.g., presentations, publications, routine official meetings).

b) **Informal discussions:** Unstructured discussions outside formal formats (e.g., ad hoc conversations, meetings without documentation, informal coordination).

#### Agenda maturity

A rule-based typology derived from two ordinal survey items (formal and informal discussion frequency). The classification imitates inferred agenda maturity consistent with a plausible hypothesis that institutionalisation pathways often require informal attention before formal time allocations.

a) **Active agenda:** Regular formal discussions (scheduled and structured attention).

b) **Emerging agenda:** Either regular informal discussions, *and/or* occasional formal discussions with strong informal signals (i.e., clear momentum).

c) **Transitioning agenda:** Some formal discussions occur but is not yet routinised enough to also show up informally, possibly due to either early institutionalisation, legacy reporting, one-off meetings, or centralised discussions that does not permeate the wider programme.

d) **Latent agenda:** Episodic discussions without reinforcement (occasional informal discussions only, or occasional formal discussions with no informal momentum).

e) **Not on agenda/unclear agenda:** No reported formal or informal discussions, *and/or* an unclear response (i.e., "not sure"), indicating either true absence of agenda attention or limited visibility of NTM discussions within the respondent's remit.

#### Perceived urgency or momentum

We examined perceived national momentum or urgency for NTM policy action across clinical management, diagnostics, and surveillance using two complementary operational definitions.

a) **Relative (profile-based) momentum:** Captures similarity in cross-domain response profiles relative to peers within WHO regions. We operationalised this using within-region hierarchical clustering of the three coded domain responses (Euclidean distance; Ward.D2 linkage), and labelled clusters as higher vs lower relative momentum based on the cluster mean of the composite index.

b) **Absolute (threshold-based) momentum:** Identifies jurisdictions reporting moderate to high momentum across domains. We operationalised this using a binary threshold on the composite index (mean of the three coded domain responses), defining high urgency as mean ≥ 3 on a 0–4 scale (0 = not aware; 4 = high). An alternative definition that we relied on was a binary threshold requiring all individual domains and composite showing high urgency as ≥ 3 on a 0–4 scale.

### S3. Relative momentum clustering

#### Survey items and score coding

Perceived national policy urgency for action on NTM was assessed across three policy domains: (1) clinical management, (2) diagnostics, and (3) surveillance. Each domain used an ordinal response scale with four substantive categories along with "not aware". Responses were mapped onto a 0–4 numeric scale to enable quantitative summarisation: 0 = Not aware; 1 = Absent/not on agenda; 2 = Low/marginal; 3 = Moderate/gaining attention; and 4 = High/strong policy momentum.

#### Composite urgency index

An overall urgency (hereafter "*composite*") index was constructed for each jurisdiction as the arithmetic mean of the three numeric domain scores (clinical management, diagnostics, surveillance). We summarised the three domains using an unweighted composite index (mean of clinical management, diagnostics, and surveillance). Differential weights were not used as the domains were treated as co-primary components of NTM policy readiness, and equal weighting avoided *a priori* imposition of normative assumptions about their relative importance. Domain-specific results are, nonetheless, reported to maintain interpretability.

#### Relative (profile-based) momentum clustering

To identify jurisdictions with similar cross-domain urgency profiles while accounting for regional context, we defined "relative momentum" using within WHO-region hierarchical clustering applied to the three numeric domain scores. For each WHO region, we computed Euclidean distances between jurisdictions using the three domain scores and performed agglomerative hierarchical clustering with Ward's minimum variance criterion (Ward.D2 linkage). Clustering was constrained to two clusters (k = 2) to yield a parsimonious archetype typology for policy interpretation and translation.

Cluster membership was then labelled using the composite urgency index calculated from the original numeric matrix; within each WHO region, the cluster with the higher mean composite score was labelled "higher momentum" and the other "lower momentum".

Importantly, these “higher” and “lower” momentum labels were defined to be region-relative by design, describing position relative to peers within the same WHO region rather than absolute urgency levels intended for direct cross-region comparison. This within-region approach had several advantages. It allows jurisdictions operating within broadly comparable regional policy and health-system contexts to cluster with their peers, improving interpretability of profile differences, and reducing dominance of confounding global structural factors (e.g., income and regional baseline response tendencies) that may drive clustering in pooled analyses. It also aligns with how WHO technical support is commonly organised and coordinated through regional structures.

**S4. Descriptive summaries**

For descriptive reporting, we summarised the composite index and each domain score by geographic and policy-relevant classifiers (e.g., WHO region, M49 standard/UNIDO simplified sub-region groupings, development groupings, and membership in regional/economic blocs), stratified by relative momentum group (region-defined higher vs lower). For each stratum we reported the number of jurisdictions (n); mean and standard deviation (SD); median and interquartile range (IQR) expressed as Q1–Q3; and the proportion of “not aware” responses (score = 0) for each domain. Quartiles were estimated using the Hyndman-Fan type 2 quantile definition, which defines quantiles using the inverted empirical distribution function with averaging at discontinuities. This approach is well-suited to discrete ordinal scales (0–4) with ties, providing quartile estimates that reflect observed response categories rather than heavily interpolated values. We used mean and standard deviation (SD) as the primary summary to retain full scale information and support comparability, while additionally reporting median and interquartile range (IQR) as supplementary ordinal summaries to validate the robustness to distributional assumptions.

**S5. Derivation for binary cross-classification (main manuscript Figure 5)**

To support interpretation of programmatic positioning, we constructed a conceptual two-axis framework combining within-region relative policy momentum and governance structure. Policy momentum was derived from three survey domains which captured reported urgency or readiness across clinical management, diagnostics and surveillance. Responses were recoded to an ordinal 0–4 scale and, consistent with the main analysis, hierarchical clustering was performed within WHO regions to classify jurisdictions into lower- and higher-momentum groups relative to regional peers.

Governance structure was defined using a simple heuristic composite based on institutionalisation and notification status. Institutionalisation responses were assigned ordinal values reflecting the degree of structural anchoring: orphaned settings scored lowest, jurisdictions reporting intent or planning scored intermediate values, and jurisdictions with an identified location either within or outside the NTP scored highest. Notification status was similarly scored from no or unknown notification, through notification under consideration and voluntary notification, to mandatory notification. These two components were summed (range 1 to 3.5 points) and dichotomized into fragmented versus partially structured governance (cutoff 2.5 points). The resulting binary governance classification was then cross-tabulated against the binary momentum classification to generate four conceptual quadrants. Counts and percentages were calculated using all jurisdictions with complete classification on both axes.

**Supplementary Table S1.** Summary of participating countries/jurisdictions in the present study, stratified by WHO region and arranged alphabetically in region (n = 171 jurisdictions; 193 responses including sub-national responses).

| Sr. No. | Country | ISO2 | ISO3 | M49 Region [a] | M49/UNIDO Sub-region [b] | UNIDO Developmental Status [c] | EIE [d] | LDCs [e] | LLDCs [f] | SIDS [g] | TB incidence [h] | SDI [i] | Respondent [j] | Notes |
|---|---|---|---|---|---|---|---|---|---|---|---|---|---|---|
| **WHO AFRO** | | | | | | | | | | | | | | |
| 1 | Algeria | DZ | DZA | Africa | Northern Africa | Middle-income industrializing | | | | | 54 | 0.658 | NTP | |
| 2 | Angola | AO | AGO | Africa | Central Africa | Middle-income industrializing | | Yes | | | 373 | 0.499 | NTP | |
| 3 | Benin | BJ | BEN | Africa | Western Africa | Middle-income industrializing | | Yes | | | 40 | 0.406 | NTP | |
| 4 | Botswana | BW | BWA | Africa | Southern Africa | Middle-income industrializing | | | Yes | | 143 | 0.678 | NTP | |
| 5 | Burkina Faso | BF | BFA | Africa | Western Africa | Low-income | | Yes | Yes | | 57 | 0.307 | NTP | |
| 6 | Cabo Verde | CV | CPV | Africa | Western Africa | Middle-income industrializing | | | | Yes | 47 | 0.562 | NTP | |
| 7 | Cameroon | CM | CMR | Africa | Central Africa | Middle-income industrializing | | | | | 132 | 0.487 | NTP | |
| 8 | Central African Republic | CF | CAF | Africa | Central Africa | Low-income | | Yes | Yes | | 419 | 0.259 | NTP | |
| 9 | Chad | TD | TCD | Africa | Central Africa | Low-income | | Yes | Yes | | 118 | 0.239 | NTP | |
| 10 | Cote d'Ivoire | CI | CIV | Africa | Western Africa | Middle-income industrializing | | | | | 99 | 0.452 | NTP | |
| 11 | Democratic Republic of Congo | CD | COD | Africa | Central Africa | Low-income | | Yes | | | 377 | 0.395 | NTP | |
| 12 | Equatorial Guinea | GQ | GNQ | Africa | Central Africa | Middle-income industrializing | | | | | 199 | 0.646 | NTP | |
| 13 | Eritrea | ER | ERI | Africa | Eastern Africa | Low-income | | Yes | | | 115 | 0.415 | NTP | |
| 14 | Eswatini | SZ | SWZ | Africa | Southern Africa | Middle-income industrial | | | Yes | | 319 | 0.607 | NTP | |
| 15 | Ethiopia | ET | ETH | Africa | Eastern Africa | Low-income | Yes | Yes | Yes | | 141 | 0.394 | NTP | |
| 16 | Gabon | GA | GAB | Africa | Central Africa | Middle-income industrializing | | | | | 371 | 0.631 | NTP | |
| 17 | Gambia | GM | GMB | Africa | Western Africa | Low-income | | Yes | | | 138 | 0.433 | NTP | |
| 18 | Ghana | GH | GHA | Africa | Western Africa | Middle-income industrializing | | | | | 126 | 0.574 | NTP | |
| 19 | Guinea | GN | GIN | Africa | Western Africa | Middle-income industrializing | | Yes | | | 178 | 0.355 | NTP | |
| 20 | Guinea-Bissau | GW | GNB | Africa | Western Africa | Low-income | | Yes | | Yes | 176 | 0.373 | NTP | |
| 21 | Kenya | KE | KEN | Africa | Eastern Africa | Middle-income industrializing | | | | | 207 | 0.549 | NTP | |
| 22 | Lesotho | LS | LSO | Africa | Southern Africa | Middle-income industrializing | | Yes | Yes | | 548 | 0.510 | NTP | |
| 23 | Liberia | LR | LBR | Africa | Western Africa | Low-income | | Yes | | | 207 | 0.383 | NTP | |
| 24 | Madagascar | MG | MDG | Africa | Eastern Africa | Low-income | | Yes | | | 224 | 0.380 | NTP | |
| 25 | Malawi | MW | MWI | Africa | Eastern Africa | Low-income | | Yes | Yes | | 113 | 0.410 | NTP | |
| 26 | Mali | ML | MLI | Africa | Western Africa | Low-income | | Yes | Yes | | 51 | 0.285 | NRL | Multiple responses (3). NTP/NRL retained. |
| 27 | Mauritania | MR | MRT | Africa | Western Africa | Middle-income industrializing | | Yes | | | 65 | 0.518 | NTP | |
| 28 | Mauritius | MU | MUS | Africa | Eastern Africa | Middle-income industrial | | | | Yes | 13 | 0.736 | NTP | |
| 29 | Mozambique | MZ | MOZ | Africa | Eastern Africa | Low-income | | Yes | | | 361 | 0.342 | NTP | |
| 30 | Namibia | NA | NAM | Africa | Southern Africa | Middle-income industrializing | | | | | 422 | 0.620 | NTP | |
| 31 | Niger | NE | NER | Africa | Western Africa | Low-income | | Yes | Yes | | 86 | 0.203 | NTP | |
| 32 | Nigeria | NG | NGA | Africa | Western Africa | Middle-income industrializing | | | | | 219 | 0.512 | NTP | |
| 33 | Republic of Congo | CG | COG | Africa | Central Africa | Middle-income industrializing | | | | | 302 | 0.578 | NTP | |
| 34 | Rwanda | RW | RWA | Africa | Eastern Africa | Low-income | Yes | Yes | Yes | | 62 | 0.462 | NRL | |

| | | | | | | | | | | | | | | |
|---|---|---|---|---|---|---|---|---|---|---|---|---|---|---|
| 35 | Sao Tome and Principe | ST | STP | Africa | Central Africa | Middle-income industrializing | | | | Yes | 87 | 0.523 | NTP | |
| 36 | Senegal | SN | SEN | Africa | Western Africa | Middle-income industrializing | | Yes | | | 132 | 0.413 | NTP | |
| 37 | Seychelles | SC | SYC | Africa | Eastern Africa | High-income industrializing | | | | Yes | 17 | 0.753 | MoH | |
| 38 | Sierra Leone | SL | SLE | Africa | Western Africa | Low-income | | Yes | | | 372 | 0.383 | NRL | |
| 39 | South Africa | ZA | ZAF | Africa | Southern Africa | Middle-income industrial | | | | | 389 | 0.703 | NTP | |
| 40 | Tanzania | TZ | TZA | Africa | Eastern Africa | Middle-income industrializing | Yes | Yes | | | 172 | 0.466 | NTP | |
| 41 | Togo | TG | TGO | Africa | Western Africa | Low-income | | Yes | | | 48 | 0.461 | NTP | |
| 42 | Uganda | UG | UGA | Africa | Eastern Africa | Low-income | | Yes | Yes | | 197 | 0.442 | NTP | |
| 43 | Zambia | ZM | ZMB | Africa | Eastern Africa | Middle-income industrializing | | Yes | Yes | | 272 | 0.506 | NTP | |
| 44 | Zimbabwe | ZW | ZWE | Africa | Eastern Africa | Middle-income industrializing | | | Yes | | 203 | 0.491 | NRL | Multiple responses (2). NTP/NRL retained. |
| **WHO AMRO** | | | | | | | | | | | | | | |
| 45 | Anguilla | AI | AIA | Americas | Caribbean | High-income industrializing | | | | Yes | 0 | 0.880 | MoH | |
| 46 | Argentina | AR | ARG | Americas | South America | Middle-income industrializing | | | | | 44 | 0.765 | NRL | |
| 47 | Aruba | AW | ABW | Americas | Caribbean | High-income industrializing | | | | Yes | 8.3 | 0.908 | NRL | See additional Footnote [5] |
| 48 | Barbados | BB | BRB | Americas | Caribbean | High-income industrializing | | | | Yes | 1 | 0.755 | NTP | |
| 49 | Belize | BZ | BLZ | Americas | Central America | Middle-income industrializing | | | | Yes | 34 | 0.628 | NTP | |
| 50 | Bolivia | BO | BOL | Americas | South America | Middle-income industrializing | | | Yes | | 114 | 0.632 | NTP | Duplicate response. Last response retained. |
| 51 | Brazil | BR | BRA | Americas | South America | Middle-income industrial | | | | | 49 | 0.684 | NTP | |
| 52 | Canada | CA | CAN | | | | | | | | | | | |
| * | Alberta | | | Americas | Northern America | High-income industrial | | | | | 6.2 | 0.887 | Expert | |
| * | British Columbia | | | Americas | Northern America | High-income industrial | | | | | 6.2 | 0.887 | Expert | |
| * | Ontario | | | Americas | Northern America | High-income industrial | | | | | 6.2 | 0.887 | Expert | |
| * | Québec | | | Americas | Northern America | High-income industrial | | | | | 6.2 | 0.887 | Expert | |
| 53 | Chile | CL | CHL | Americas | South America | High-income industrial | | | | | 19 | 0.789 | NTP | Multiple responses (2). NTP/NRL retained. |
| 54 | Colombia | CO | COL | Americas | South America | Middle-income industrializing | | | | | 49 | 0.689 | NTP | |
| 55 | Costa Rica | CR | CRI | Americas | Central America | Middle-income industrial | | | | | 9.3 | 0.715 | MoH | |
| 56 | Cuba | CU | CUB | Americas | Caribbean | Middle-income industrializing | | | | Yes | 11 | 0.695 | MoH | |
| 57 | Dominica | DM | DMA | Americas | Caribbean | Middle-income industrializing | | | | Yes | 4.5 | 0.754 | MoH | |
| 58 | Dominican Republic | DO | DOM | Americas | Caribbean | Middle-income industrial | | | | Yes | 51 | 0.658 | NTP | |
| 59 | Ecuador | EC | ECU | Americas | South America | Middle-income industrializing | | | | | 66 | 0.691 | NTP | |
| 60 | El Salvador | SV | SLV | Americas | Central America | Middle-income industrializing | | | | | 116 | 0.608 | Expert | |
| 61 | Grenada | GD | GRD | Americas | Caribbean | Middle-income industrializing | | | | Yes | 1.7 | 0.678 | MoH | |
| * | Guadeloupe (French Department) | GP | GLP | Americas | Caribbean | High-income industrial | | | | | 7.8 | 0.861 | MoH | See additional Footnote [2] |
| 62 | Guatemala | GT | GTM | Americas | Central America | Middle-income industrializing | | | | | 28 | 0.554 | NTP | |
| 63 | Guyana | GY | GUY | Americas | South America | High-income industrializing | | | | Yes | 67 | 0.696 | MoH | |
| 64 | Haiti | HT | HTI | Americas | Caribbean | Middle-income industrializing | | Yes | | Yes | 175 | 0.464 | NTP | |
| 65 | Honduras | HN | HND | Americas | Central America | Middle-income industrializing | | | | | 38 | 0.520 | NTP | |
| 66 | Jamaica | JM | JAM | Americas | Caribbean | Middle-income industrializing | | | | Yes | 2.3 | 0.692 | NTP | |

| | | | | | | | | | | | | | |
|---|---|---|---|---|---|---|---|---|---|---|---|---|---|
| 67 | Mexico | MX | MEX | Americas | Central America | Middle-income industrial | | | | 28 | 0.693 | NTP | |
| 68 | Panama | PA | PAN | Americas | Central America | High-income industrializing | | | | 63 | 0.715 | NRL | |
| 69 | Paraguay | PY | PRY | Americas | South America | Middle-income industrializing | | Yes | | 81 | 0.674 | NTP | |
| 70 | Peru | PE | PER | Americas | South America | Middle-income industrializing | | | | 139 | 0.686 | NRL | |
| 71 | Puerto Rico | PR | PRI | Americas | Caribbean | High-income industrial | | | Yes | 0.55 | 0.848 | NTP | |
| 72 | Saint Kitts and Nevis | KN | KNA | Americas | Caribbean | High-income industrializing | | | Yes | 1.1 | 0.754 | NTP | |
| 73 | Saint Lucia | LC | LCA | Americas | Caribbean | Middle-income industrializing | | | Yes | 0.56 | 0.703 | MoH | |
| 74 | Suriname | SR | SUR | Americas | South America | Middle-income industrializing | | | Yes | 34 | 0.655 | NTP | |
| 75 | Trinidad and Tobago | TT | TTO | Americas | Caribbean | High-income industrializing | | | Yes | 27 | 0.770 | NTP | |
| 76 | Uruguay | UY | URY | Americas | South America | High-income industrial | | | | 47 | 0.745 | NTP | |
| 77 | United States of America | US | USA | | | | | | | | | | |
| * | California | | | Americas | Northern America | High-income industrial | | | | 3.2 | 0.881 | NTP | |
| * | Colorado | | | Americas | Northern America | High-income industrial | | | | 3.2 | 0.892 | Expert | |
| * | Idaho | | | Americas | Northern America | High-income industrial | | | | 3.2 | 0.856 | NTP | |
| * | Illinois | | | Americas | Northern America | High-income industrial | | | | 3.2 | 0.891 | NTP | |
| * | Massachusetts | | | Americas | Northern America | High-income industrial | | | | 3.2 | 0.914 | NTP | |
| * | Minnesota | | | Americas | Northern America | High-income industrial | | | | 3.2 | 0.896 | Expert | |
| * | Montana | | | Americas | Northern America | High-income industrial | | | | 3.2 | 0.873 | NTP | |
| * | New Hampshire | | | Americas | Northern America | High-income industrial | | | | 3.2 | 0.904 | NRL | |
| * | North Carolina | | | Americas | Northern America | High-income industrial | | | | 3.2 | 0.864 | NTP | |
| * | Oregon | | | Americas | Northern America | High-income industrial | | | | 3.2 | 0.884 | Expert | |
| * | South Dakota | | | Americas | Northern America | High-income industrial | | | | 3.2 | 0.862 | NTP | |
| * | West Virginia | | | Americas | Northern America | High-income industrial | | | | 3.2 | 0.834 | NTP | |
| * | Wisconsin | | | Americas | Northern America | High-income industrial | | | | 3.2 | 0.883 | NTP | |
| 78 | Venezuela | VE | VEN | Americas | South America | Middle-income industrializing | | | | 46 | 0.580 | NTP | Multiple responses (2). NTP/NRL retained. |
| **WHO EMRO** | | | | | | | | | | | | | |
| 79 | Afghanistan | AF | AFG | Asia | Southern Asia | Low-income | Yes | Yes | | 203 | 0.334 | NRL | |
| 80 | Iran | IR | IRN | Asia | Southern Asia | Middle-income industrializing | | | | 10 | 0.718 | NRL | |
| 81 | Iraq | IQ | IRQ | Asia | Western Asia | Middle-income industrializing | | | | 21 | 0.654 | Expert | |
| 82 | Jordan | JO | JOR | Asia | Western Asia | Middle-income industrializing | | | | 3.5 | 0.720 | NTP | |
| 83 | Lebanon | LB | LBN | Asia | Western Asia | Middle-income industrializing | | | | 13 | 0.724 | NTP | |
| 84 | Libya | LY | LBY | Africa | Northern Africa | Middle-income industrializing | | | | 63 | 0.770 | NTP | |
| 85 | Oman | OM | OMN | Asia | Western Asia | High-income industrializing | | | | 11 | 0.781 | NRL | |
| 86 | Pakistan | PK | PAK | Asia | Southern Asia | Middle-income industrializing | | | | 266 | 0.513 | NTP | Duplicate response. Last response retained. |
| 87 | Qatar | QA | QAT | Asia | Western Asia | High-income industrializing | | | | 37 | 0.860 | MoH | |
| 88 | Saudi Arabia | SA | SAU | Asia | Western Asia | High-income industrializing | | | | 8.4 | 0.828 | NTP | |
| 89 | Somalia | SO | SOM | Africa | Eastern Africa | Low-income | Yes | | | 151 | 0.229 | NTP | |
| 90 | Sudan | SD | SDN | Africa | Northern Africa | Low-income | Yes | | | 47 | 0.573 | NTP | |

| | | | | | | | | | | | | |
|---|---|---|---|---|---|---|---|---|---|---|---|---|
| 91 | Tunisia | TN | TN | Africa | Northern Africa | Middle-income industrializing | | | 39 | 0.684 | NRL | |
| 92 | United Arab Emirates | AE | ARE | Asia | Western Asia | High-income industrializing | | | 0.63 | 0.862 | NTP | |
| 93 | Yemen | YE | YEM | Asia | Western Asia | Low-income | Yes | | 40 | 0.446 | MoH | |
| **WHO EURO** | | | | | | | | | | | | |
| 94 | Albania | AL | ALB | Europe | Southern Europe | Middle-income industrializing | | | 15 | 0.707 | NTP | |
| 95 | Armenia | AM | ARM | Asia | Western Asia | Middle-income industrializing | | Yes | 19 | 0.717 | NRL | |
| 96 | Austria | AT | AUT | Europe | Western Europe | High-income industrial | | | 4.9 | 0.854 | NRL | |
| 97 | Azerbaijan | AZ | AZE | Asia | Western Asia | Middle-income industrializing | | Yes | 64 | 0.705 | MoH | |
| 98 | Belarus | BY | BLR | Europe | Eastern Europe | Middle-income industrial | | | 17 | 0.804 | MoH | |
| 99 | Belgium | BE | BEL | Europe | Western Europe | High-income industrial | | | 2.7 | 0.877 | NRL | Multiple responses (2). NTP/NRL retained. |
| 100 | Bulgaria | BG | BGR | Europe | Eastern Europe | High-income industrial | | | 12 | 0.771 | NRL | |
| 101 | Croatia | HR | HRV | Europe | Southern Europe | High-income industrial | | | 6.6 | 0.782 | Expert | |
| 102 | Cyprus | CY | CYP | Asia | Western Asia | High-income industrial | | | 5.5 | 0.856 | NTP | |
| 103 | Czechia | CZ | CZE | Europe | Eastern Europe | High-income industrial | | | 4.9 | 0.831 | NTP | |
| 104 | Denmark | DK | DNK | Europe | Northern Europe | High-income industrial | | | 3.2 | 0.916 | NRL | |
| 105 | Estonia | EE | EST | Europe | Northern Europe | High-income industrial | | | 8.2 | 0.855 | NTP | Multiple responses (2). NTP/NRL retained. |
| 106 | Finland | FI | FIN | Europe | Northern Europe | High-income industrial | | | 3.7 | 0.891 | NTP | |
| 107 | France | FR | FRA | Europe | Western Europe | High-income industrial | | | 7.8 | 0.861 | NRL | |
| 108 | Georgia | GE | GEO | Asia | Western Asia | Middle-income industrializing | | | 45 | 0.755 | NTP | |
| 109 | Germany | DE | DEU | Europe | Western Europe | High-income industrial | | | 5.4 | 0.879 | NRL | |
| 110 | Greece | GR | GRC | Europe | Southern Europe | High-income industrial | | | 3.6 | 0.831 | NTP | Multiple responses (2). NTP/NRL retained. |
| 111 | Hungary | HU | HUN | Europe | Eastern Europe | High-income industrial | | | 5.6 | 0.794 | NRL | |
| 112 | Iceland | IS | ISL | Europe | Northern Europe | High-income industrial | | | 3 | 0.889 | Expert | Duplicate response. Last response retained. |
| 113 | Ireland | IE | IRL | Europe | Northern Europe | High-income industrial | | | 6.2 | 0.907 | Expert | |
| 114 | Israel | IL | ISR | Asia | Western Asia | High-income industrial | | | 2.4 | 0.841 | NTP | |
| 115 | Italy | IT | ITA | Europe | Southern Europe | High-income industrial | | | 5 | 0.834 | NRL | |
| 116 | Kazakhstan | KZ | KAZ | Asia | Central Asia | Middle-income industrializing | | Yes | 45 | 0.727 | MoH | |
| 117 | Kosovo | XK | XKX | Europe | Southern Europe | Middle-income industrializing | | | 33 | 0.785 | NTP | See additional Footnotes [3] and [7] |
| 118 | Kyrgyzstan | KG | KGZ | Asia | Central Asia | Middle-income industrializing | | Yes | 128 | 0.625 | NTP | |
| 119 | Latvia | LV | LVA | Europe | Northern Europe | High-income industrial | | | 21 | 0.851 | NRL | Multiple responses (2). NTP/NRL retained. |
| 120 | Lithuania | LT | LTU | Europe | Northern Europe | High-income industrial | | | 31 | 0.856 | NRL | |
| 121 | Luxembourg | LU | LUX | Europe | Western Europe | High-income industrial | | | 7.5 | 0.895 | MoH | |
| 122 | Malta | MT | MLT | Europe | Southern Europe | High-income industrial | | | 15 | 0.830 | NTP | |
| 123 | Moldova | MD | MDA | Europe | Eastern Europe | Middle-income industrializing | | Yes | 64 | 0.734 | NTP | |
| 124 | Montenegro | ME | MNE | Europe | Southern Europe | Middle-income industrializing | | | 16 | 0.789 | NRL | |
| 125 | Netherlands | NL | NLD | Europe | Western Europe | High-income industrial | | | 4.7 | 0.908 | NRL | |
| 126 | North Macedonia | MK | MKD | Europe | Southern Europe | Middle-income industrial | | Yes | 11 | 0.758 | NTP | |
| 127 | Norway | NO | NOR | Europe | Northern Europe | High-income industrial | | | 3.3 | 0.917 | MoH | |

| | | | | | | | | | | | | | | |
|---|---|---|---|---|---|---|---|---|---|---|---|---|---|---|
| 128 | Poland | PL | POL | Europe | Eastern Europe | High-income industrial | | | | | 11 | 0.847 | MoH | |
| 129 | Portugal | PT | PRT | Europe | Southern Europe | High-income industrial | | | | | 15 | 0.786 | NTP | |
| 130 | Romania | RO | ROU | Europe | Eastern Europe | High-income industrial | | | | | 60 | 0.769 | NTP | |
| 131 | Serbia | RS | SRB | Europe | Southern Europe | Middle-income industrial | | | | | 8.7 | 0.785 | NTP | |
| 132 | Slovakia | SK | SVK | Europe | Eastern Europe | High-income industrial | | | | | 3.3 | 0.813 | NTP | |
| 133 | Slovenia | SI | SVN | Europe | Southern Europe | High-income industrial | | | | | 2.6 | 0.845 | NTP | |
| 134 | Spain | ES | ESP | Europe | Southern Europe | High-income industrial | | | | | 7.9 | 0.812 | NTP | |
| 135 | Sweden | SE | SWE | Europe | Northern Europe | High-income industrial | | | | | 3.3 | 0.911 | NTP | |
| 136 | Switzerland | CH | CHE | Europe | Western Europe | High-income industrial | | | | | 5.4 | 0.946 | Expert | |
| 137 | Türkiye | TR | TUR | Asia | Western Asia | Middle-income industrial | | | | | 13 | 0.731 | Expert | |
| 138 | Ukraine | UA | UKR | Europe | Eastern Europe | Middle-income industrializing | | | | | 86 | 0.765 | NTP | |
| 139 | United Kingdom | GB | GBR | Europe | Northern Europe | High-income industrial | | | | | 9.7 | 0.880 | NTP | |
| 140 | Uzbekistan | UZ | UZB | Asia | Central Asia | Middle-income industrializing | | | Yes | | 51 | 0.632 | NTP | |
| **WHO SEARO** | | | | | | | | | | | | | | |
| 141 | Bangladesh | BD | BGD | Asia | Southern Asia | Middle-income industrializing | Yes | Yes | | | 221 | 0.517 | NTP | |
| 142 | Bhutan | BT | BTN | Asia | Southern Asia | Middle-income industrializing | | | Yes | | 191 | 0.524 | NTP | |
| 143 | India | IN | IND | Asia | Southern Asia | Middle-income industrializing | Yes | | | | 187 | 0.607 | NTP | |
| 144 | Maldives | MV | MDV | Asia | Southern Asia | Middle-income industrializing | | | | Yes | 40 | 0.661 | NTP | |
| 145 | Myanmar | MM | MMR | Asia | South-eastern Asia | Middle-income industrializing | Yes | Yes | | | 482 | 0.532 | NTP | |
| 146 | Nepal | NP | NPL | Asia | Southern Asia | Middle-income industrializing | | Yes | Yes | | 227 | 0.471 | NTP | |
| 147 | Sri Lanka | LK | LKA | Asia | Southern Asia | Middle-income industrializing | | | | | 59 | 0.731 | NTP | |
| 148 | Thailand | TH | THA | Asia | South-eastern Asia | Middle-income industrial | | | | | 146 | 0.687 | NTP | Multiple responses (2). NTP/NRL retained. |
| 149 | Timor-Leste | TL | TLS | Asia | South-eastern Asia | Middle-income industrializing | | Yes | | Yes | 496 | 0.485 | NTP | |
| **WHO WPRO** | | | | | | | | | | | | | | |
| 150 | Australia | AU | AUS | | | | | | | | | | | |
| * | Australia Capital Territory | | | Oceania | Australia and New Zealand | High-income industrial | | | | | 6.9 | 0.853 | NTP | |
| * | Northern Territory | | | Oceania | Australia and New Zealand | High-income industrial | | | | | 6.9 | 0.853 | NTP | |
| * | Queensland | | | Oceania | Australia and New Zealand | High-income industrial | | | | | 6.9 | 0.853 | NTP | |
| * | South Australia | | | Oceania | Australia and New Zealand | High-income industrial | | | | | 6.9 | 0.853 | NRL | |
| * | Tasmania, Australia | | | Oceania | Australia and New Zealand | High-income industrial | | | | | 6.9 | 0.853 | NTP | |
| * | Victoria | | | Oceania | Australia and New Zealand | High-income industrial | | | | | 6.9 | 0.853 | NTP | |
| * | Western Australia | | | Oceania | Australia and New Zealand | High-income industrial | | | | | 6.9 | 0.853 | NTP | |
| 151 | Brunei Darussalam | BN | BRN | Asia | South-eastern Asia | High-income industrializing | | | | | 65 | 0.832 | NRL | |
| 152 | China | CN | CHN | Asia | Eastern Asia | Middle-income industrial | Yes | | | | 49 | 0.728 | NTP | |
| 153 | Fiji | FJ | FJI | Oceania | Melanesia | Middle-income industrializing | | | | Yes | 104 | 0.686 | NTP | |
| 154 | Guam | GU | GUM | Oceania | Micronesia | High-income industrial | | | | Yes | 55 | 0.812 | NTP | See additional Footnote [1] |
| 155 | Hong Kong SAR | HK | HKG | Asia | Eastern Asia | High-income industrializing | | | | | 66 | 0.849 | NTP | |
| 156 | Indonesia | ID | IDN | Asia | South-eastern Asia | Middle-income industrializing | Yes | | | | 382 | 0.666 | NRL | |

| | | | | | | | | | | | | | | |
|---|---|---|---|---|---|---|---|---|---|---|---|---|---|---|
| 157 | Japan | JP | JPN | Asia | Eastern Asia | High-income industrial | | | | | 9.8 | 0.876 | Expert | |
| 158 | Malaysia | MY | MYS | Asia | South-eastern Asia | Middle-income industrial | Yes | | | | 96 | 0.770 | NTP | Multiple responses (2). NTP/NRL retained. |
| 159 | Marshall Islands | MH | MHL | Oceania | Micronesia | Middle-income industrializing | | | | Yes | 411 | 0.605 | MoH | |
| 160 | Mongolia | MN | MNG | Asia | Eastern Asia | Middle-income industrializing | | | Yes | | 446 | 0.660 | NRL | |
| 161 | Nauru | NR | NRU | Oceania | Micronesia | High-income industrial | | | | Yes | 152 | 0.626 | NTP | |
| 162 | New Zealand | NZ | NZL | Oceania | Australia and New Zealand | High-income industrial | | | | | 7.9 | 0.866 | NRL | |
| 163 | Northern Mariana Islands | MP | MNP | Oceania | Micronesia | High-income industrial | | | | Yes | 35 | 0.791 | MoH | See additional Footnote [1] |
| 164 | Palau | PW | PLW | Oceania | Micronesia | High-income industrializing | | | | Yes | 28 | 0.756 | MoH | |
| 165 | Papua New Guinea | PG | PNG | Oceania | Melanesia | Middle-income industrializing | | | | Yes | 664 | 0.437 | NTP | |
| 166 | Philippines | PH | PHL | Asia | South-eastern Asia | Middle-income industrializing | | | | | 625 | 0.669 | NTP | |
| 167 | Singapore | SG | SGP | Asia | South-eastern Asia | High-income industrial | | | | Yes | 43 | 0.870 | NTP | |
| 168 | South Korea | KR | KOR | Asia | Eastern Asia | High-income industrial | | | | | 35 | 0.892 | Expert | |
| 169 | Taiwan | TW | TWN | Asia | Eastern Asia | High-income industrial | | | | | 28 | 0.882 | MoH | See additional Footnote [4] |
| 170 | Vanuatu | VU | VUT | Oceania | Melanesia | Middle-income industrializing | | | | Yes | 44 | 0.477 | NTP | |
| 171 | Viet Nam | VN | VNM | Asia | South-eastern Asia | Middle-income industrial | Yes | | | | 182 | 0.643 | NTP | |

[*] Sub-national jurisdictions that were not accounted for when calculating the overall response rate (n = 171). These sub-national jurisdictions were accounted as separate independent units in all other analysis (n = 193).
[a] Region of the jurisdiction as defined by the United Nations (UN) M49 Standard (available at https://unstats.un.org/unsd/methodology/m49/).
[b] Simplified sub-regional groupings based on the M49 standard as defined by the United Nations Industrial Development Organization Country Classification Edition 2025 (UNIDO; available at https://stat.unido.org/portal/storage/file/publications/classif/country-classif-report-2025.pdf).
[c] Stage of industrial development grouping as defined by the United Nations Industrial Development Organization Country Classification Edition 2025 (UNIDO; available at https://stat.unido.org/portal/storage/file/publications/classif/country-classif-report-2025.pdf).
[d] Emerging Industrial Economies (EIE) as defined by the United Nations Industrial Development Organization Country Classification Edition 2025 (UNIDO; available at https://stat.unido.org/portal/storage/file/publications/classif/country-classif-report-2025.pdf).
[e] Least Developed Countries (LDCs) as defined by the United Nations (UN) M49 Standard (available at https://unstats.un.org/unsd/methodology/m49/).
[f] Land Locked Developing Countries (LLDCs) as defined by the United Nations (UN) M49 Standard (available at https://unstats.un.org/unsd/methodology/m49/).
[g] Small Island Developing States (SIDS) as defined by the United Nations (UN) M49 Standard (available at https://unstats.un.org/unsd/methodology/m49/).
[h] Estimated incidence (all forms) of tuberculosis (TB) per 100 000 population as provided by the World Health Organization (WHO) Global Tuberculosis Report 2025 (available at https://www.who.int/teams/global-tuberculosis-programme/data#app).
[i] Socio-Demographic Index (SDI) as reported by the Global Burden of Disease (GBD) 2023 estimates, Institute for Health Metrics and Evaluation (IHME), University of Washington, US (available at https://ghdx.healthdata.org/record/gbd-2023-socio-demographic-index-sdi).
[j] Respondent type was determined and deduced based on the affiliation and current role in National TB Program (as reported by the respondent), matched with available online information before sending participation invitation. NTP – National TB Control Program; NRL – National TB/Mycobacteria Reference Laboratory; MoH – Ministry of Health. There could be slight variations in interpretation and assignment.

[1] For Guam and the Northern Mariana Islands, UN M49/UNIDO classifications are not available. They were classified as Micronesia based on geography; United States (US) developmental status was applied.
[2] For Guadeloupe, UN M49/UNIDO classifications and SDI are not available. It was classified as Caribbean based on geography; French (France) developmental status and SDI was applied.
[3] TB incidence for Kosovo was taken from Minias A et al., First insights into the genetic composition of *M. tuberculosis* population circulating in Kosovo. J Clin Tuberc Other Mycobact Dis. 2025; 40:100537. doi: 10.1016/j.jctube.2025.100537.
[4] TB incidence for Taiwan was taken from Chan PC et al., Assessing the impact of the TB response in Taiwan - the journey towards ending TB. IJTLD Open. 2025; 2(5):251-259. doi: 10.5588/ijtldopen.25.0103.
[5] For Aruba, SDI was not available. It was assigned Dutch (The Netherlands) SDI given its status as a constituent island country within the Kingdom of the Netherlands.
[6] For Anguilla, SDI was not available. It was assigned British (United Kingdom) SDI given its status as a British Overseas Territory.
[7] For Kosovo, SDI was not available. SDI of Serbia was assigned.

**Supplementary Table S2.** Associations of NTM institutionalisation and notification status with TB incidence and SDI from univariable multinomial logistic regression models.

| | log(1+TB incidence) | | SDI | |
|---|---|---|---|---|
| | **RRR (95% CI)** | **P value** | **RRR (95% CI)** | **P value** |
| **Institutionalisation** | | | | |
| NTP integrated | Ref | Ref | Ref | Ref |
| Intent (NTP interest) | 1.07 (0.86 to 1.33) | 0.529 | 0.94 (0.78 to 1.14) | 0.543 |
| Outside NTP | 0.73 (0.55 to 0.98) | 0.037 | 1.39 (1.04 to 1.86) | 0.028 |
| Orphaned | 0.77 (0.57 to 1.04) | 0.090 | 1.43 (1.04 to 1.97) | 0.030 |
| | Global LR = 10.06, df = 3, P = 0.018 | | Global LR = 13.54, df = 3, P = 0.004 | |
| **Notification status** | | | | |
| Mandatory | Ref | Ref | Ref | Ref |
| Voluntary | 1.63 (1.12 to 2.36) | 0.010 | 0.68 (0.49 to 0.93) | 0.015 |
| Under consideration | 1.55 (1.16 to 2.06) | 0.003 | 0.77 (0.60 to 1.00) | 0.048 |
| Not done | 1.22 (0.94 to 1.57) | 0.134 | 1.03 (0.81 to 1.32) | 0.788 |
| Not aware | 2.40 (1.36 to 4.24) | 0.002 | 0.55 (0.37 to 0.83) | 0.005 |
| | Global LR = 19.08, df = 4, P < 0.001 | | Global LR = 19.96, df = 4, P < 0.001 | |

* Relative risk ratios (RRRs) with 95% confidence intervals (CIs) are shown from separate univariable multinomial logistic regression models. For institutionalisation, “NTP integrated” was the reference category; for notification status, “Mandatory” was the reference category. TB incidence was modelled as log(1+TB incidence). SDI was modelled per 0.1-unit increase. Abbreviations: NTP – national tuberculosis programme; LR – likelihood ratio test.

**Supplementary Table S3.** Crosstabulation summary for NTM institutionalisation status based on survey respondent and reporting status*.

| | **NTP integrated (n = 66)** | **Intent (NTP interest) (n = 72)** | **Outside NTP (n = 30)** | **Orphaned (n = 25)** |
|---|---|---|---|---|
| **Survey respondent** | | | | |
| NTP | 44 (66.7%) | 45 (62.5%) | 17 (56.7%) | 19 (76.0%) |
| NRL | 9 (13.6%) | 17 (23.6%) | 4 (13.3%) | 1 (4.0%) |
| MoH | 9 (13.6%) | 6 (8.3%) | 4 (13.3%) | 2 (8.0%) |
| Expert | 4 (6.1%) | 4 (5.6%) | 5 (16.7%) | 3 (12.0%) |
| **Reporting status** | | | | |
| Mandatory | 27 (40.9%) | 5 (6.9%) | 6 (20.0%) | 3 (12.0%) |
| Voluntary | 8 (12.1%) | 6 (8.3%) | 3 (10.0%) | 2 (8.0%) |
| Under consideration | 20 (30.3%) | 18 (25.0%) | 6 (20.0%) | 3 (12.0%) |
| Not done | 10 (15.2%) | 37 (51.4%) | 14 (46.7%) | 16 (64.0%) |
| Not aware | 1 (1.5%) | 6 (8.3%) | 1 (3.3%) | 1 (4.0%) |

* Data presented as count (percentage). Abbreviations: NTP – national tuberculosis control programme; NRL – national mycobacteriology reference laboratory; MoH – ministry of health.

**Supplementary Table S4.** Topics discussed in NTM discussions by agenda stage, WHO region, and institutionalisation. For each topic, the number and percentage selecting the topic (Yes) within each category is shown*.

| Discussion Domain | Agenda stage | | | | | WHO region | | | | | | Institutionalisation | | | |
|---|---|---|---|---|---|---|---|---|---|---|---|---|---|---|---|
| | Not on agenda/unclear | Latent agenda | Transitional | Emerging agenda | Active agenda | AFRO | AMRO | EMRO | EURO | SEARO | WPRO | Orphaned | Intent (within NTP) | NTP integrated | Outside NTP |
| | n = 35 | n = 40 | n = 3 | n = 80 | n = 35 | n = 44 | n = 50 | n = 15 | n = 47 | n = 9 | n = 28 | n = 25 | n = 72 | n = 66 | n = 30 |
| Clinical management | 6 (17.1%) | 30 (75.0%) | 2 (66.7%) | 69 (86.2%) | 32 (91.4%) | 35 (79.5%) | 29 (58.0%) | 9 (60.0%) | 41 (87.2%) | 7 (77.8%) | 18 (64.3%) | 8 (32.0%) | 49 (68.1%) | 59 (89.4%) | 23 (76.7%) |
| Diagnostics / labs | 6 (17.1%) | 30 (75.0%) | 3 (100.0%) | 72 (90.0%) | 32 (91.4%) | 31 (70.5%) | 38 (76.0%) | 12 (80.0%) | 37 (78.7%) | 7 (77.8%) | 18 (64.3%) | 12 (48.0%) | 46 (63.9%) | 59 (89.4%) | 26 (86.7%) |
| Epidemiology / burden | 4 (11.4%) | 16 (40.0%) | 1 (33.3%) | 41 (51.2%) | 27 (77.1%) | 21 (47.7%) | 24 (48.0%) | 6 (40.0%) | 26 (55.3%) | 1 (11.1%) | 11 (39.3%) | 7 (28.0%) | 28 (38.9%) | 34 (51.5%) | 20 (66.7%) |
| Funding / resources | 3 (8.6%) | 9 (22.5%) | 0 (0.0%) | 25 (31.2%) | 20 (57.1%) | 17 (38.6%) | 12 (24.0%) | 5 (33.3%) | 14 (29.8%) | 2 (22.2%) | 7 (25.0%) | 1 (4.0%) | 25 (34.7%) | 20 (30.3%) | 11 (36.7%) |
| No specific aspects | 27 (77.1%) | 1 (2.5%) | 0 (0.0%) | 0 (0.0%) | 0 (0.0%) | 6 (13.6%) | 9 (18.0%) | 3 (20.0%) | 3 (6.4%) | 1 (11.1%) | 6 (21.4%) | 10 (40.0%) | 15 (20.8%) | 1 (1.5%) | 2 (6.7%) |
| TB integration | 2 (5.7%) | 6 (15.0%) | 0 (0.0%) | 29 (36.2%) | 13 (37.1%) | 15 (34.1%) | 11 (22.0%) | 6 (40.0%) | 12 (25.5%) | 1 (11.1%) | 5 (17.9%) | 0 (0.0%) | 15 (20.8%) | 31 (47.0%) | 4 (13.3%) |
| Training / awareness | 4 (11.4%) | 14 (35.0%) | 1 (33.3%) | 36 (45.0%) | 27 (77.1%) | 20 (45.5%) | 19 (38.0%) | 5 (33.3%) | 23 (48.9%) | 3 (33.3%) | 12 (42.9%) | 5 (20.0%) | 29 (40.3%) | 29 (43.9%) | 19 (63.3%) |

* Data presented as count (percentage). Abbreviations: AFRO – African Region; AMRO – Regions of Americas; EMRO – Eastern Mediterranean Region; EURO – European Region; SEARO – South-East Asia Region; WPRO – Western Pacific Region; NTP – national tuberculosis control programme.

**Supplementary Table S5.** Topics discussed in NTM discussions by UNIDO developmental status and economic groupings. For each topic the number and percentage selecting the topic (Yes) within each category is shown*.

| Discussion Domain | UNIDO Developmental Status | | | | | | Economic Groupings | | | | | | | | |
|---|---|---|---|---|---|---|---|---|---|---|---|---|---|---|---|
| | High income industrial | High income industrializing | Middle income industrial | Middle income industrializing | Low income | Emerging industrial | APEC | ASEAN | BRICS | CARICOM | CIS | ECOWAS | EU | G20 | OECD |
| | n = 68 | n = 15 | n = 15 | n = 73 | n = 22 | n = 10 | n = 41 | n = 9 | n = 9 | n = 11 | n = 8 | n = 15 | n = 27 | n = 39 | n = 59 |
| Clinical management | 49 (72.1%) | 6 (40.0%) | 14 (93.3%) | 55 (75.3%) | 15 (68.2%) | 7 (70.0%) | 25 (61.0%) | 6 (66.7%) | 6 (66.7%) | 4 (36.4%) | 8 (100.0%) | 14 (93.3%) | 24 (88.9%) | 27 (69.2%) | 46 (78.0%) |
| Diagnostics / labs | 50 (73.5%) | 7 (46.7%) | 13 (86.7%) | 57 (78.1%) | 16 (72.7%) | 8 (80.0%) | 29 (70.7%) | 7 (77.8%) | 7 (77.8%) | 5 (45.5%) | 7 (87.5%) | 14 (93.3%) | 20 (74.1%) | 29 (74.4%) | 45 (76.3%) |
| Epidemiology / burden | 35 (51.5%) | 5 (33.3%) | 10 (66.7%) | 29 (39.7%) | 10 (45.5%) | 4 (40.0%) | 20 (48.8%) | 3 (33.3%) | 5 (55.6%) | 4 (36.4%) | 4 (50.0%) | 10 (66.7%) | 16 (59.3%) | 23 (59.0%) | 33 (55.9%) |
| Funding / resources | 19 (27.9%) | 1 (6.7%) | 4 (26.7%) | 23 (31.5%) | 10 (45.5%) | 3 (30.0%) | 9 (22.0%) | 0 (0.0%) | 2 (22.2%) | 1 (9.1%) | 3 (37.5%) | 7 (46.7%) | 9 (33.3%) | 12 (30.8%) | 17 (28.8%) |
| No specific aspects | 10 (14.7%) | 4 (26.7%) | 0 (0.0%) | 10 (13.7%) | 4 (18.2%) | 1 (10.0%) | 7 (17.1%) | 1 (11.1%) | 1 (11.1%) | 4 (36.4%) | 0 (0.0%) | 1 (6.7%) | 3 (11.1%) | 5 (12.8%) | 6 (10.2%) |
| TB integration | 9 (13.2%) | 6 (40.0%) | 4 (26.7%) | 23 (31.5%) | 8 (36.4%) | 5 (50.0%) | 6 (14.6%) | 2 (22.2%) | 4 (44.4%) | 4 (36.4%) | 5 (62.5%) | 6 (40.0%) | 6 (22.2%) | 5 (12.8%) | 8 (13.6%) |
| Training / awareness | 30 (44.1%) | 2 (13.3%) | 9 (60.0%) | 32 (43.8%) | 9 (40.9%) | 5 (50.0%) | 17 (41.5%) | 2 (22.2%) | 3 (33.3%) | 3 (27.3%) | 4 (50.0%) | 8 (53.3%) | 14 (51.9%) | 18 (46.2%) | 27 (45.8%) |

* Data presented as count (percentage). Abbreviations: APEC – Asia-Pacific Economic Cooperation; ASEAN – Association of Southeast Asian Nations; CARICOM – Caribbean Community; CIS – Commonwealth of Independent States; ECOWAS – Economic Community of West African States; EU – European Union; G20 – Group of 20; OECD – Organization for Economic Co-operation and Development.

**Supplementary Table S6.** Pulmonary and extra-pulmonary NTM discussion frequency, stratified by NTM institutionalisation status, respondent type, different geo-memberships, development status, and economical groupings*.

| Classifier | NTM form | All the time | Often | Occasionally / Sometimes | Never |
|---|---|---|---|---|---|
| **Institutionalisation status** | | | | | |
| NTP integrated | Pulmonary | 24 (36.4%) | 21 (31.8%) | 20 (30.3%) | 1 (1.5%) |
| | Extra-pulmonary | 11 (16.7%) | 15 (22.7%) | 33 (50.0%) | 7 (10.6%) |
| Intent (NTP interest) | Pulmonary | 16 (22.2%) | 18 (25.0%) | 25 (34.7%) | 13 (18.1%) |
| | Extra-pulmonary | 4 (5.6%) | 10 (13.9%) | 31 (43.1%) | 27 (37.5%) |
| Outside NTP | Pulmonary | 11 (36.7%) | 10 (33.3%) | 6 (20.0%) | 3 (10.0%) |
| | Extra-pulmonary | 7 (23.3%) | 8 (26.7%) | 10 (33.3%) | 5 (16.7%) |
| Orphaned | Pulmonary | 2 (8.0%) | 2 (8.0%) | 11 (44.0%) | 10 (40.0%) |
| | Extra-pulmonary | 0 (0.0%) | 4 (16.0%) | 9 (36.0%) | 12 (48.0%) |
| **Survey respondent** | | | | | |
| NTP | Pulmonary | 34 (27.2%) | 31 (24.8%) | 43 (34.4%) | 17 (13.6%) |
| | Extra-pulmonary | 12 (9.6%) | 23 (18.4%) | 55 (44.0%) | 35 (28.0%) |
| NRL | Pulmonary | 8 (25.8%) | 12 (38.7%) | 8 (25.8%) | 3 (9.7%) |
| | Extra-pulmonary | 7 (22.6%) | 7 (22.6%) | 13 (41.9%) | 4 (12.9%) |
| MoH | Pulmonary | 4 (19.0%) | 4 (19.0%) | 8 (38.1%) | 5 (23.8%) |
| | Extra-pulmonary | 1 (4.8%) | 2 (9.5%) | 8 (38.1%) | 10 (47.6%) |
| Expert | Pulmonary | 7 (43.8%) | 4 (25.0%) | 3 (18.8%) | 2 (12.5%) |
| | Extra-pulmonary | 2 (12.5%) | 5 (31.2%) | 7 (43.8%) | 2 (12.5%) |
| **WHO Regions** | | | | | |
| Global | Pulmonary | 53 (27.5%) | 51 (26.4%) | 62 (32.1%) | 27 (14.0%) |
| | Extra-pulmonary | 22 (11.4%) | 37 (19.2%) | 83 (43.0%) | 51 (26.4%) |
| AFRO | Pulmonary | 11 (25.0%) | 9 (20.5%) | 17 (38.6%) | 7 (15.9%) |
| | Extra-pulmonary | 3 (6.8%) | 4 (9.1%) | 21 (47.7%) | 16 (36.4%) |
| AMRO | Pulmonary | 15 (30.0%) | 8 (16.0%) | 19 (38.0%) | 8 (16.0%) |
| | Extra-pulmonary | 8 (16.0%) | 7 (14.0%) | 20 (40.0%) | 15 (30.0%) |
| EMRO | Pulmonary | 2 (13.3%) | 5 (33.3%) | 5 (33.3%) | 3 (20.0%) |
| | Extra-pulmonary | 0 (0.0%) | 7 (46.7%) | 4 (26.7%) | 4 (26.7%) |
| EURO | Pulmonary | 18 (38.3%) | 18 (38.3%) | 8 (17.0%) | 3 (6.4%) |
| | Extra-pulmonary | 9 (19.1%) | 9 (19.1%) | 23 (48.9%) | 6 (12.8%) |
| SEARO | Pulmonary | 1 (11.1%) | 3 (33.3%) | 4 (44.4%) | 1 (11.1%) |
| | Extra-pulmonary | 0 (0.0%) | 3 (33.3%) | 4 (44.4%) | 2 (22.2%) |
| WPRO | Pulmonary | 6 (21.4%) | 8 (28.6%) | 9 (32.1%) | 5 (17.9%) |
| | Extra-pulmonary | 2 (7.1%) | 7 (25.0%) | 11 (39.3%) | 8 (28.6%) |
| **M49 Standard Regions** | | | | | |
| Africa | Pulmonary | 11 (22.9%) | 10 (20.8%) | 20 (41.7%) | 7 (14.6%) |
| | Extra-pulmonary | 3 (6.2%) | 6 (12.5%) | 23 (47.9%) | 16 (33.3%) |
| Americas | Pulmonary | 15 (30.0%) | 8 (16.0%) | 19 (38.0%) | 8 (16.0%) |
| | Extra-pulmonary | 8 (16.0%) | 7 (14.0%) | 20 (40.0%) | 15 (30.0%) |
| Asia | Pulmonary | 14 (34.1%) | 12 (29.3%) | 9 (22.0%) | 6 (14.6%) |
| | Extra-pulmonary | 3 (7.3%) | 12 (29.3%) | 14 (34.1%) | 12 (29.3%) |
| Europe | Pulmonary | 12 (31.6%) | 15 (39.5%) | 8 (21.1%) | 3 (7.9%) |
| | Extra-pulmonary | 7 (18.4%) | 7 (18.4%) | 21 (55.3%) | 3 (7.9%) |
| Oceania | Pulmonary | 1 (6.2%) | 6 (37.5%) | 6 (37.5%) | 3 (18.8%) |
| | Extra-pulmonary | 1 (6.2%) | 5 (31.2%) | 5 (31.2%) | 5 (31.2%) |
| **M49 Standard – UNIDO Simplified Sub Regions** | | | | | |
| ***M49 Standard – African Region*** | | | | | |
| Central Africa | Pulmonary | 0 (0.0%) | 1 (11.1%) | 3 (33.3%) | 5 (55.6%) |

| | | | | | |
|---|---|---|---|---|---|
| | Extra-pulmonary | 0 (0.0%) | 1 (11.1%) | 3 (33.3%) | 5 (55.6%) |
| Eastern Africa | Pulmonary | 5 (35.7%) | 3 (21.4%) | 5 (35.7%) | 1 (7.1%) |
| | Extra-pulmonary | 1 (7.1%) | 2 (14.3%) | 6 (42.9%) | 5 (35.7%) |
| Northern Africa | Pulmonary | 0 (0.0%) | 0 (0.0%) | 4 (100.0%) | 0 (0.0%) |
| | Extra-pulmonary | 0 (0.0%) | 1 (25.0%) | 3 (75.0%) | 0 (0.0%) |
| Southern Africa | Pulmonary | 1 (20.0%) | 1 (20.0%) | 3 (60.0%) | 0 (0.0%) |
| | Extra-pulmonary | 0 (0.0%) | 0 (0.0%) | 4 (80.0%) | 1 (20.0%) |
| Western Africa | Pulmonary | 5 (31.2%) | 5 (31.2%) | 5 (31.2%) | 1 (6.2%) |
| | Extra-pulmonary | 2 (12.5%) | 2 (12.5%) | 7 (43.8%) | 5 (31.2%) |
| ***M49 Standard – American Region*** | | | | | |
| Caribbean | Pulmonary | 4 (28.6%) | 2 (14.3%) | 5 (35.7%) | 3 (21.4%) |
| | Extra-pulmonary | 3 (21.4%) | 1 (7.1%) | 3 (21.4%) | 7 (50.0%) |
| Central America | Pulmonary | 3 (42.9%) | 1 (14.3%) | 2 (28.6%) | 1 (14.3%) |
| | Extra-pulmonary | 0 (0.0%) | 1 (14.3%) | 5 (71.4%) | 1 (14.3%) |
| Northern America | Pulmonary | 4 (23.5%) | 4 (23.5%) | 5 (29.4%) | 4 (23.5%) |
| | Extra-pulmonary | 2 (11.8%) | 5 (29.4%) | 4 (23.5%) | 6 (35.3%) |
| South America | Pulmonary | 4 (33.3%) | 1 (8.3%) | 7 (58.3%) | 0 (0.0%) |
| | Extra-pulmonary | 3 (25.0%) | 0 (0.0%) | 8 (66.7%) | 1 (8.3%) |
| ***M49 Standard – Asian Region*** | | | | | |
| Central Asia | Pulmonary | 2 (66.7%) | 1 (33.3%) | 0 (0.0%) | 0 (0.0%) |
| | Extra-pulmonary | 0 (0.0%) | 0 (0.0%) | 1 (33.3%) | 2 (66.7%) |
| Eastern Asia | Pulmonary | 3 (50.0%) | 1 (16.7%) | 1 (16.7%) | 1 (16.7%) |
| | Extra-pulmonary | 1 (16.7%) | 1 (16.7%) | 3 (50.0%) | 1 (16.7%) |
| South-eastern Asia | Pulmonary | 2 (22.2%) | 2 (22.2%) | 4 (44.4%) | 1 (11.1%) |
| | Extra-pulmonary | 0 (0.0%) | 3 (33.3%) | 4 (44.4%) | 2 (22.2%) |
| Southern Asia | Pulmonary | 1 (11.1%) | 3 (33.3%) | 3 (33.3%) | 2 (22.2%) |
| | Extra-pulmonary | 0 (0.0%) | 2 (22.2%) | 3 (33.3%) | 4 (44.4%) |
| Western Asia | Pulmonary | 6 (42.9%) | 5 (35.7%) | 1 (7.1%) | 2 (14.3%) |
| | Extra-pulmonary | 2 (14.3%) | 6 (42.9%) | 3 (21.4%) | 3 (21.4%) |
| ***M49 Standard – European Region*** | | | | | |
| Eastern Europe | Pulmonary | 2 (22.2%) | 5 (55.6%) | 2 (22.2%) | 0 (0.0%) |
| | Extra-pulmonary | 1 (11.1%) | 3 (33.3%) | 5 (55.6%) | 0 (0.0%) |
| Northern Europe | Pulmonary | 3 (30.0%) | 3 (30.0%) | 3 (30.0%) | 1 (10.0%) |
| | Extra-pulmonary | 1 (10.0%) | 2 (20.0%) | 6 (60.0%) | 1 (10.0%) |
| Southern Europe | Pulmonary | 5 (41.7%) | 4 (33.3%) | 2 (16.7%) | 1 (8.3%) |
| | Extra-pulmonary | 3 (25.0%) | 0 (0.0%) | 8 (66.7%) | 1 (8.3%) |
| Western Europe | Pulmonary | 2 (28.6%) | 3 (42.9%) | 1 (14.3%) | 1 (14.3%) |
| | Extra-pulmonary | 2 (28.6%) | 2 (28.6%) | 2 (28.6%) | 1 (14.3%) |
| ***M49 Standard – Oceania Region*** | | | | | |
| Australia and New Zealand | Pulmonary | 1 (12.5%) | 5 (62.5%) | 2 (25.0%) | 0 (0.0%) |
| | Extra-pulmonary | 1 (12.5%) | 4 (50.0%) | 3 (37.5%) | 0 (0.0%) |
| Melanesia | Pulmonary | 0 (0.0%) | 0 (0.0%) | 2 (66.7%) | 1 (33.3%) |
| | Extra-pulmonary | 0 (0.0%) | 1 (33.3%) | 1 (33.3%) | 1 (33.3%) |
| Micronesia | Pulmonary | 0 (0.0%) | 1 (20.0%) | 2 (40.0%) | 2 (40.0%) |
| | Extra-pulmonary | 0 (0.0%) | 0 (0.0%) | 1 (20.0%) | 4 (80.0%) |
| **M49 Standard Other Groupings** | | | | | |
| LDCs | Pulmonary | 7 (20.0%) | 8 (22.9%) | 15 (42.9%) | 5 (14.3%) |
| | Extra-pulmonary | 1 (2.9%) | 6 (17.1%) | 15 (42.9%) | 13 (37.1%) |
| LLDCs | Pulmonary | 8 (29.6%) | 6 (22.2%) | 9 (33.3%) | 4 (14.8%) |
| | Extra-pulmonary | 4 (14.8%) | 2 (7.4%) | 13 (48.1%) | 8 (29.6%) |
| SIDS | Pulmonary | 7 (21.9%) | 4 (12.5%) | 13 (40.6%) | 8 (25.0%) |
| | Extra-pulmonary | 3 (9.4%) | 4 (12.5%) | 9 (28.1%) | 16 (50.0%) |
| **UNIDO Development Groups** | | | | | |
| High-income industrial | Pulmonary | 20 (29.4%) | 23 (33.8%) | 16 (23.5%) | 9 (13.2%) |

| | | | | | |
|---|---|---|---|---|---|
| economies | Extra-pulmonary | 11 (16.2%) | 16 (23.5%) | 28 (41.2%) | 13 (19.1%) |
| High-income industrializing economies | Pulmonary | 3 (20.0%) | 3 (20.0%) | 5 (33.3%) | 4 (26.7%) |
| | Extra-pulmonary | 2 (13.3%) | 3 (20.0%) | 3 (20.0%) | 7 (46.7%) |
| Middle-income industrial economies | Pulmonary | 6 (40.0%) | 5 (33.3%) | 4 (26.7%) | 0 (0.0%) |
| | Extra-pulmonary | 3 (20.0%) | 4 (26.7%) | 8 (53.3%) | 0 (0.0%) |
| Middle-income industrializing economies | Pulmonary | 18 (24.7%) | 15 (20.5%) | 30 (41.1%) | 10 (13.7%) |
| | Extra-pulmonary | 5 (6.8%) | 11 (15.1%) | 34 (46.6%) | 23 (31.5%) |
| Low-income economies | Pulmonary | 6 (27.3%) | 5 (22.7%) | 7 (31.8%) | 4 (18.2%) |
| | Extra-pulmonary | 1 (4.5%) | 3 (13.6%) | 10 (45.5%) | 8 (36.4%) |
| Emerging industrial economies | Pulmonary | 2 (20.0%) | 3 (30.0%) | 4 (40.0%) | 1 (10.0%) |
| | Extra-pulmonary | 0 (0.0%) | 3 (30.0%) | 5 (50.0%) | 2 (20.0%) |
| **Economic Groupings** | | | | | |
| APEC | Pulmonary | 11 (26.8%) | 13 (31.7%) | 10 (24.4%) | 7 (17.1%) |
| | Extra-pulmonary | 4 (9.8%) | 12 (29.3%) | 15 (36.6%) | 10 (24.4%) |
| ASEAN | Pulmonary | 2 (22.2%) | 2 (22.2%) | 4 (44.4%) | 1 (11.1%) |
| | Extra-pulmonary | 0 (0.0%) | 3 (33.3%) | 4 (44.4%) | 2 (22.2%) |
| BRICS | Pulmonary | 2 (22.2%) | 3 (33.3%) | 3 (33.3%) | 1 (11.1%) |
| | Extra-pulmonary | 1 (11.1%) | 3 (33.3%) | 4 (44.4%) | 1 (11.1%) |
| CARICOM | Pulmonary | 2 (18.2%) | 1 (9.1%) | 5 (45.5%) | 3 (27.3%) |
| | Extra-pulmonary | 1 (9.1%) | 0 (0.0%) | 3 (27.3%) | 7 (63.6%) |
| CIS | Pulmonary | 3 (37.5%) | 4 (50.0%) | 1 (12.5%) | 0 (0.0%) |
| | Extra-pulmonary | 1 (12.5%) | 1 (12.5%) | 4 (50.0%) | 2 (25.0%) |
| ECOWAS | Pulmonary | 5 (33.3%) | 4 (26.7%) | 5 (33.3%) | 1 (6.7%) |
| | Extra-pulmonary | 2 (13.3%) | 2 (13.3%) | 7 (46.7%) | 4 (26.7%) |
| EU | Pulmonary | 8 (29.6%) | 13 (48.1%) | 3 (11.1%) | 3 (11.1%) |
| | Extra-pulmonary | 6 (22.2%) | 6 (22.2%) | 12 (44.4%) | 3 (11.1%) |
| G20 | Pulmonary | 12 (30.8%) | 14 (35.9%) | 8 (20.5%) | 5 (12.8%) |
| | Extra-pulmonary | 6 (15.4%) | 12 (30.8%) | 14 (35.9%) | 7 (17.9%) |
| OECD | Pulmonary | 20 (33.9%) | 19 (32.2%) | 14 (23.7%) | 6 (10.2%) |
| | Extra-pulmonary | 10 (16.9%) | 15 (25.4%) | 26 (44.1%) | 8 (13.6%) |

* Data presented as count (percentage). Abbreviations: AFRO – African Region; AMRO – Regions of Americas; EMRO – Eastern Mediterranean Region; EURO – European Region; SEARO – South-East Asia Region; WPRO – Western Pacific Region; NTP – national tuberculosis control programme; NRL – national mycobacteriology reference laboratory; MoH – ministry of health; LDCs – least developed countries; LLDCs – landlocked developing countries; SIDS – small island developing states; APEC – Asia-Pacific Economic Cooperation; ASEAN – Association of Southeast Asian Nations; CARICOM – Caribbean Community; CIS – Commonwealth of Independent States; ECOWAS – Economic Community of West African States; EU – European Union; G20 – Group of 20; OECD – Organization for Economic Co-operation and Development.

**Supplementary Table S7.** Descriptive summary for relative region-defined momentum scores, stratified by different geo-memberships, development status, and economical groupings [*, **, ***, ****].

| Geo-classifier | Momentum Group | n (%) | Overall Composite | | NTM Clinical Management | | | NTM Diagnostics | | | NTM Surveillance | | |
|---|---|---|---|---|---|---|---|---|---|---|---|---|---|
| | | | Mean (SD) | Median (IQR) | Mean (SD) | Median (IQR) | % Not aware | Mean (SD) | Median (IQR) | % Not aware | Mean (SD) | Median (IQR) | % Not aware |
| **WHO Regions** | | | | | | | | | | | | | |
| AFRO (n = 44) | Higher momentum | 19 (43.2%) | 3.28 (0.55) | 3.00 (3.00–4.00) | 3.37 (0.60) | 3.00 (3.00–4.00) | - | 3.37 (0.60) | 3.00 (3.00–4.00) | - | 3.11 (0.74) | 3.00 (3.00–4.00) | - |
| | Lower momentum | 25 (56.8%) | 1.51 (0.71) | 1.67 (1.00–2.00) | 1.56 (1.04) | 2.00 (1.00–2.00) | 16.0% | 1.68 (0.95) | 2.00 (1.00–2.00) | 12.0% | 1.28 (0.84) | 1.00 (1.00–2.00) | 16.0% |
| AMRO (n = 50) | Higher momentum | 34 (68.0%) | 2.53 (0.68) | 2.33 (2.00–3.00) | 2.62 (0.74) | 3.00 (2.00–3.00) | - | 2.62 (0.78) | 3.00 (2.00–3.00) | - | 2.35 (0.95) | 2.00 (2.00–3.00) | - |
| | Lower momentum | 16 (32.0%) | 0.83 (0.37) | 1.00 (1.00–1.00) | 0.62 (0.50) | 1.00 (0.00–1.00) | 37.5% | 0.94 (0.77) | 1.00 (0.50–1.00) | 25.0% | 0.94 (0.68) | 1.00 (1.00–1.00) | 18.8% |
| EMRO (n = 15) | Higher momentum | 8 (53.3%) | 3.42 (0.50) | 3.50 (3.00–3.83) | 3.50 (0.53) | 3.50 (3.00–4.00) | - | 3.50 (0.76) | 4.00 (3.00–4.00) | - | 3.25 (0.46) | 3.00 (3.00–3.50) | - |
| | Lower momentum | 7 (46.7%) | 1.19 (0.84) | 1.67 (0.00–1.67) | 1.71 (1.25) | 2.00 (0.00–3.00) | 28.6% | 1.00 (1.00) | 1.00 (0.00–2.00) | 42.9% | 0.86 (0.69) | 1.00 (0.00–1.00) | 28.6% |
| EURO (n = 47) | Higher momentum | 26 (55.3%) | 3.13 (0.48) | 3.00 (2.67–3.33) | 3.19 (0.49) | 3.00 (3.00–3.00) | - | 3.31 (0.47) | 3.00 (3.00–4.00) | - | 2.88 (0.82) | 3.00 (2.00–3.00) | - |
| | Lower momentum | 21 (44.7%) | 1.54 (0.47) | 1.67 (1.33–2.00) | 1.43 (0.68) | 2.00 (1.00–2.00) | 9.5% | 1.71 (0.72) | 2.00 (1.00–2.00) | 4.8% | 1.48 (0.51) | 1.00 (1.00–2.00) | - |
| SEARO (n = 9) | Higher momentum | 1 (11.1%) | 2.33 (NA) | 2.33 (2.33–2.33) | 2.00 (NA) | 2.00 (2.00–2.00) | - | 1.00 (NA) | 1.00 (1.00–1.00) | - | 4.00 (NA) | 4.00 (4.00–4.00) | - |
| | Lower momentum | 8 (88.9%) | 2.00 (0.50) | 2.00 (1.67–2.17) | 2.50 (0.76) | 2.00 (2.00–3.00) | - | 2.00 (0.76) | 2.00 (1.50–2.50) | - | 1.50 (0.53) | 1.50 (1.00–2.00) | - |
| WPRO (n = 28) | Higher momentum | 11 (39.3%) | 3.06 (0.39) | 3.00 (2.67–3.33) | 3.18 (0.40) | 3.00 (3.00–3.00) | - | 3.09 (0.30) | 3.00 (3.00–3.00) | - | 2.91 (0.70) | 3.00 (2.00–3.00) | - |
| | Lower momentum | 17 (60.7%) | 1.47 (0.51) | 1.33 (1.00–2.00) | 1.35 (0.86) | 1.00 (1.00–2.00) | 11.8% | 1.59 (0.71) | 1.00 (1.00–2.00) | - | 1.47 (0.62) | 1.00 (1.00–2.00) | - |
| **M49 Standard** | | | | | | | | | | | | | |
| Africa (n = 48) | Higher momentum | 20 (41.7%) | 3.32 (0.56) | 3.17 (3.00–4.00) | 3.40 (0.60) | 3.00 (3.00–4.00) | - | 3.40 (0.60) | 3.00 (3.00–4.00) | - | 3.15 (0.75) | 3.00 (3.00–4.00) | - |
| | Lower momentum | 28 (58.3%) | 1.46 (0.73) | 1.67 (1.00–2.00) | 1.57 (1.07) | 2.00 (1.00–2.00) | 17.9% | 1.57 (1.00) | 2.00 (1.00–2.00) | 17.9% | 1.25 (0.84) | 1.00 (1.00–2.00) | 17.9% |
| Americas (n = 50) | Higher momentum | 34 (68.0%) | 2.53 (0.68) | 2.33 (2.00–3.00) | 2.62 (0.74) | 3.00 (2.00–3.00) | - | 2.62 (0.78) | 3.00 (2.00–3.00) | - | 2.35 (0.95) | 2.00 (2.00–3.00) | - |
| | Lower momentum | 16 (32.0%) | 0.83 (0.37) | 1.00 (1.00–1.00) | 0.62 (0.50) | 1.00 (0.00–1.00) | 37.5% | 0.94 (0.77) | 1.00 (0.50–1.00) | 25.0% | 0.94 (0.68) | 1.00 (1.00–1.00) | 18.8% |
| Asia (n = 41) | Higher momentum | 20 (48.8%) | 3.20 (0.49) | 3.17 (2.83–3.50) | 3.25 (0.55) | 3.00 (3.00–4.00) | - | 3.25 (0.79) | 3.00 (3.00–4.00) | - | 3.10 (0.64) | 3.00 (3.00–3.50) | - |
| | Lower momentum | 21 (51.2%) | 1.67 (0.62) | 1.67 (1.33–2.00) | 1.90 (1.00) | 2.00 (1.00–2.00) | 9.5% | 1.81 (0.81) | 2.00 (1.00–2.00) | 4.8% | 1.29 (0.56) | 1.00 (1.00–2.00) | 4.8% |
| Europe (n = 38) | Higher momentum | 20 (52.6%) | 3.08 (0.49) | 3.00 (2.67–3.33) | 3.15 (0.49) | 3.00 (3.00–3.00) | - | 3.25 (0.44) | 3.00 (3.00–3.50) | - | 2.85 (0.88) | 3.00 (2.00–3.50) | - |
| | Lower momentum | 18 (47.4%) | 1.54 (0.47) | 1.67 (1.33–2.00) | 1.44 (0.70) | 2.00 (1.00–2.00) | 11.1% | 1.67 (0.69) | 2.00 (1.00–2.00) | 5.6% | 1.50 (0.51) | 1.50 (1.00–2.00) | - |
| Oceania (n = 16) | Higher momentum | 5 (31.2%) | 3.00 (0.24) | 3.00 (3.00–3.00) | 3.20 (0.45) | 3.00 (3.00–3.00) | - | 3.00 (0.00) | 3.00 (3.00–3.00) | - | 2.80 (0.45) | 3.00 (3.00–3.00) | - |
| | Lower momentum | 11 (68.8%) | 1.42 (0.52) | 1.00 (1.00–2.00) | 1.27 (0.79) | 1.00 (1.00–2.00) | 9.1% | 1.45 (0.69) | 1.00 (1.00–2.00) | - | 1.55 (0.69) | 1.00 (1.00–2.00) | - |
| **M49 Standard – UNIDO Simplified Sub Regions** | | | | | | | | | | | | | |
| ***M49 Standard – African Region*** | | | | | | | | | | | | | |
| Central Africa (n = 9) | Higher momentum | 1 (11.1%) | 3.00 (NA) | 3.00 (3.00–3.00) | 3.00 (NA) | 3.00 (3.00–3.00) | - | 3.00 (NA) | 3.00 (3.00–3.00) | - | 3.00 (NA) | 3.00 (3.00–3.00) | - |
| | Lower momentum | 8 (88.9%) | 1.08 (0.96) | 1.33 (0.00–2.00) | 1.25 (1.16) | 1.50 (0.00–2.00) | 37.5% | 1.00 (0.93) | 1.00 (0.00–2.00) | 37.5% | 1.00 (0.93) | 1.00 (0.00–2.00) | 37.5% |

| | | | | | | | | | | | | | |
|---|---|---|---|---|---|---|---|---|---|---|---|---|---|
| Eastern Africa (n = 14) | Higher momentum | 4 (28.6%) | 3.50 (0.43) | 3.50 (3.17–3.83) | 3.50 (0.58) | 3.50 (3.00–4.00) | - | 3.50 (0.58) | 3.50 (3.00–4.00) | - | 3.50 (0.58) | 3.50 (3.00–4.00) | - |
| | Lower momentum | 10 (71.4%) | 1.63 (0.53) | 1.67 (1.00–2.00) | 1.70 (0.82) | 1.50 (1.00–2.00) | - | 1.70 (1.06) | 1.50 (1.00–3.00) | 10.0% | 1.50 (0.71) | 1.00 (1.00–2.00) | - |
| Northern Africa (n = 4) | Higher momentum | 1 (25.0%) | 4.00 (NA) | 4.00 (4.00–4.00) | 4.00 (NA) | 4.00 (4.00–4.00) | - | 4.00 (NA) | 4.00 (4.00–4.00) | - | 4.00 (NA) | 4.00 (4.00–4.00) | - |
| | Lower momentum | 3 (75.0%) | 1.22 (1.07) | 1.67 (0.00–2.00) | 1.33 (1.15) | 2.00 (0.00–2.00) | 33.3% | 1.33 (1.15) | 2.00 (0.00–2.00) | 33.3% | 1.00 (1.00) | 1.00 (0.00–2.00) | 33.3% |
| Southern Africa (n = 5) | Higher momentum | 3 (60.0%) | 2.78 (0.38) | 3.00 (2.33–3.00) | 3.00 (1.00) | 3.00 (2.00–4.00) | - | 3.00 (0.00) | 3.00 (3.00–3.00) | - | 2.33 (0.58) | 2.00 (2.00–3.00) | |
| | Lower momentum | 2 (40.0%) | 2.17 (0.24) | 2.17 (2.00–2.33) | 3.50 (0.71) | 3.50 (3.00–4.00) | - | 2.50 (0.71) | 2.50 (2.00–3.00) | - | 0.50 (0.71) | 0.50 (0.00–1.00) | 50.0% |
| Western Africa (n = 16) | Higher momentum | 11 (68.8%) | 3.36 (0.59) | 3.33 (2.67–4.00) | 3.45 (0.52) | 3.00 (3.00–4.00) | - | 3.45 (0.69) | 4.00 (3.00–4.00) | - | 3.18 (0.75) | 3.00 (3.00–4.00) | - |
| | Lower momentum | 5 (31.2%) | 1.60 (0.37) | 1.67 (1.67–1.67) | 1.20 (0.84) | 1.00 (1.00–2.00) | 20.0% | 2.00 (0.71) | 2.00 (2.00–2.00) | - | 1.60 (0.89) | 1.00 (1.00–2.00) | - |
| ***M49 Standard – American Region*** | | | | | | | | | | | | | |
| Caribbean (n = 14) | Higher momentum | 9 (64.3%) | 2.59 (0.85) | 2.00 (2.00–3.00) | 2.67 (0.71) | 3.00 (2.00–3.00) | | 2.56 (1.01) | 2.00 (2.00–3.00) | - | 2.56 (1.01) | 2.00 (2.00–3.00) | - |
| | Lower momentum | 5 (35.7%) | 0.80 (0.45) | 1.00 (1.00–1.00) | 0.60 (0.55) | 1.00 (0.00–1.00) | 40.0% | 1.00 (0.71) | 1.00 (1.00–1.00) | 20.0% | 0.80 (0.45) | 1.00 (1.00–1.00) | 20.0% |
| Central America (n = 7) | Higher momentum | 5 (71.4%) | 2.67 (0.47) | 3.00 (2.33–3.00) | 2.80 (0.45) | 3.00 (3.00–3.00) | - | 2.80 (0.45) | 3.00 (3.00–3.00) | - | 2.40 (0.89) | 3.00 (2.00–3.00) | - |
| | Lower momentum | 2 (28.6%) | 1.00 (0.00) | 1.00 (1.00–1.00) | 0.50 (0.71) | 0.50 (0.00–1.00) | 50.0% | 2.00 (1.41) | 2.00 (1.00–3.00) | - | 0.50 (0.71) | 0.50 (0.00–1.00) | 50.0% |
| Northern America (n = 17) | Higher momentum | 12 (70.6%) | 2.50 (0.72) | 2.33 (2.00–3.00) | 2.83 (0.72) | 3.00 (2.00–3.00) | - | 2.42 (0.79) | 2.00 (2.00–3.00) | - | 2.25 (1.06) | 2.00 (1.50–3.00) | - |
| | Lower momentum | 5 (29.4%) | 0.87 (0.30) | 1.00 (1.00–1.00) | 0.60 (0.55) | 1.00 (0.00–1.00) | 40.0% | 0.60 (0.55) | 1.00 (0.00–1.00) | 40.0% | 1.40 (0.89) | 1.00 (1.00–1.00) | - |
| South America (n = 12) | Higher momentum | 8 (66.7%) | 2.42 (0.64) | 2.50 (2.00–2.83) | 2.12 (0.83) | 2.00 (1.50–3.00) | - | 2.88 (0.64) | 3.00 (2.50–3.00) | - | 2.25 (0.89) | 2.50 (1.50–3.00) | - |
| | Lower momentum | 4 (33.3%) | 0.75 (0.50) | 1.00 (0.50–1.00) | 0.75 (0.50) | 1.00 (0.50–1.00) | 25.0% | 0.75 (0.50) | 1.00 (0.50–1.00) | 25.0% | 0.75 (0.50) | 1.00 (0.50–1.00) | 25.0% |
| ***M49 Standard – Asian Region*** | | | | | | | | | | | | | |
| Central Asia (n = 3) | Higher momentum | 2 (66.7%) | 3.50 (0.71) | 3.50 (3.00–4.00) | 3.50 (0.71) | 3.50 (3.00–4.00) | - | 3.50 (0.71) | 3.50 (3.00–4.00) | - | 3.50 (0.71) | 3.50 (3.00–4.00) | - |
| | Lower momentum | 1 (33.3%) | 1.00 (NA) | 1.00 (1.00–1.00) | 1.00 (NA) | 1.00 (1.00–1.00) | - | 1.00 (NA) | 1.00 (1.00–1.00) | - | 1.00 (NA) | 1.00 (1.00–1.00) | - |
| Eastern Asia (n = 6) | Higher momentum | 3 (50.0%) | 3.00 (0.33) | 3.00 (2.67–3.33) | 3.00 (0.00) | 3.00 (3.00–3.00) | - | 3.00 (0.00) | 3.00 (3.00–3.00) | 0.0% | 3.00 (1.00) | 3.00 (2.00–4.00) | 0.0% |
| | Lower momentum | 3 (50.0%) | 1.67 (0.67) | 1.67 (1.00–2.33) | 2.00 (1.00) | 2.00 (1.00–3.00) | - | 1.67 (0.58) | 2.00 (1.00–2.00) | 0.0% | 1.33 (0.58) | 1.00 (1.00–2.00) | 0.0% |
| South-eastern Asia (n = 9) | Higher momentum | 4 (44.4%) | 3.00 (0.72) | 2.83 (2.50–3.50) | 3.00 (0.82) | 3.00 (2.50–3.50) | - | 2.75 (1.26) | 3.00 (2.00–3.50) | - | 3.25 (0.96) | 3.50 (2.50–4.00) | |
| | Lower momentum | 5 (55.6%) | 1.80 (0.77) | 1.67 (1.33–2.00) | 2.00 (1.58) | 2.00 (1.00–3.00) | 20.0% | 2.00 (1.00) | 2.00 (1.00–3.00) | - | 1.40 (0.55) | 1.00 (1.00–2.00) | |
| Southern Asia (n = 9) | Higher momentum | - | - | - | - | - | - | - | - | - | - | - | - |
| | Lower momentum | 9 (100.0%) | 1.59 (0.68) | 1.67 (1.33–2.00) | 1.89 (0.78) | 2.00 (2.00–2.00) | 11.1% | 1.67 (0.87) | 2.00 (1.00–2.00) | 11.1% | 1.22 (0.67) | 1.00 (1.00–2.00) | 11.1% |
| Western Asia (n = 14) | Higher momentum | 11 (78.6%) | 3.27 (0.42) | 3.33 (3.00–3.67) | 3.36 (0.50) | 3.00 (3.00–4.00) | - | 3.45 (0.69) | 4.00 (3.00–4.00) | - | 3.00 (0.45) | 3.00 (3.00–3.00) | - |
| | Lower momentum | 3 (21.4%) | 1.89 (0.19) | 2.00 (1.67–2.00) | 2.00 (1.00) | 2.00 (1.00–3.00) | - | 2.33 (0.58) | 2.00 (2.00–3.00) | - | 1.33 (0.58) | 1.00 (1.00–2.00) | - |
| ***M49 Standard – European Region*** | | | | | | | | | | | | | |
| Eastern Europe (n = 9) | Higher momentum | 6 (66.7%) | 3.39 (0.57) | 3.33 (3.00–4.00) | 3.33 (0.52) | 3.00 (3.00–4.00) | - | 3.50 (0.55) | 3.50 (3.00–4.00) | 0.0% | 3.33 (0.82) | 3.50 (3.00–4.00) | 0.0% |
| | Lower momentum | 3 (33.3%) | 1.78 (0.38) | 2.00 (1.33–2.00) | 1.67 (0.58) | 2.00 (1.00–2.00) | - | 1.67 (0.58) | 2.00 (1.00–2.00) | 0.0% | 2.00 (0.00) | 2.00 (2.00–2.00) | 0.0% |

| | | | | | | | | | | | | | |
|---|---|---|---|---|---|---|---|---|---|---|---|---|---|
| Northern Europe (n = 10) | Higher momentum | 4 (40.0%) | 3.08 (0.63) | 2.83 (2.67–3.50) | 3.50 (0.58) | 3.50 (3.00–4.00) | - | 3.25 (0.50) | 3.00 (3.00–3.50) | - | 2.50 (1.00) | 2.00 (2.00–3.00) | - |
| | Lower momentum | 6 (60.0%) | 1.33 (0.60) | 1.50 (1.00–1.67) | 1.17 (0.98) | 1.50 (0.00–2.00) | 33.3% | 1.50 (0.84) | 2.00 (1.00–2.00) | 16.7% | 1.33 (0.52) | 1.00 (1.00–2.00) | - |
| Southern Europe (n = 12) | Higher momentum | 7 (58.3%) | 2.90 (0.25) | 3.00 (2.67–3.00) | 2.86 (0.38) | 3.00 (3.00–3.00) | - | 3.00 (0.00) | 3.00 (3.00–3.00) | - | 2.86 (0.69) | 3.00 (2.00–3.00) | - |
| | Lower momentum | 5 (41.7%) | 1.60 (0.43) | 1.67 (1.33–2.00) | 1.60 (0.55) | 2.00 (1.00–2.00) | - | 1.60 (0.55) | 2.00 (1.00–2.00) | - | 1.60 (0.55) | 2.00 (1.00–2.00) | - |
| Western Europe (n = 7) | Higher momentum | 3 (42.9%) | 2.89 (0.51) | 3.00 (2.33–3.33) | 3.00 (0.00) | 3.00 (3.00–3.00) | - | 3.33 (0.58) | 3.00 (3.00–4.00) | - | 2.33 (1.15) | 3.00 (1.00–3.00) | - |
| | Lower momentum | 4 (57.1%) | 1.58 (0.42) | 1.67 (1.33–1.83) | 1.50 (0.58) | 1.50 (1.00–2.00) | - | 2.00 (0.82) | 2.00 (1.50–2.50) | - | 1.25 (0.50) | 1.00 (1.00–1.50) | - |
| ***M49 Standard – Oceania Region*** | | | | | | | | | | | | | |
| Australia and New Zealand (n = 8) | Higher momentum | 3 (37.5%) | 3.00 (0.33) | 3.00 (2.67–3.33) | 3.33 (0.58) | 3.00 (3.00–4.00) | - | 3.00 (0.00) | 3.00 (3.00–3.00) | - | 2.67 (0.58) | 3.00 (2.00–3.00) | - |
| | Lower momentum | 5 (62.5%) | 1.67 (0.62) | 2.00 (1.00–2.00) | 1.80 (0.84) | 2.00 (1.00–2.00) | - | 1.40 (0.55) | 1.00 (1.00–2.00) | - | 1.80 (0.84) | 2.00 (1.00–2.00) | - |
| Melanesia (n = 3) | Higher momentum | - | - | - | - | - | - | - | - | - | - | - | - |
| | Lower momentum | 3 (100.0%) | 1.44 (0.38) | 1.67 (1.00–1.67) | 0.67 (0.58) | 1.00 (0.00–1.00) | 33.3% | 2.00 (1.00) | 2.00 (1.00–3.00) | - | 1.67 (0.58) | 2.00 (1.00–2.00) | - |
| Micronesia (n = 5) | Higher momentum | 2 (40.0%) | 3.00 (0.00) | 3.00 (3.00–3.00) | 3.00 (0.00) | 3.00 (3.00–3.00) | - | 3.00 (0.00) | 3.00 (3.00–3.00) | - | 3.00 (0.00) | 3.00 (3.00–3.00) | - |
| | Lower momentum | 3 (60.0%) | 1.00 (0.00) | 1.00 (1.00–1.00) | 1.00 (0.00) | 1.00 (1.00–1.00) | - | 1.00 (0.00) | 1.00 (1.00–1.00) | - | 1.00 (0.00) | 1.00 (1.00–1.00) | - |
| **M49 Standard Other Groupings** | | | | | | | | | | | | | |
| Least developed countries LDCs (n = 35) | Higher momentum | 16 (45.7%) | 3.27 (0.56) | 3.17 (2.83–3.83) | 3.31 (0.60) | 3.00 (3.00–4.00) | - | 3.25 (0.86) | 3.00 (3.00–4.00) | - | 3.25 (0.68) | 3.00 (3.00–4.00) | - |
| | Lower momentum | 19 (54.3%) | 1.46 (0.82) | 1.67 (1.00–2.00) | 1.53 (1.22) | 1.00 (1.00–2.00) | 21.1% | 1.58 (1.12) | 2.00 (1.00–3.00) | 21.1% | 1.26 (0.93) | 1.00 (1.00–2.00) | 21.1% |
| Landlocked developing countries LLDCs (n = 27) | Higher momentum | 12 (44.4%) | 3.08 (0.45) | 3.00 (2.83–3.33) | 3.08 (0.51) | 3.00 (3.00–3.00) | - | 3.33 (0.49) | 3.00 (3.00–4.00) | - | 2.83 (0.58) | 3.00 (2.50–3.00) | - |
| | Lower momentum | 15 (55.6%) | 1.42 (0.75) | 1.67 (1.00–2.00) | 1.67 (1.11) | 2.00 (1.00–2.00) | 13.3% | 1.60 (0.99) | 2.00 (1.00–2.00) | 13.3% | 1.00 (0.65) | 1.00 (1.00–1.00) | 20.0% |
| Small island developing states SIDS (n = 32) | Higher momentum | 14 (43.8%) | 2.60 (0.78) | 2.83 (2.00–3.00) | 2.64 (0.74) | 3.00 (2.00–3.00) | - | 2.57 (0.85) | 2.50 (2.00–3.00) | - | 2.57 (0.94) | 3.00 (2.00–3.00) | - |
| | Lower momentum | 18 (56.2%) | 1.28 (0.64) | 1.00 (1.00–1.67) | 1.22 (0.88) | 1.00 (1.00–1.00) | 11.1% | 1.39 (0.78) | 1.00 (1.00–2.00) | 5.6% | 1.22 (0.55) | 1.00 (1.00–2.00) | 5.6% |
| **UNIDO Development Groups** | | | | | | | | | | | | | |
| High-income industrial economies (n = 68) | Higher momentum | 35 (51.5%) | 2.78 (0.62) | 2.67 (2.33–3.33) | 2.97 (0.66) | 3.00 (3.00–3.00) | - | 2.89 (0.72) | 3.00 (2.00–3.00) | - | 2.49 (0.92) | 2.00 (2.00–3.00) | - |
| | Lower momentum | 33 (48.5%) | 1.35 (0.58) | 1.33 (1.00–2.00) | 1.27 (0.84) | 1.00 (1.00–2.00) | 18.2% | 1.36 (0.74) | 1.00 (1.00–2.00) | 12.1% | 1.42 (0.66) | 1.00 (1.00–2.00) | 3.0% |
| High-income industrializing economies (n = 15) | Higher momentum | 9 (60.0%) | 2.70 (0.89) | 3.00 (2.00–3.67) | 2.67 (1.00) | 3.00 (2.00–3.00) | - | 2.89 (0.93) | 3.00 (2.00–4.00) | - | 2.56 (0.88) | 3.00 (2.00–3.00) | - |
| | Lower momentum | 6 (40.0%) | 1.22 (0.40) | 1.00 (1.00–1.33) | 1.00 (1.10) | 1.00 (0.00–1.00) | 33.3% | 1.83 (0.98) | 1.50 (1.00–3.00) | - | 0.83 (0.41) | 1.00 (1.00–1.00) | 16.7% |
| Middle-income industrial economies (n = 15) | Higher momentum | 12 (80.0%) | 3.06 (0.40) | 3.00 (3.00–3.17) | 3.00 (0.43) | 3.00 (3.00–3.00) | - | 3.17 (0.39) | 3.00 (3.00–3.00) | - | 3.00 (0.60) | 3.00 (3.00–3.00) | - |
| | Lower momentum | 3 (20.0%) | 1.89 (0.19) | 2.00 (1.67–2.00) | 2.33 (0.58) | 2.00 (2.00–3.00) | - | 1.67 (0.58) | 2.00 (1.00–2.00) | - | 1.67 (0.58) | 2.00 (1.00–2.00) | - |
| Middle-income industrializing economies (n = 73) | Higher momentum | 33 (45.2%) | 3.09 (0.66) | 3.00 (2.67–3.33) | 3.12 (0.70) | 3.00 (3.00–4.00) | - | 3.12 (0.78) | 3.00 (3.00–4.00) | - | 3.03 (0.88) | 3.00 (3.00–4.00) | - |
| | Lower momentum | 40 (54.8%) | 1.48 (0.69) | 1.67 (1.00–2.00) | 1.57 (1.03) | 2.00 (1.00–2.00) | 15.0% | 1.65 (0.89) | 2.00 (1.00–2.00) | 10.0% | 1.23 (0.70) | 1.00 (1.00–2.00) | 12.5% |
| Low-income economies (n = 22) | Higher momentum | 10 (45.5%) | 3.27 (0.54) | 3.17 (2.67–3.67) | 3.40 (0.52) | 3.00 (3.00–4.00) | - | 3.40 (0.70) | 3.50 (3.00–4.00) | - | 3.00 (0.67) | 3.00 (3.00–3.00) | - |
| | Lower momentum | 12 (54.5%) | 1.31 (0.74) | 1.50 (1.00–1.83) | 1.33 (0.89) | 1.00 (1.00–2.00) | 16.7% | 1.33 (1.07) | 1.00 (0.50–2.00) | 25.0% | 1.25 (0.87) | 1.00 (1.00–2.00) | 16.7% |

| | | | | | | | | | | | | | |
|---|---|---|---|---|---|---|---|---|---|---|---|---|---|
| Emerging industrial economies (n = 10) | Higher momentum | 6 (60.0%) | 3.11 (0.58) | 3.17 (2.67–3.33) | 3.00 (0.63) | 3.00 (3.00–3.00) | - | 2.83 (0.98) | 3.00 (3.00–3.00) | - | 3.50 (0.84) | 4.00 (3.00–4.00) | - |
| | Lower momentum | 4 (40.0%) | 1.50 (0.58) | 1.33 (1.17–1.83) | 2.00 (0.82) | 2.00 (1.50–2.50) | - | 1.50 (1.00) | 1.00 (1.00–2.00) | - | 1.00 (0.00) | 1.00 (1.00–1.00) | - |
| **Economic Groupings** | | | | | | | | | | | | | |
| APEC (n = 41) | Higher momentum | 24 (58.5%) | 2.72 (0.64) | 2.67 (2.17–3.17) | 2.88 (0.74) | 3.00 (2.50–3.00) | - | 2.71 (0.69) | 3.00 (2.00–3.00) | - | 2.58 (0.93) | 3.00 (2.00–3.00) | - |
| | Lower momentum | 17 (41.5%) | 1.39 (0.58) | 1.00 (1.00–2.00) | 1.29 (1.05) | 1.00 (1.00–2.00) | 23.5% | 1.35 (0.86) | 1.00 (1.00–2.00) | 11.8% | 1.53 (0.72) | 1.00 (1.00–2.00) | - |
| ASEAN (n = 9) | Higher momentum | 4 (44.4%) | 3.00 (0.72) | 2.83 (2.50–3.50) | 3.00 (0.82) | 3.00 (2.50–3.50) | - | 2.75 (1.26) | 3.00 (2.00–3.50) | - | 3.25 (0.96) | 3.50 (2.50–4.00) | - |
| | Lower momentum | 5 (55.6%) | 1.80 (0.77) | 1.67 (1.33–2.00) | 2.00 (1.58) | 2.00 (1.00–3.00) | 20.0% | 2.00 (1.00) | 2.00 (1.00–3.00) | - | 1.40 (0.55) | 1.00 (1.00–2.00) | - |
| BRICS (n = 9) | Higher momentum | 5 (55.6%) | 3.33 (0.62) | 3.33 (3.33–3.67) | 3.20 (0.84) | 3.00 (3.00–4.00) | - | 3.60 (0.55) | 4.00 (3.00–4.00) | - | 3.20 (0.84) | 3.00 (3.00–4.00) | - |
| | Lower momentum | 4 (44.4%) | 1.67 (0.61) | 1.67 (1.17–2.17) | 2.25 (0.96) | 2.50 (1.50–3.00) | - | 1.75 (0.96) | 1.50 (1.00–2.50) | - | 1.00 (0.00) | 1.00 (1.00–1.00) | - |
| CARICOM (n = 11) | Higher momentum | 6 (54.5%) | 2.44 (1.11) | 2.00 (1.67–3.67) | 2.33 (1.03) | 2.00 (2.00–3.00) | - | 2.50 (1.22) | 2.00 (2.00–4.00) | - | 2.50 (1.22) | 2.00 (2.00–4.00) | - |
| | Lower momentum | 5 (45.5%) | 0.80 (0.45) | 1.00 (1.00–1.00) | 0.80 (0.45) | 1.00 (1.00–1.00) | 20.0% | 0.80 (0.45) | 1.00 (1.00–1.00) | 20.0% | 0.80 (0.45) | 1.00 (1.00–1.00) | 20.0% |
| CIS (n = 8) | Higher momentum | 6 (75.0%) | 3.33 (0.56) | 3.17 (3.00–4.00) | 3.33 (0.52) | 3.00 (3.00–4.00) | - | 3.50 (0.55) | 3.50 (3.00–4.00) | - | 3.17 (0.75) | 3.00 (3.00–4.00) | - |
| | Lower momentum | 2 (25.0%) | 1.33 (0.47) | 1.33 (1.00–1.67) | 1.00 (0.00) | 1.00 (1.00–1.00) | - | 2.00 (1.41) | 2.00 (1.00–3.00) | - | 1.00 (0.00) | 1.00 (1.00–1.00) | - |
| ECOWAS (n = 15) | Higher momentum | 11 (73.3%) | 3.36 (0.59) | 3.33 (2.67–4.00) | 3.45 (0.52) | 3.00 (3.00–4.00) | - | 3.45 (0.69) | 4.00 (3.00–4.00) | - | 3.18 (0.75) | 3.00 (3.00–4.00) | - |
| | Lower momentum | 4 (26.7%) | 1.58 (0.42) | 1.67 (1.33–1.83) | 1.50 (0.58) | 1.50 (1.00–2.00) | - | 2.00 (0.82) | 2.00 (1.50–2.50) | - | 1.25 (0.50) | 1.00 (1.00–1.50) | - |
| EU (n = 27) | Higher momentum | 14 (51.9%) | 3.07 (0.53) | 3.00 (2.67–3.33) | 3.07 (0.47) | 3.00 (3.00–3.00) | - | 3.36 (0.50) | 3.00 (3.00–4.00) | - | 2.79 (0.89) | 3.00 (2.00–3.00) | - |
| | Lower momentum | 13 (48.1%) | 1.64 (0.46) | 1.67 (1.67–2.00) | 1.62 (0.65) | 2.00 (1.00–2.00) | 7.7% | 1.77 (0.73) | 2.00 (2.00–2.00) | 7.7% | 1.54 (0.52) | 2.00 (1.00–2.00) | - |
| G20 (n = 39) | Higher momentum | 24 (61.5%) | 2.75 (0.65) | 2.67 (2.33–3.33) | 3.00 (0.66) | 3.00 (3.00–3.00) | - | 2.79 (0.72) | 3.00 (2.00–3.00) | - | 2.46 (0.98) | 2.00 (2.00–3.00) | - |
| | Lower momentum | 15 (38.5%) | 1.56 (0.66) | 2.00 (1.00–2.00) | 1.67 (1.05) | 2.00 (1.00–3.00) | 13.3% | 1.40 (0.83) | 1.00 (1.00–2.00) | 13.3% | 1.60 (0.74) | 1.00 (1.00–2.00) | - |
| OECD (n = 59) | Higher momentum | 33 (55.9%) | 2.77 (0.58) | 2.67 (2.33–3.00) | 2.94 (0.66) | 3.00 (3.00–3.00) | - | 2.85 (0.67) | 3.00 (3.00–3.00) | - | 2.52 (0.87) | 3.00 (2.00–3.00) | - |
| | Lower momentum | 26 (44.1%) | 1.46 (0.57) | 1.50 (1.00–2.00) | 1.42 (0.86) | 1.50 (1.00–2.00) | 15.4% | 1.42 (0.76) | 1.50 (1.00–2.00) | 11.5% | 1.54 (0.65) | 1.00 (1.00–2.00) | - |

* NA = spread measurement cannot be estimated and is "not appliable" due to only one contributing program in that momentum group.
** Summaries were stratified by readiness cluster (higher vs lower) and by each country classifier; quartiles (Q1 and Q3) for inter-quartile range (IQR) were estimated using the Hyndman–Fan type 2 sample quantile definition.
*** The "higher" and "lower" momentum labels are WHO region-relative by design; they describe position relative to peers within the same WHO region rather than within each sub-region or economic groupings.
**** Abbreviations: APEC – Asia-Pacific Economic Cooperation; ASEAN – Association of Southeast Asian Nations; CARICOM – Caribbean Community; CIS – Commonwealth of Independent States; ECOWAS – Economic Community of West African States; EU – European Union; G20 – Group of 20; OECD – Organization for Economic Co-operation and Development.

**Sensitivity analysis for clustering**

We conducted multiple complementary sensitivity analyses to assess whether the observed two-profile structure (higher vs lower momentum) was robust to alternative clustering specifications and sampling variability. We re-ran within–WHO-region clustering under alternative distance and linkage choices and quantified agreement with the primary specification using the Adjusted Rand Index (ARI). ARI = 1 indicated identical partitions and values near 0 indicated chance-level agreement; negative values indicated agreement worse than expected by chance. The primary specification used Euclidean distance with Ward.D2 linkage. Alternatives included (i) Euclidean distance with average linkage; (ii) Euclidean distance with complete linkage; (iii) Gower distance (applied to ordered responses) with average linkage; and (iv) Gower distance with complete linkage (**Supplementary Table S8**).

**Supplementary Table S8.** ARI matrix for alternative clustering specifications*.

| WHO Region | Euclid + Average | Euclid + Complete | Gower + Average | Gower + Complete |
|---|---|---|---|---|
| AFRO | −0.020 | 0.452 | −0.020 | 0.662 |
| AMRO | 0.919 | 0.769 | 0.075 | −0.074 |
| EMRO | 1.000 | 1.000 | 1.000 | 1.000 |
| EURO | 1.000 | 1.000 | 1.000 | 0.915 |
| SEARO | 1.000 | 1.000 | −0.141 | −0.141 |
| WPRO | 1.000 | 1.000 | 1.000 | 1.000 |

* Cells are shaded using pre-specified interpretive thresholds (note that ARI has no universal cut-offs); green = very high agreement (ARI ≥ 0.90); blue = broadly consistent (ARI 0.70–0.89); pink = low agreement (ARI < 0.70).

Agreement with the primary specification was high across most regions and Euclidean-linkage variants (ARI ~ 1.00), whereas agreement decreased under Gower distance in AFRO, AMRO, and SEARO, indicating sensitivity to distance metric choice in some regions. To further validate our primary findings, we first ran a robustness check to assess whether the region-relative higher vs lower momentum profiles were robust to treating the 0–4 domain scores as ordinal (with Gower) rather than numeric (with Euclidean).

Within each WHO region, we re-estimated the k = 2 hierarchical clustering using an ordinal-aware Gower dissimilarity (with ordered factors) with average and complete linkage and compared the resulting higher–lower separation to the primary Euclidean+Ward.D2 specification. We evaluated the region-level Higher–Lower momentum differences (Δ) in composite and domain means. We observed that momentum mean differences were directionally consistent under ordinal-aware Gower clustering for five regions, indicating that the two-profile separation was robust to the distance metric (**Supplementary Table S9**). SEARO showed sensitivity consistent with small sample size and a single high-surveillance outlier, producing minor sign changes without materially affecting the composite separation.

**Supplementary Table S9.** Sensitivity checks for directional preservation under alternative clustering specifications stratified by WHO regions*.

| WHO Region | n (higher) | n (lower) | Δ Composite | Δ Clinical Management | Δ Diagnostics | Δ Surveillance |
|---|---|---|---|---|---|---|
| **AFRO** | | | | | | |
| Euclidean + Ward.D2 (primary) | 19 | 25 | 1.77 | 1.81 | 1.69 | 1.83 |
| Gower (ordered) + average | 41 | 3 | 2.44 | 2.51 | 2.59 | 2.22 |
| Gower (ordered) + complete | 21 | 23 | 1.73 | 2.08 | 1.68 | 1.42 |
| **AMRO** | | | | | | |
| Euclidean + Ward.D2 (primary) | 34 | 16 | 1.70 | 1.99 | 1.68 | 1.42 |
| Gower (ordered) + average | 17 | 33 | 1.68 | 1.63 | 1.75 | 1.67 |
| Gower (ordered) + complete | 6 | 44 | 1.85 | 1.73 | 1.80 | 2.01 |

| | | | | | | |
|---|---|---|---|---|---|---|
| **EMRO** | | | | | | |
| Euclidean + Ward.D2 (primary) | 8 | 7 | 2.23 | 1.79 | 2.50 | 2.39 |
| Gower (ordered) + average | 8 | 7 | 2.23 | 1.79 | 2.50 | 2.39 |
| Gower (ordered) + complete | 8 | 7 | 2.23 | 1.79 | 2.50 | 2.39 |
| **EURO** | | | | | | |
| Euclidean + Ward.D2 (primary) | 26 | 21 | 1.59 | 1.76 | 1.59 | 1.41 |
| Gower (ordered) + average | 26 | 21 | 1.59 | 1.76 | 1.59 | 1.41 |
| Gower (ordered) + complete | 25 | 22 | 1.58 | 1.70 | 1.55 | 1.51 |
| **SEARO** | | | | | | |
| Euclidean + Ward.D2 (primary) | 1 | 8 | 0.33 | −0.50 | −1.00 | 2.50 |
| Gower (ordered) + average | 2 | 7 | 0.81 | 1.36 | 1.43 | −0.36 |
| Gower (ordered) + complete | 2 | 7 | 0.81 | 1.36 | 1.43 | −0.36 |
| **WPRO** | | | | | | |
| Euclidean + Ward.D2 (primary) | 11 | 17 | 1.59 | 1.83 | 1.50 | 1.44 |
| Gower (ordered) + average | 11 | 17 | 1.59 | 1.83 | 1.50 | 1.44 |
| Gower (ordered) + complete | 11 | 17 | 1.59 | 1.83 | 1.50 | 1.44 |

* Delta (Δ) was calculated by subtracting the lower momentum group mean from higher momentum group mean under the respective clustering specification. A positive value indicates that higher momentum group had a mean score higher than the lower momentum cluster (*ideal result*).

Next, we also checked whether the domain gradient (structural preservation), i.e., whether large surveillance deficits and higher urgency for diagnostics, is largely preserved with Gower distance. Indeed, the gradient structure was highly preserved across all WHO regions (**Supplementary Table S10**).

**Supplementary Table S10.** Descriptive summary for relative region-defined momentum scores under alternative clustering specifications, stratified by WHO regions.

| WHO Region | Clinical Management | | Diagnostics | | Surveillance | |
|---|---|---|---|---|---|---|
| | Higher momentum | Lower momentum | Higher momentum | Lower momentum | Higher momentum | Lower momentum |
| **AFRO** | | | | | | |
| Euclidean + Ward.D2 (primary) | 3.37 (0.60) | 1.56 (1.04) | 3.37 (0.60) | 1.68 (0.95) | 3.11 (0.74) | 1.28 (0.84) |
| Gower (ordered) + average | 2.51 (1.12) | 0.00 (0.00) | 2.59 (1.00) | 0.00 (0.00) | 2.22 (1.11) | 0.00 (0.00) |
| Gower (ordered) + complete | 3.43 (0.51) | 1.35 (0.83) | 3.29 (0.64) | 1.61 (0.94) | 2.81 (1.12) | 1.39 (0.84) |
| **AMRO** | | | | | | |
| Euclidean + Ward.D2 (primary) | 2.62 (0.74) | 0.62 (0.50) | 2.62 (0.78) | 0.94 (0.77) | 2.35 (0.95) | 0.94 (0.68) |
| Gower (ordered) + average | 3.06 (0.56) | 1.42 (0.97) | 3.24 (0.44) | 1.48 (0.83) | 3.00 (0.71) | 1.33 (0.78) |
| Gower (ordered) + complete | 3.50 (0.55) | 1.77 (1.05) | 3.67 (0.52) | 1.86 (0.98) | 3.67 (0.52) | 1.66 (0.91) |
| **EMRO** | | | | | | |
| Euclidean + Ward.D2 (primary) | 3.50 (0.53) | 1.71 (1.25) | 3.50 (0.76) | 1.00 (1.00) | 3.25 (0.46) | 0.86 (0.69) |
| Gower (ordered) + average | 3.50 (0.53) | 1.71 (1.25) | 3.50 (0.76) | 1.00 (1.00) | 3.25 (0.46) | 0.86 (0.69) |
| Gower (ordered) + complete | 3.50 (0.53) | 1.71 (1.25) | 3.50 (0.76) | 1.00 (1.00) | 3.25 (0.46) | 0.86 (0.69) |
| **EURO** | | | | | | |
| Euclidean + Ward.D2 (primary) | 3.19 (0.49) | 1.43 (0.68) | 3.31 (0.47) | 1.71 (0.72) | 2.88 (0.82) | 1.48 (0.51) |
| Gower (ordered) + average | 3.19 (0.49) | 1.43 (0.68) | 3.31 (0.47) | 1.71 (0.72) | 2.88 (0.82) | 1.48 (0.51) |
| Gower (ordered) + complete | 3.20 (0.50) | 1.50 (0.74) | 3.32 (0.48) | 1.77 (0.75) | 2.96 (0.73) | 1.45 (0.51) |
| **SEARO** | | | | | | |
| Euclidean + Ward.D2 (primary) | 2.00 (NA) | 2.50 (0.76) | 1.00 (NA) | 2.00 (0.76) | 4.00 (NA) | 1.50 (0.53) |
| Gower (ordered) + average | 3.50 (0.71) | 2.14 (0.38) | 3.00 (0.00) | 1.57 (0.53) | 1.50 (0.71) | 1.86 (1.07) |
| Gower (ordered) + complete | 3.50 (0.71) | 2.14 (0.38) | 3.00 (0.00) | 1.57 (0.53) | 1.50 (0.71) | 1.86 (1.07) |
| **WPRO** | | | | | | |
| Euclidean + Ward.D2 (primary) | 3.18 (0.40) | 1.35 (0.86) | 3.09 (0.30) | 1.59 (0.71) | 2.91 (0.70) | 1.47 (0.62) |
| Gower (ordered) + average | 3.18 (0.40) | 1.35 (0.86) | 3.09 (0.30) | 1.59 (0.71) | 2.91 (0.70) | 1.47 (0.62) |
| Gower (ordered) + complete | 3.18 (0.40) | 1.35 (0.86) | 3.09 (0.30) | 1.59 (0.71) | 2.91 (0.70) | 1.47 (0.62) |

Furthermore, to evaluate whether the proposed two-cluster classification within each WHO region was stable, we performed bootstrap re-sampling (seed = 1; B = 500) within each region. Cluster stability was quantified using the mean Jaccard similarity for each cluster across bootstrap samples. As a rule of thumb, Jaccard > 0.75 indicates stable clusters. Bootstrap resampling indicated high within-region cluster stability (mean Jaccard ~ 0.82 to ~ 0.99 across regions; **Supplementary Table S11**), supporting reproducibility of the two-profile classification. Stability was symmetric in all regions; in SEARO (n = 9), one cluster showed lower stability, consistent with expectations given the inherent small sample size of the region.

**Supplementary Table S11.** Mean Jaccard scores within each region.

| WHO Region | n | Jaccard_1 * | Jaccard_2** | Mean Jaccard |
|---|---|---|---|---|
| AFRO | 44 | 0.843 | 0.803 | 0.823 |
| AMRO | 50 | 0.835 | 0.867 | 0.851 |
| EMRO | 15 | 0.965 | 0.971 | 0.968 |
| EURO | 47 | 0.987 | 0.986 | 0.987 |
| SEARO | 9 | 0.902 | 0.763 | 0.833 |
| WPRO | 28 | 0.874 | 0.863 | 0.869 |

* Jaccard_1 represents lower momentum cluster.

** Jaccard_2 represents higher momentum cluster.

**Sensitivity analysis for Likert coding**

Responses to the three urgency domains (clinical management, diagnostics, surveillance) were mapped to a 0–4 ordinal scale (0 = *Not aware*, 1 = *Absent*, 2 = *Low*, 3 = *Moderate*, 4 = *High*). Because "Not aware" and "Absent" represent distinct low states (low salience vs explicitly not on the agenda), we treated them as independent ordered categories in the primary analysis. Importantly, we treated 'not aware' as the minimum urgency state (0), and 'absent' as a slightly higher but still minimal urgency judgement (1), preserving the ordinal ordering from no salience to high momentum. To assess whether the region-relative clustering results were sensitive to this low-end scale coding, we conducted two separate stress tests:

1. **Collapse 0 and 1**: recoded both "Not aware" (0) and "Absent" (1) to a single lowest category (0), then re-ran the within-region Euclidean+Ward.D2 clustering (k = 2) on the recoded domain responses.

2. **Exclude any 'Not aware' responses**: removed program observations with any domain response of 0 ("Not aware") across the three domains, then re-ran within-region clustering on the remaining subset. This test evaluated whether the region-relative profiles are primarily driven by "Not aware" responses in settings where these are common.

In both stress tests, we compared cluster assignments to the primary specification using (i) percent agreement and (ii) Adjusted Rand Index (ARI) within each WHO region, restricting comparisons to overlapping program observations. Collapsing 0 ("Not aware") and 1 ("Absent") yielded near-identical cluster assignments in most WHO regions (agreement 96% to 100% in AFRO, AMRO, EMRO, EURO; ARI 0.84 to 1.00), indicating that the primary region-relative profile structure did not depend on distinguishing between these two lowest response categories. Lower agreement was observed in SEARO and WPRO; however, SEARO had a very small sample (n = 9), and minor changes can substantially affect two-cluster assignments in small strata (**Supplementary Table S12**).

**Supplementary Table S12.** ARI matrix for under different sensitivity analyses.

| Scenario | WHO region | n (overlap) | Agreement (%) | ARI |
|---|---|---|---|---|
| Collapse 0/1 (Not aware + Absent → 0) | AFRO | 44 | 100.0 | 1.000 |
| | AMRO | 50 | 96.0 | 0.841 |
| | EMRO | 15 | 100.0 | 1.000 |
| | EURO | 47 | 97.9 | 0.915 |
| | SEARO | 9 | 22.2 | 0.222 |
| | WPRO | 28 | 82.1 | 0.392 |
| Exclude any 'Not aware' (drop any 0) | AFRO | 39 | 94.9 | 0.800 |
| | AMRO | 44 | 50.0 | -0.080 |
| | EMRO | 12 | 100.0 | 1.000 |
| | EURO | 45 | 100.0 | 1.000 |
| | SEARO | 9 | 100.0 | 1.000 |
| | WPRO | 26 | 100.0 | 1.000 |

Excluding program observations with any "Not aware" response also produced identical assignments in EMRO, EURO, SEARO, and WPRO (agreement 100%; ARI 1.00), and high agreement in AFRO (94.9%; ARI 0.80). In AMRO, agreement was lower, consistent with "Not aware" responses representing an important axis of separation in this region; removing these responses changed the underlying population being clustered (i.e., restricting to "aware-only" programs). Importantly, across stress tests, the substantive interpretation was preserved: within each region, the higher profile retained higher composite mean and domain means than the lower profile. Surveillance continued to the most significant bottleneck (**Supplementary Table S13**).

**Supplementary Table S13.** Descriptive summary for relative region-defined momentum scores under different sensitivity analyses, stratified by WHO regions.

| Scenario | WHO region | Relative Profile | n | Composite mean (SD) | Clinical mean (SD) | Diagnostics mean (SD) | Surveillance mean (SD) |
|---|---|---|---|---|---|---|---|
| Collapse 0/1 (Not aware + Absent → 0) | AFRO | Higher | 19 | 3.28 (0.55) | 3.37 (0.60) | 3.37 (0.60) | 3.11 (0.74) |
| | | Lower | 25 | 1.15 (0.90) | 1.24 (1.30) | 1.40 (1.22) | 0.80 (1.12) |
| | AMRO | Higher | 34 | 2.43 (0.80) | 2.56 (0.89) | 2.50 (1.02) | 2.24 (1.23) |
| | | Lower | 16 | 0.15 (0.32) | 0.00 (0.00) | 0.44 (0.96) | 0.00 (0.00) |
| | EMRO | Higher | 8 | 3.42 (0.50) | 3.50 (0.53) | 3.50 (0.76) | 3.25 (0.46) |
| | | Lower | 7 | 0.95 (0.73) | 1.71 (1.25) | 0.86 (1.07) | 0.29 (0.76) |
| | EURO | Higher | 25 | 3.16 (0.46) | 3.20 (0.50) | 3.32 (0.48) | 2.96 (0.73) |
| | | Lower | 22 | 1.18 (0.79) | 1.14 (1.08) | 1.50 (1.10) | 0.91 (1.02) |
| | SEARO | Higher | 6 | 2.06 (0.53) | 2.50 (0.84) | 2.33 (0.52) | 1.33 (1.03) |
| | | Lower | 3 | 1.22 (0.69) | 2.33 (0.58) | 0.00 (0.00) | 1.33 (2.31) |
| | WPRO | Higher | 16 | 2.75 (0.59) | 2.94 (0.57) | 2.62 (0.89) | 2.69 (0.70) |
| | | Lower | 12 | 0.44 (0.67) | 0.17 (0.58) | 0.83 (1.27) | 0.33 (0.78) |
| Exclude any 'Not aware' (drop any 0) | AFRO | Higher | 21 | 3.17 (0.62) | 3.33 (0.58) | 3.29 (0.64) | 2.90 (0.94) |
| | | Lower | 18 | 1.63 (0.46) | 1.61 (0.61) | 1.78 (0.73) | 1.50 (0.62) |
| | AMRO | Higher | 12 | 3.31 (0.36) | 3.25 (0.45) | 3.33 (0.49) | 3.33 (0.49) |
| | | Lower | 32 | 1.76 (0.60) | 1.88 (0.79) | 1.84 (0.77) | 1.56 (0.67) |
| | EMRO | Higher | 8 | 3.42 (0.50) | 3.50 (0.53) | 3.50 (0.76) | 3.25 (0.46) |
| | | Lower | 4 | 1.67 (0.27) | 2.25 (0.50) | 1.75 (0.50) | 1.00 (0.00) |
| | EURO | Higher | 26 | 3.13 (0.48) | 3.19 (0.49) | 3.31 (0.47) | 2.88 (0.82) |
| | | Lower | 19 | 1.61 (0.39) | 1.58 (0.51) | 1.79 (0.63) | 1.47 (0.51) |
| | SEARO | Higher | 1 | 2.33 (NA) | 2.00 (NA) | 1.00 (NA) | 4.00 (NA) |
| | | Lower | 8 | 2.00 (0.50) | 2.50 (0.76) | 2.00 (0.76) | 1.50 (0.53) |
| | WPRO | Higher | 11 | 3.06 (0.39) | 3.18 (0.40) | 3.09 (0.30) | 2.91 (0.70) |
| | | Lower | 15 | 1.47 (0.55) | 1.53 (0.74) | 1.40 (0.51) | 1.47 (0.64) |

**Supplementary Table S14.** Crosstabulation for group membership under higher and lower momentum profiles by respondent type.

| WHO region | Respondent | Higher momentum, n (%) | Lower momentum, n (%) | Total | Fisher P (within region) |
|---|---|---|---|---|---|
| **AFRO** | MoH | 0 (0.0%) | 1 (100.0%) | 1 | 0.783 |
| | NRL | 1 (25.0%) | 3 (75.0%) | 4 | |
| | NTP | 18 (46.2%) | 21 (53.8%) | 39 | |
| **AMRO** | Expert | 5 (62.5%) | 3 (37.5%) | 8 | 0.490 |
| | MoH | 4 (50.0%) | 4 (50.0%) | 8 | |
| | NRL | 3 (60.0%) | 2 (40.0%) | 5 | |
| | NTP | 22 (75.9%) | 7 (24.1%) | 29 | |
| **EMRO** | Expert | 1 (100.0%) | 0 (0.0%) | 1 | 0.369 |
| | MoH | 2 (100.0%) | 0 (0.0%) | 2 | |
| | NRL | 1 (25.0%) | 3 (75.0%) | 4 | |
| | NTP | 4 (50.0%) | 4 (50.0%) | 8 | |
| **EURO** | Expert | 0 (0.0%) | 5 (100.0%) | 5 | 0.065 |
| | MoH | 3 (50.0%) | 3 (50.0%) | 6 | |
| | NRL | 8 (61.5%) | 5 (38.5%) | 13 | |
| | NTP | 15 (65.2%) | 8 (34.8%) | 23 | |
| **SEARO** | NTP | 1 (11.1%) | 8 (88.9%) | 9 | NA |
| **WPRO** | Expert | 1 (50.0%) | 1 (50.0%) | 2 | 0.474 |
| | MoH | 3 (75.0%) | 1 (25.0%) | 4 | |
| | NRL | 1 (20.0%) | 4 (80.0%) | 5 | |
| | NTP | 6 (35.3%) | 11 (64.7%) | 17 | |
| Combined P value across regions (Fisher's method): 0.369 | | | | | |

**Supplementary Table S15.** Crosstabulation for group membership under higher and lower momentum profiles by TB burden and SDI.

| WHO region | log1p(TB) median (Q1–Q3), Higher | log1p(TB) median (Q1–Q3), Lower | Wilcoxon P | SDI median (Q1–Q3), Higher | SDI median (Q1–Q3), Lower | Wilcoxon P |
|---|---|---|---|---|---|---|
| **AFRO** | 5.18 (4.47–5.77) | 4.96 (4.48–5.42) | 0.570 | 0.461 (0.342–0.562) | 0.487 (0.406–0.578) | 0.286 |
| **AMRO** | 2.28 (1.44–3.91) | 1.97 (1.44–3.68) | 0.637 | 0.755 (0.686–0.883) | 0.759 (0.685–0.877) | 0.909 |
| **EMRO** | 2.87 (1.99–3.68) | 4.16 (2.40–5.32) | 0.118 | 0.722 (0.613–0.820) | 0.684 (0.334–0.770) | 0.272 |
| **EURO** | 2.43 (1.79–3.83) | 2.03 (1.77–2.77) | 0.266 | 0.799 (0.758–0.856) | 0.847 (0.789–0.889) | 0.140 |
| **SEARO** | 6.18 (6.18–6.18) | 5.25 (4.54–5.42) | 0.333 | 0.532 (0.532–0.532) | 0.566 (0.501–0.674) | 1.000 |
| **WPRO** | 3.58 (2.07–5.21) | 3.81 (2.19–4.65) | 0.794 | 0.770 (0.666–0.853) | 0.832 (0.669–0.853) | 0.887 |
| Combined P values across regions (Fisher's method): 0.477 | | | | Combined P values across regions (Fisher's method): 0.662 | | |

**Supplementary Table S16.** Crosstabulation for group membership under higher and lower momentum profiles by NTM institutionalisation status (collapsed version).

| WHO Region | Institutionalisation (2×2) * | Higher momentum, n (%) | Lower momentum, n (%) | Total | OR (Higher momentum) | 95% CI (exact) | Fisher P |
|---|---|---|---|---|---|---|---|
| **AFRO** | Not under NTP | 7 (25.9%) | 20 (74.1%) | 27 | Ref | Ref | Ref |
| | NTP integrated | 12 (70.6%) | 5 (29.4%) | 17 | 6.515 | 1.50–33.38 | 0.005 |
| **AMRO** | Not under NTP | 19 (55.9%) | 15 (44.1%) | 34 | Ref | Ref | Ref |
| | NTP integrated | 15 (93.8%) | 1 (6.2%) | 16 | 11.376 | 1.43–529.62 | 0.009 |
| **EMRO** | Not under NTP | 2 (33.3%) | 4 (66.7%) | 6 | Ref | Ref | Ref |
| | NTP integrated | 6 (66.7%) | 3 (33.3%) | 9 | 3.619 | 0.30–64.06 | 0.315 |
| **EURO** | Not under NTP | 17 (54.8%) | 14 (45.2%) | 31 | Ref | Ref | Ref |
| | NTP integrated | 9 (56.2%) | 7 (43.8%) | 16 | 1.058 | 0.27–4.29 | 1.000 |
| **SEARO** | Not under NTP | 1 (25.0%) | 3 (75.0%) | 4 | Ref | Ref | Ref |
| | NTP integrated | 0 (0.0%) | 5 (100.0%) | 5 | 0.000 | 0.00–31.20 | 0.444 |
| **WPRO** | Not under NTP | 8 (32.0%) | 17 (68.0%) | 25 | Ref | Ref | Ref |
| | NTP integrated | 3 (100.0%) | 0 (0.0%) | 3 | Inf | 0.70–Inf | 0.050 |
| Combined p-value across regions (Fisher's method on region-specific Fisher p-values): 0.003 | | | | | | | |

* For 2×2 institutionalisation grouping, "NTP integrated" includes both pulmonary only and all NTM responses; "Not under NTP" includes orphaned, intent (NTP interest to expand portfolio), and institutionalised outside NTP.

**Supplementary Table S17.** Stakeholder-ranked barriers and enablers for translating NTM discussions and policy urgency into implementable program action, presented as n and % per rank. Note that for NTP integrated, there were eight themes to rank while for intent (within NTP) and orphaned, there were seven themes to rank. The additional "competing TB priorities" was only available to the NTP integrated respondents.

| Level | Institutionalisation | Barrier/Enabler | Rank 1 | Rank 2 | Rank 3 | Rank 4 | Rank 5 | Rank 6 | Rank 7 | Rank 8 |
|---|---|---|---|---|---|---|---|---|---|---|
| Global (Overall) | NTP integrated (n = 66) | Burden/incidence data | 27 (40.9%) | 6 (9.1%) | 10 (15.2%) | 4 (6.1%) | 5 (7.6%) | 5 (7.6%) | 2 (3.0%) | 7 (10.6%) |
| | | Clinical awareness among physicians | 5 (7.6%) | 14 (21.2%) | 14 (21.2%) | 8 (12.1%) | 9 (13.6%) | 5 (7.6%) | 7 (10.6%) | 4 (6.1%) |
| | | Competing TB priorities | 8 (12.1%) | 11 (16.7%) | 6 (9.1%) | 7 (10.6%) | 10 (15.2%) | 12 (18.2%) | 8 (12.1%) | 4 (6.1%) |
| | | Dedicated funding/budget | 12 (18.2%) | 10 (15.2%) | 17 (25.8%) | 8 (12.1%) | 9 (13.6%) | 5 (7.6%) | 2 (3.0%) | 3 (4.5%) |
| | | Political/institutional barriers | 3 (4.5%) | 2 (3.0%) | 2 (3.0%) | 8 (12.1%) | 4 (6.1%) | 11 (16.7%) | 18 (27.3%) | 18 (27.3%) |
| | | Public health interest/urgency | 3 (4.5%) | 14 (21.2%) | 6 (9.1%) | 13 (19.7%) | 12 (18.2%) | 7 (10.6%) | 8 (12.1%) | 3 (4.5%) |
| | | Stakeholder demand | 3 (4.5%) | 3 (4.5%) | 7 (10.6%) | 11 (16.7%) | 7 (10.6%) | 9 (13.6%) | 11 (16.7%) | 15 (22.7%) |
| | | Technical/lab expertise | 5 (7.6%) | 6 (9.1%) | 4 (6.1%) | 7 (10.6%) | 10 (15.2%) | 12 (18.2%) | 10 (15.2%) | 12 (18.2%) |
| | Intent (within NTP) (n = 72) | Burden/incidence data | 20 (27.8%) | 13 (18.1%) | 14 (19.4%) | 9 (12.5%) | 10 (13.9%) | 4 (5.6%) | 2 (2.8%) | |
| | | Clinical awareness among physicians | 8 (11.1%) | 9 (12.5%) | 14 (19.4%) | 16 (22.2%) | 13 (18.1%) | 6 (8.3%) | 6 (8.3%) | |
| | | Dedicated funding/budget | 17 (23.6%) | 17 (23.6%) | 11 (15.3%) | 11 (15.3%) | 11 (15.3%) | 5 (6.9%) | 0 (0.0%) | |
| | | Political/institutional barriers | 3 (4.2%) | 1 (1.4%) | 6 (8.3%) | 5 (6.9%) | 10 (13.9%) | 16 (22.2%) | 31 (43.1%) | |
| | | Public health interest/urgency | 13 (18.1%) | 10 (13.9%) | 9 (12.5%) | 13 (18.1%) | 11 (15.3%) | 9 (12.5%) | 7 (9.7%) | |
| | | Stakeholder demand | 0 (0.0%) | 3 (4.2%) | 9 (12.5%) | 9 (12.5%) | 8 (11.1%) | 23 (31.9%) | 20 (27.8%) | |
| | | Technical/lab expertise | 11 (15.3%) | 19 (26.4%) | 9 (12.5%) | 9 (12.5%) | 9 (12.5%) | 9 (12.5%) | 6 (8.3%) | |
| | Orphaned (n = 25) | Burden/incidence data | 8 (32.0%) | 2 (8.0%) | 7 (28.0%) | 1 (4.0%) | 4 (16.0%) | 1 (4.0%) | 2 (8.0%) | |
| | | Clinical awareness among physicians | 4 (16.0%) | 2 (8.0%) | 6 (24.0%) | 2 (8.0%) | 5 (20.0%) | 3 (12.0%) | 3 (12.0%) | |
| | | Dedicated funding/budget | 7 (28.0%) | 4 (16.0%) | 4 (16.0%) | 2 (8.0%) | 0 (0.0%) | 6 (24.0%) | 2 (8.0%) | |
| | | Political/institutional barriers | 0 (0.0%) | 1 (4.0%) | 2 (8.0%) | 3 (12.0%) | 2 (8.0%) | 6 (24.0%) | 11 (44.0%) | |
| | | Public health interest/urgency | 3 (12.0%) | 6 (24.0%) | 2 (8.0%) | 9 (36.0%) | 4 (16.0%) | 0 (0.0%) | 1 (4.0%) | |
| | | Stakeholder demand | 1 (4.0%) | 5 (20.0%) | 2 (8.0%) | 6 (24.0%) | 3 (12.0%) | 6 (24.0%) | 2 (8.0%) | |
| | | Technical/lab expertise | 2 (8.0%) | 5 (20.0%) | 2 (8.0%) | 2 (8.0%) | 7 (28.0%) | 3 (12.0%) | 4 (16.0%) | |
| Higher within-region (relative) momentum | NTP integrated (n = 66) | Burden / incidence data | 16 (35.6%) | 4 (8.9%) | 6 (13.3%) | 3 (6.7%) | 5 (11.1%) | 4 (8.9%) | 1 (2.2%) | 6 (13.3%) |
| | | Clinical awareness among physicians | 4 (8.9%) | 10 (22.2%) | 12 (26.7%) | 4 (8.9%) | 6 (13.3%) | 3 (6.7%) | 4 (8.9%) | 2 (4.4%) |
| | | Competing TB priorities | 7 (15.6%) | 7 (15.6%) | 2 (4.4%) | 6 (13.3%) | 9 (20.0%) | 7 (15.6%) | 5 (11.1%) | 2 (4.4%) |
| | | Dedicated funding / budget | 10 (22.2%) | 7 (15.6%) | 13 (28.9%) | 2 (4.4%) | 6 (13.3%) | 3 (6.7%) | 2 (4.4%) | 2 (4.4%) |
| | | Political / institutional barriers | 2 (4.4%) | 1 (2.2%) | 1 (2.2%) | 6 (13.3%) | 3 (6.7%) | 7 (15.6%) | 14 (31.1%) | 11 (24.4%) |
| | | Public health interest / urgency | 2 (4.4%) | 10 (22.2%) | 3 (6.7%) | 10 (22.2%) | 7 (15.6%) | 6 (13.3%) | 5 (11.1%) | 2 (4.4%) |
| | | Stakeholder demand | 1 (2.2%) | 3 (6.7%) | 5 (11.1%) | 10 (22.2%) | 4 (8.9%) | 5 (11.1%) | 6 (13.3%) | 11 (24.4%) |
| | | Technical / lab expertise | 3 (6.7%) | 3 (6.7%) | 3 (6.7%) | 4 (8.9%) | 5 (11.1%) | 10 (22.2%) | 8 (17.8%) | 9 (20.0%) |
| | Intent (within NTP) (n = 27) | Burden / incidence data | 8 (29.6%) | 6 (22.2%) | 3 (11.1%) | 2 (7.4%) | 5 (18.5%) | 2 (7.4%) | 1 (3.7%) | |
| | | Clinical awareness among physicians | 1 (3.7%) | 3 (11.1%) | 5 (18.5%) | 5 (18.5%) | 6 (22.2%) | 4 (14.8%) | 3 (11.1%) | |
| | | Dedicated funding / budget | 8 (29.6%) | 6 (22.2%) | 8 (29.6%) | 2 (7.4%) | 2 (7.4%) | 1 (3.7%) | 0 (0.0%) | |

| | | | | | | | | | | |
|---|---|---|---|---|---|---|---|---|---|---|
| | | Political / institutional barriers | 2 (7.4%) | 0 (0.0%) | 3 (11.1%) | 3 (11.1%) | 3 (11.1%) | 8 (29.6%) | 8 (29.6%) | |
| | | Public health interest / urgency | 4 (14.8%) | 7 (25.9%) | 3 (11.1%) | 5 (18.5%) | 5 (18.5%) | 1 (3.7%) | 2 (7.4%) | |
| | | Stakeholder demand | 0 (0.0%) | 0 (0.0%) | 2 (7.4%) | 4 (14.8%) | 5 (18.5%) | 7 (25.9%) | 9 (33.3%) | |
| | | Technical / lab expertise | 4 (14.8%) | 5 (18.5%) | 3 (11.1%) | 6 (22.2%) | 1 (3.7%) | 4 (14.8%) | 4 (14.8%) | |
| | Orphaned (n = 6) | Burden / incidence data | 1 (16.7%) | 1 (16.7%) | 2 (33.3%) | 0 (0.0%) | 2 (33.3%) | 0 (0.0%) | 0 (0.0%) | |
| | | Clinical awareness among physicians | 1 (16.7%) | 0 (0.0%) | 0 (0.0%) | 1 (16.7%) | 1 (16.7%) | 1 (16.7%) | 2 (33.3%) | |
| | | Dedicated funding / budget | 4 (66.7%) | 0 (0.0%) | 1 (16.7%) | 0 (0.0%) | 0 (0.0%) | 1 (16.7%) | 0 (0.0%) | |
| | | Political / institutional barriers | 0 (0.0%) | 1 (16.7%) | 0 (0.0%) | 0 (0.0%) | 0 (0.0%) | 2 (33.3%) | 3 (50.0%) | |
| | | Public health interest / urgency | 0 (0.0%) | 2 (33.3%) | 1 (16.7%) | 2 (33.3%) | 1 (16.7%) | 0 (0.0%) | 0 (0.0%) | |
| | | Stakeholder demand | 0 (0.0%) | 0 (0.0%) | 1 (16.7%) | 1 (16.7%) | 1 (16.7%) | 2 (33.3%) | 1 (16.7%) | |
| | | Technical / lab expertise | 0 (0.0%) | 2 (33.3%) | 1 (16.7%) | 2 (33.3%) | 1 (16.7%) | 0 (0.0%) | 0 (0.0%) | |
| **Lower within-region (relative) momentum** | NTP integrated (n = 66) | Burden / incidence data | 11 (52.4%) | 2 (9.5%) | 4 (19.0%) | 1 (4.8%) | 0 (0.0%) | 1 (4.8%) | 1 (4.8%) | 1 (4.8%) |
| | | Clinical awareness among physicians | 1 (4.8%) | 4 (19.0%) | 2 (9.5%) | 4 (19.0%) | 3 (14.3%) | 2 (9.5%) | 3 (14.3%) | 2 (9.5%) |
| | | Competing TB priorities | 1 (4.8%) | 4 (19.0%) | 4 (19.0%) | 1 (4.8%) | 1 (4.8%) | 5 (23.8%) | 3 (14.3%) | 2 (9.5%) |
| | | Dedicated funding / budget | 2 (9.5%) | 3 (14.3%) | 4 (19.0%) | 6 (28.6%) | 3 (14.3%) | 2 (9.5%) | 0 (0.0%) | 1 (4.8%) |
| | | Political / institutional barriers | 1 (4.8%) | 1 (4.8%) | 1 (4.8%) | 2 (9.5%) | 1 (4.8%) | 4 (19.0%) | 4 (19.0%) | 7 (33.3%) |
| | | Public health interest / urgency | 1 (4.8%) | 4 (19.0%) | 3 (14.3%) | 3 (14.3%) | 5 (23.8%) | 1 (4.8%) | 3 (14.3%) | 1 (4.8%) |
| | | Stakeholder demand | 2 (9.5%) | 0 (0.0%) | 2 (9.5%) | 1 (4.8%) | 3 (14.3%) | 4 (19.0%) | 5 (23.8%) | 4 (19.0%) |
| | | Technical / lab expertise | 2 (9.5%) | 3 (14.3%) | 1 (4.8%) | 3 (14.3%) | 5 (23.8%) | 2 (9.5%) | 2 (9.5%) | 3 (14.3%) |
| | Intent (within NTP) (n = 45) | Burden / incidence data | 12 (26.7%) | 7 (15.6%) | 11 (24.4%) | 7 (15.6%) | 5 (11.1%) | 2 (4.4%) | 1 (2.2%) | |
| | | Clinical awareness among physicians | 7 (15.6%) | 6 (13.3%) | 9 (20.0%) | 11 (24.4%) | 7 (15.6%) | 2 (4.4%) | 3 (6.7%) | |
| | | Dedicated funding / budget | 9 (20.0%) | 11 (24.4%) | 3 (6.7%) | 9 (20.0%) | 9 (20.0%) | 4 (8.9%) | 0 (0.0%) | |
| | | Political / institutional barriers | 1 (2.2%) | 1 (2.2%) | 3 (6.7%) | 2 (4.4%) | 7 (15.6%) | 8 (17.8%) | 23 (51.1%) | |
| | | Public health interest / urgency | 9 (20.0%) | 3 (6.7%) | 6 (13.3%) | 8 (17.8%) | 6 (13.3%) | 8 (17.8%) | 5 (11.1%) | |
| | | Stakeholder demand | 0 (0.0%) | 3 (6.7%) | 7 (15.6%) | 5 (11.1%) | 3 (6.7%) | 16 (35.6%) | 11 (24.4%) | |
| | | Technical / lab expertise | 7 (15.6%) | 14 (31.1%) | 6 (13.3%) | 3 (6.7%) | 8 (17.8%) | 5 (11.1%) | 2 (4.4%) | |
| | Orphaned (n = 19) | Burden / incidence data | 7 (36.8%) | 1 (5.3%) | 5 (26.3%) | 1 (5.3%) | 2 (10.5%) | 1 (5.3%) | 2 (10.5%) | |
| | | Clinical awareness among physicians | 3 (15.8%) | 2 (10.5%) | 6 (31.6%) | 1 (5.3%) | 4 (21.1%) | 2 (10.5%) | 1 (5.3%) | |
| | | Dedicated funding / budget | 3 (15.8%) | 4 (21.1%) | 3 (15.8%) | 2 (10.5%) | 0 (0.0%) | 5 (26.3%) | 2 (10.5%) | |
| | | Political / institutional barriers | 0 (0.0%) | 0 (0.0%) | 2 (10.5%) | 3 (15.8%) | 2 (10.5%) | 4 (21.1%) | 8 (42.1%) | |
| | | Public health interest / urgency | 3 (15.8%) | 4 (21.1%) | 1 (5.3%) | 7 (36.8%) | 3 (15.8%) | 0 (0.0%) | 1 (5.3%) | |
| | | Stakeholder demand | 1 (5.3%) | 5 (26.3%) | 1 (5.3%) | 5 (26.3%) | 2 (10.5%) | 4 (21.1%) | 1 (5.3%) | |
| | | Technical / lab expertise | 2 (10.5%) | 3 (15.8%) | 1 (5.3%) | 0 (0.0%) | 6 (31.6%) | 3 (15.8%) | 4 (21.1%) | |

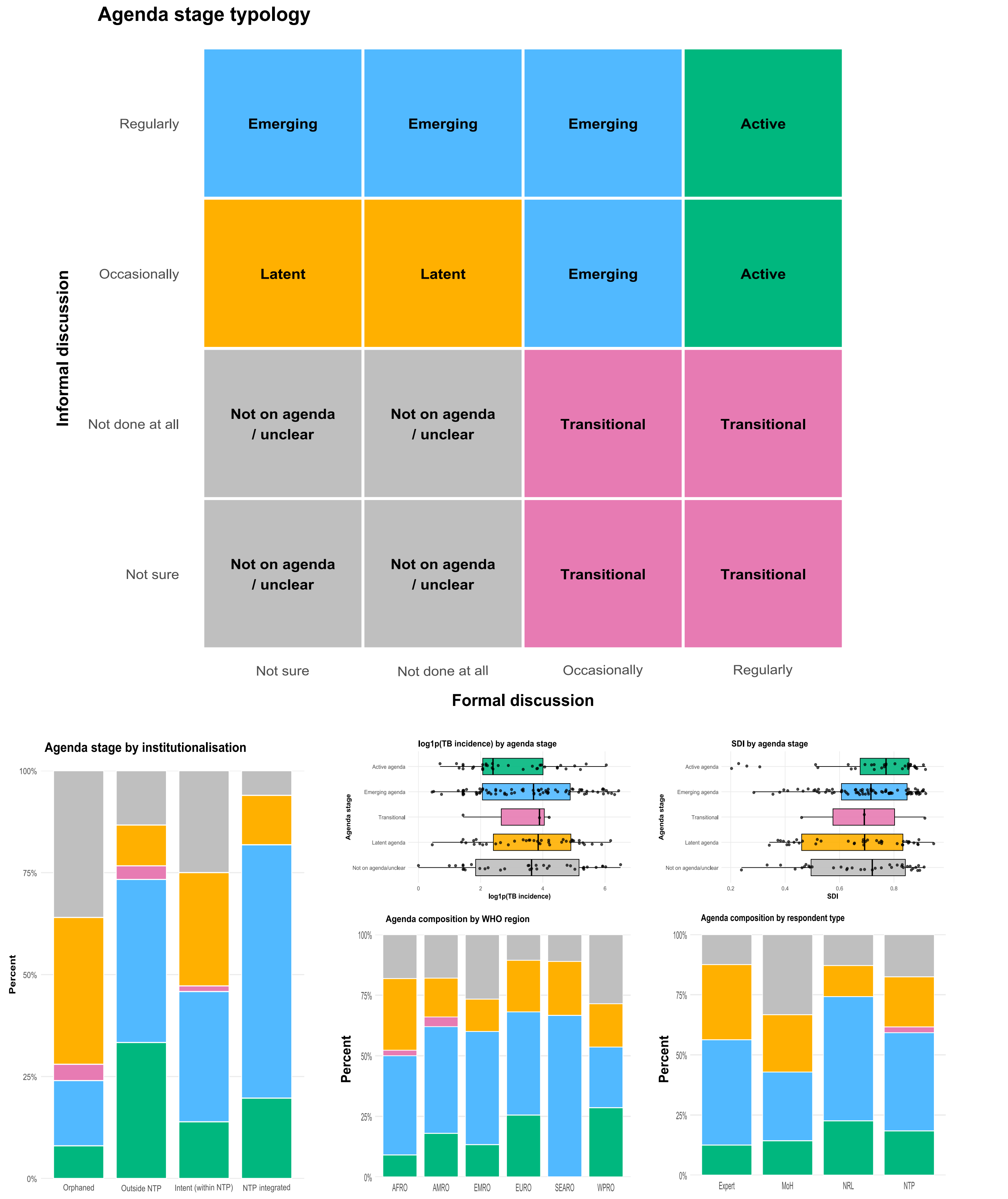


**Supplementary Figure S1. Agenda maturity typology and distribution across institutionalisation, epidemiologic context, region, and respondent type.** The upper panel shows the conceptual 4×4 agenda-stage typology derived from the cross-classification of formal discussion and informal discussion, each coded as not sure, not done at all, occasionally, or regularly. These combinations were grouped into five ordered agenda stages: not on agenda/unclear (gray), latent agenda (orange), transitional (pink), emerging agenda (blue), and active agenda (green). The lower panels summarise the empirical distribution of agenda stage across selected characteristics. The left panel shows the percentage distribution of agenda stages within collapsed institutionalisation categories. The upper middle and upper right panels show horizontal boxplots with overlaid jittered observations for log-transformed TB incidence [log1p(TB incidence)] and SDI, respectively, across agenda stages. The lower middle panel shows the % distribution of agenda stages within WHO regions. The lower right panel shows the percentage distribution of agenda stages within respondent types. In all stacked bar charts, bars sum to 100%, and colours are consistent across panels to represent the same agenda stages.

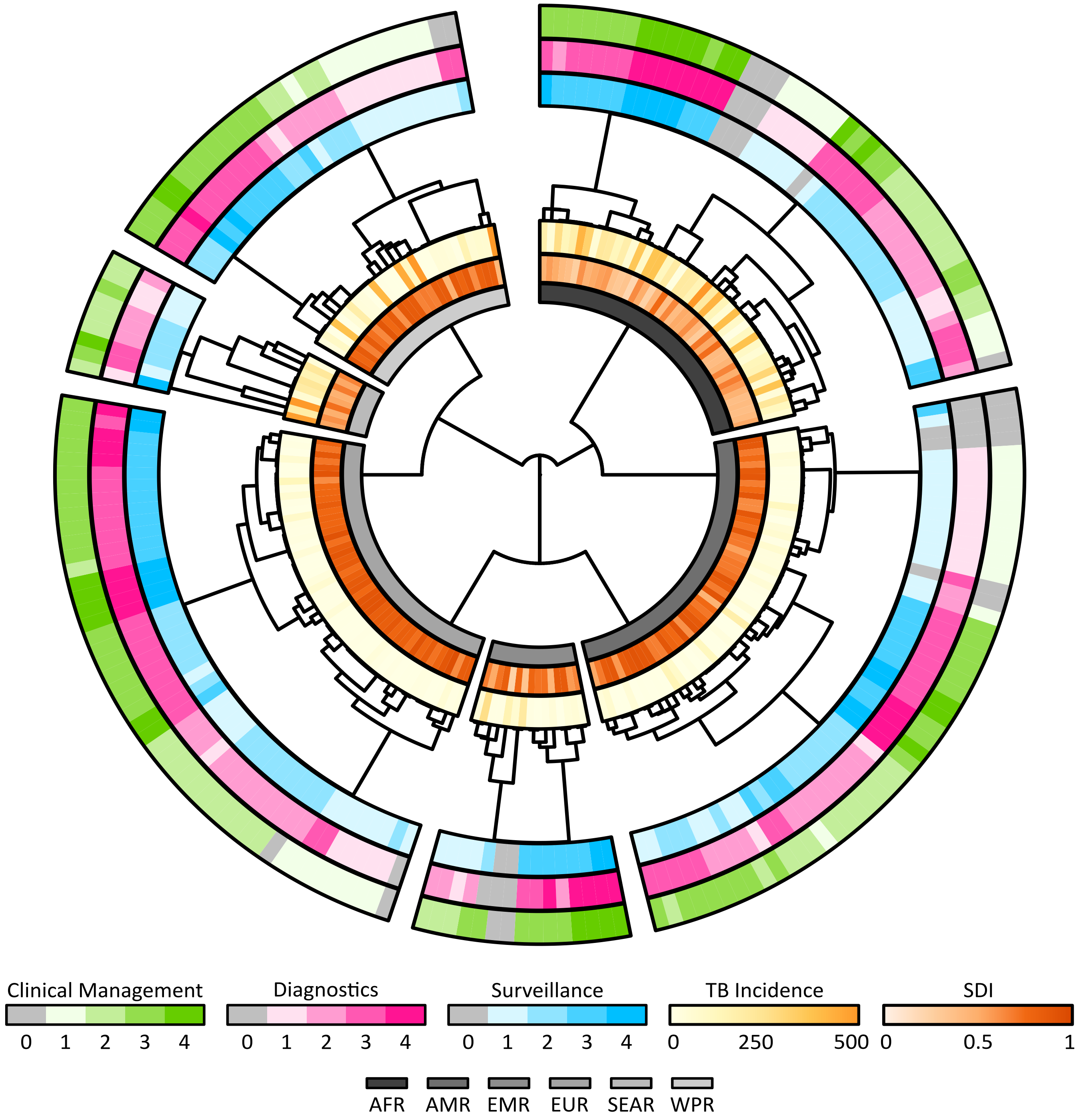


**Supplementary Figure S2. Perceived national policy urgency (relative momentum) for addressing NTM clinical management, diagnostics, and surveillance, by WHO region, estimated TB incidence, and socio-demographic index (SDI).** Each tile represents one country/state/jurisdiction, grouped by WHO region and ordered within regions using hierarchical clustering based on responses to three survey items: perceived urgency to address policy on NTM clinical management (green), diagnostics (pink), and surveillance (blue) at a national/state/jurisdictional level. Responses are encoded on a 0–4 scale (0 = Not Aware; 1 = Absent i.e., not on agenda; 2 = Low i.e., considered marginal; 3 = Moderate i.e., gaining attention; and 4 = High i.e., policy momentum), shown as shaded rings. Inner rings show estimated national TB incidence (all forms; 0-500 per 100 000 population; light to dark yellow) and SDI from Global Burden of Disease 2023 (0-1; light to dark coral). Peripheral dendrograms display within-region clustering (Euclidean distance; Ward.D2 linkage). The central fan clustering shows global clustering of WHO regions based on region-level response profiles, mean TB incidence, and mean SDI levels.

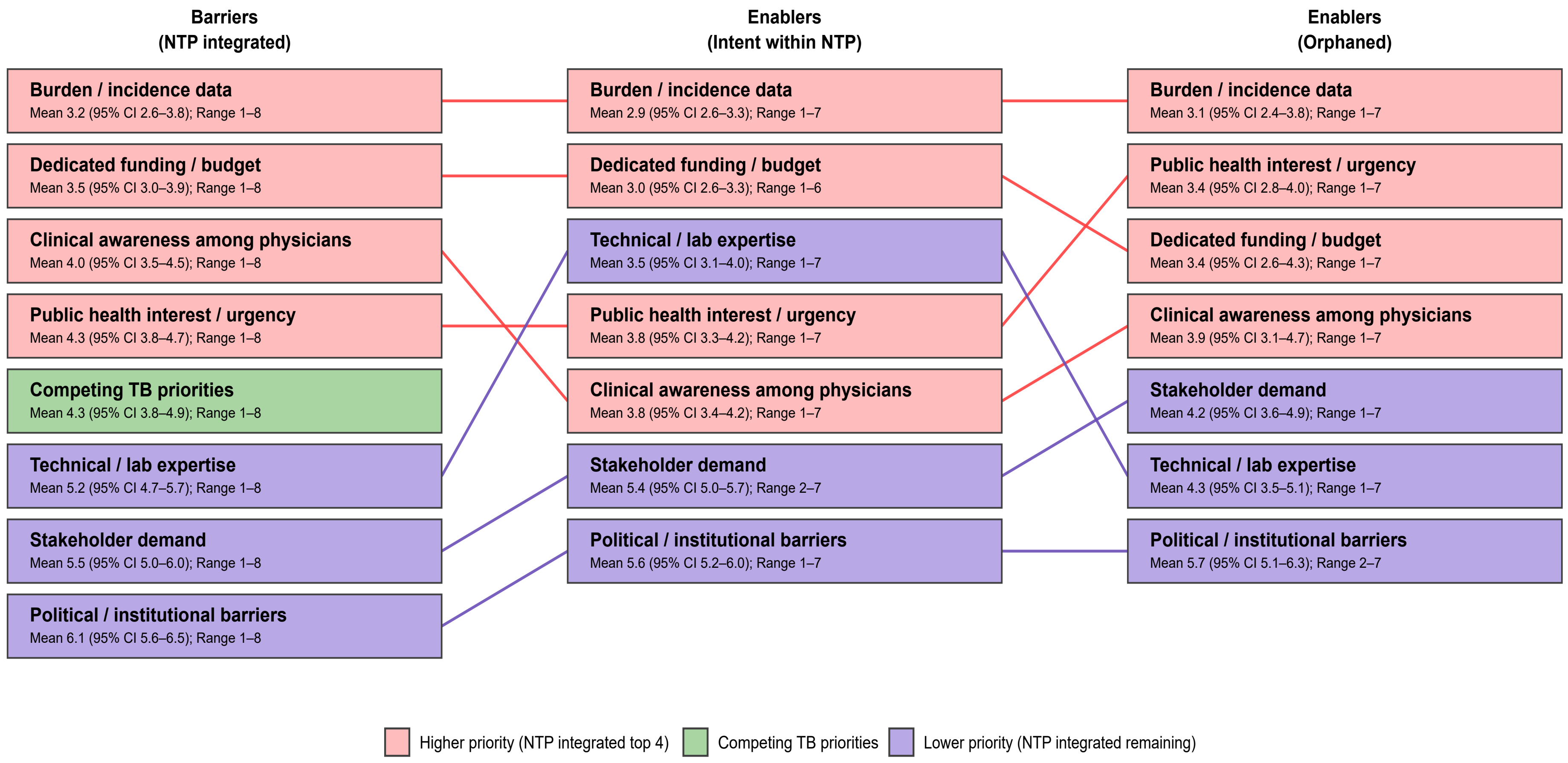


**Supplementary Figure S3. Stakeholder-ranked barriers and enablers for translating NTM discussions and policy urgency into implementable program action (global overall).** Boxes show the mean rank with bootstrapped 95% confidence intervals (CIs) and observed rank range for each theme. Boxes are further separated by Barriers (for NTP integrated settings) and Enablers for jurisdictions expressing Intent within NTP or Orphaned institutionalisation. Themes are ordered within each column by mean rank (lower = higher priority). Colours reflect the global Under-NTP ordering: pink = higher-priority themes, green = “Competing TB priorities”, and purple = remaining lower-priority themes. Connecting lines link the same theme across columns and are coloured by the same tier.

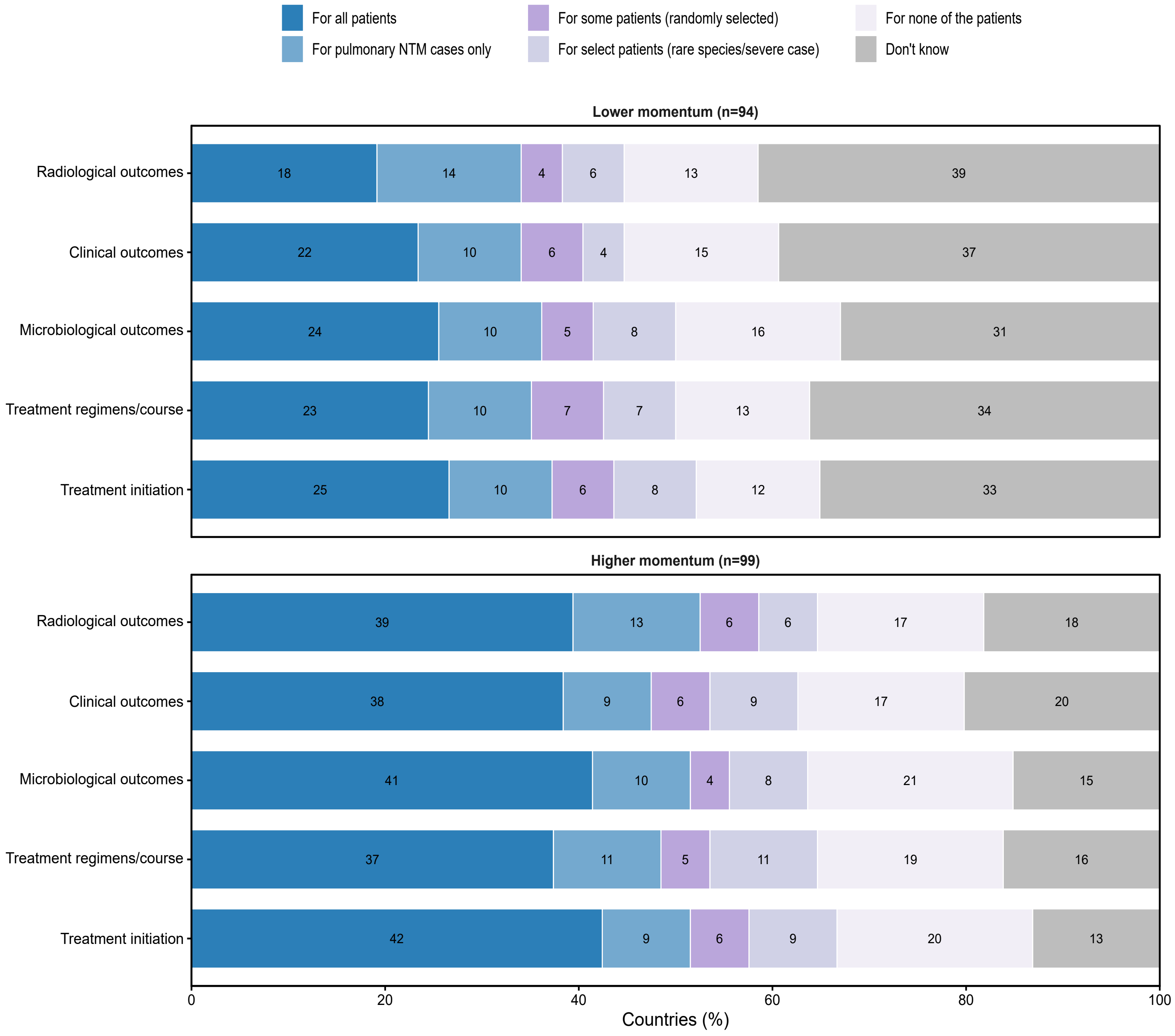


**Supplementary Figure S4. Reported recording of selected NTM clinical and laboratory trajectory elements, by relative policy momentum group.** Horizontal stacked bars show the distribution of responses to the survey question on whether specified elements of the clinical and laboratory trajectory of patients with presumed or confirmed NTM isolates are currently recorded. Response options ranged from recording for all patients to none of the patients, with additional intermediate and "don't know" categories. Numbers within segments indicate the number of jurisdictions selecting each response category; the n in each panel heading indicates the total number of jurisdictions in that momentum group.

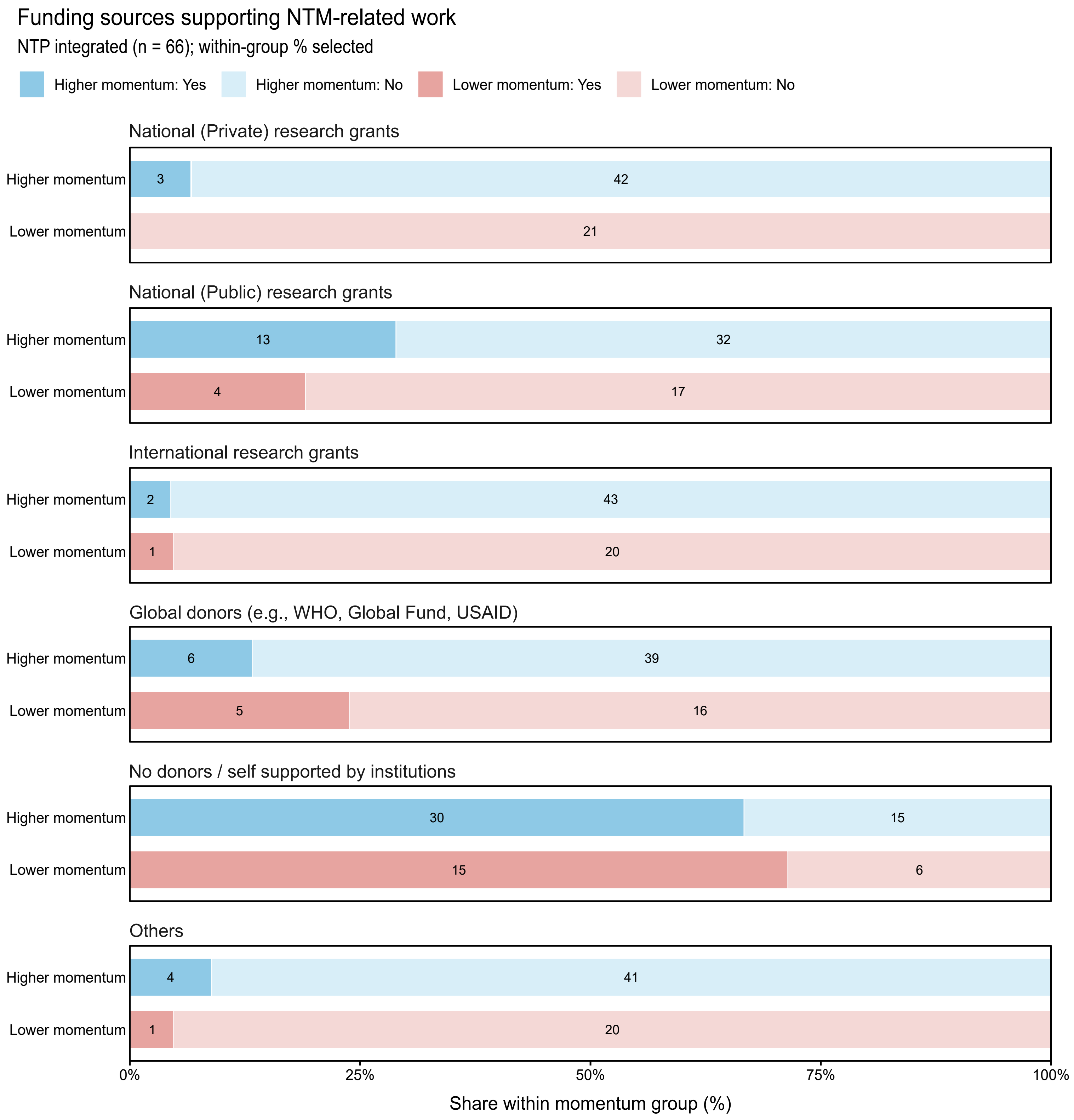


**Supplementary Figure S5. Funding sources currently supporting NTM-related work in jurisdictions with NTM integrated within the national TB programmes (NTPs), stratified by relative policy momentum.** Horizontal stacked bars display, for each funding source, the within-group proportion reporting that the source currently supports NTM-related activities at national level. Bars are shown separately for lower- and higher-momentum groups. Darker shades indicate "Yes" (selected), lighter shades indicate "No" (not selected); counts are printed within segments. Funding categories are not mutually exclusive (multiple selections were permitted).

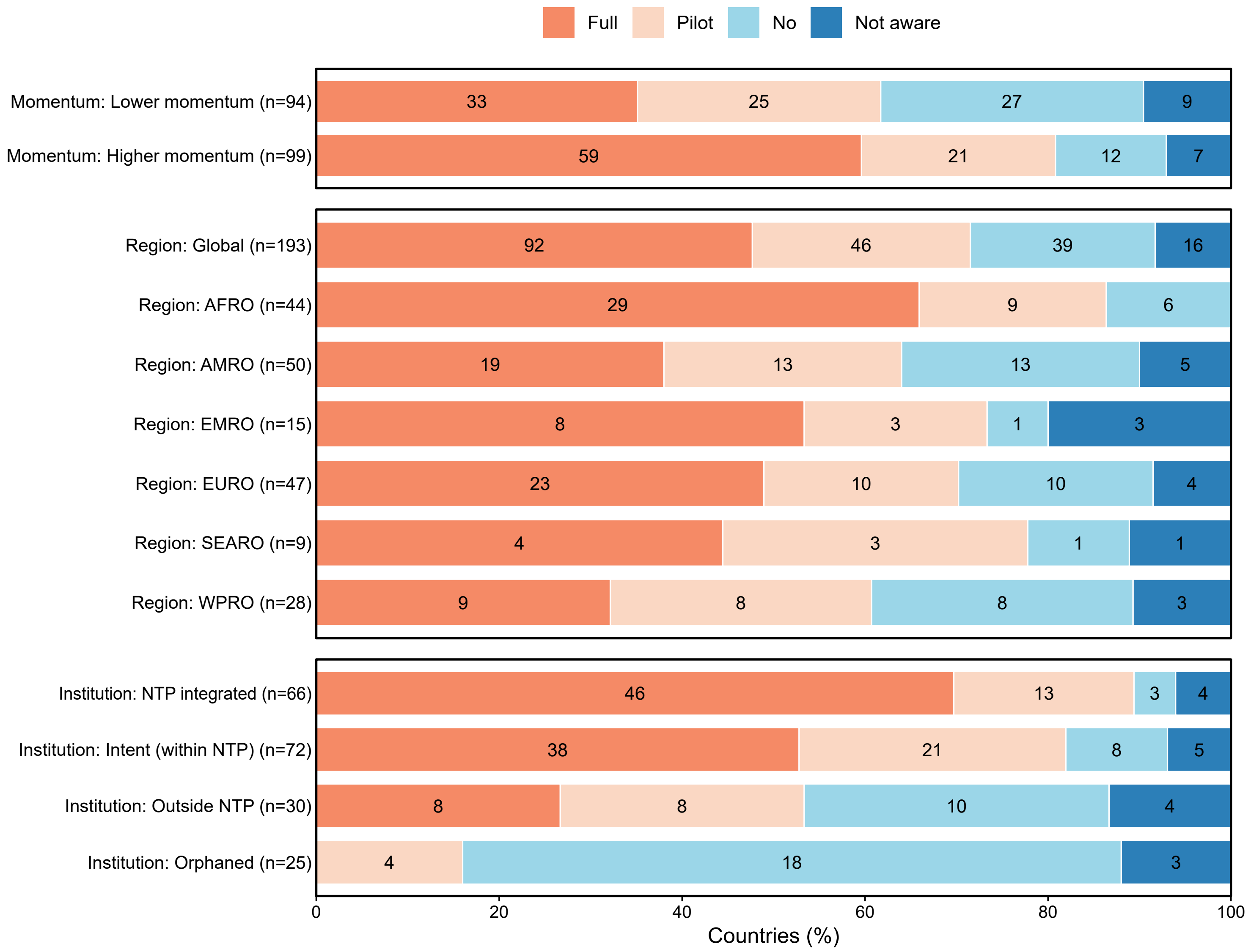


**Supplementary Figure S6. Reported willingness to support integration of NTM surveillance into national tuberculosis programmes (NTP) under current circumstances, by relative momentum, WHO region and institutionalisation.** Horizontal stacked bars show the percentage of responding jurisdictions selecting one of the options: full integration, pilot integration, no integration, or not aware. Numbers within segments indicate the count of jurisdictions selecting that response; n in each row label indicates the total number of jurisdictions in that stratum. Responses reflect stated willingness under current conditions rather than confirmed existing integration or implementation capacity.

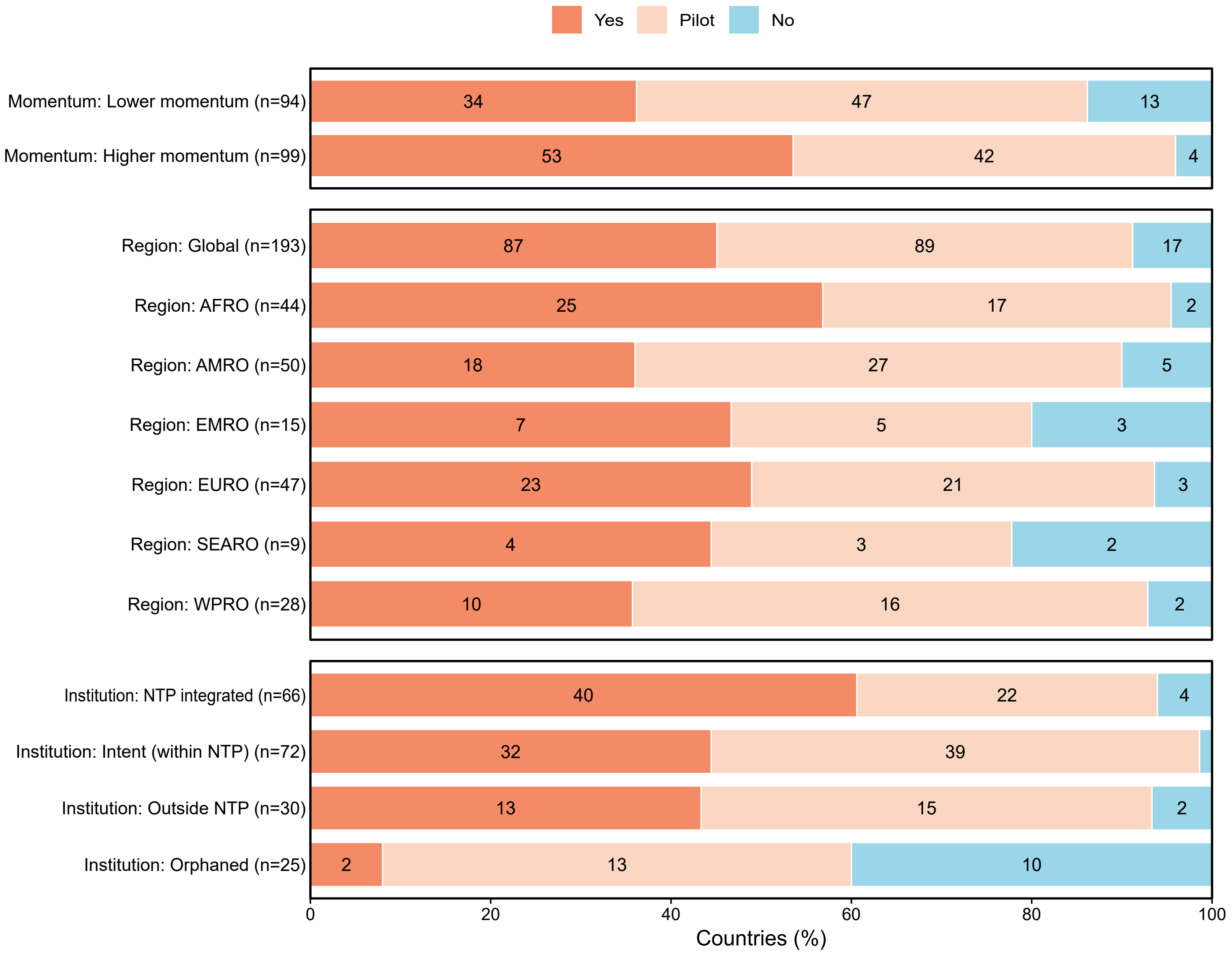


**Supplementary Figure S7. Reported willingness to support establishment of a global NTM surveillance effort or registry under current circumstances, by relative momentum, WHO region and institutionalisation.** Horizontal stacked bars show the percentage of responding jurisdictions selecting one of the options: Yes, Pilot, or No. Numbers within segments indicate the count of jurisdictions selecting that response; n in each row label indicates the total number of jurisdictions in that stratum. Note that the responses reflect stated willingness under current conditions rather than confirmed existing integration or implementation capacity.
